# Impact of gastric resection and enteric anastomotic configuration on delayed gastric emptying after pancreaticoduodenectomy: a network meta-analysis of randomized trials

**DOI:** 10.1101/2021.01.24.21250401

**Authors:** Chris Varghese, Sameer Bhat, Tim Wang, Gregory O’Grady, Sanjay Pandanaboyana

**Author notes:** Corresponding author Mr Sanjay Pandanaboyana, MS, FRCS, Mphil, Consultant surgeon, HPB and Transplant Unit, Freeman Hospital, Newcastle Upon Tyne, UK. **Funding** This study was supported by the New Zealand Health Research Council and the Royal Australasian College of Surgeons John Mitchell Crouch Fellowship. **Pre-registration:** A review protocol with analysis plan was submitted to PROSPERO (ID number 227637) on the 23 of December 2020.

## Abstract

**Introduction:** Delayed gastric emptying (DGE) is frequent after pancreaticoduodenectomy (PD). Several randomised controlled trials (RCTs) have explored operative strategies to minimise DGE, however, the optimal combination of gastric resection approach, anastomotic route, and configuration, role of Braun enteroenterostomy remains unclear.

**Methods:** MEDLINE, Embase, and CENTRAL databases were systematically searched for RCTs comparing gastric resection (Classic Whipple, pylorus-resecting, and pylorus-preserving), anastomotic route (antecolic *vs* retrocolic) and configuration (Billroth II *vs* Roux-en-Y), and enteroenterostomy (Braun *vs* no Braun). A random-effects, Bayesian network meta-analysis with non-informative priors was conducted to determine the optimal combination of approaches to PD for minimising DGE.

**Results:** Twenty-four RCTs, including 2526 patients and 14 approaches were included. There was some heterogeneity, although inconsistency was low. The overall incidence of DGE was 25.6% (n = 647). Pylorus-resecting, antecolic, Billroth II with Braun enteroenterostomy was associated with the lowest rates of DGE and ranked the best in 35% of comparisons. Classic Whipple, retrocolic, Billroth II with Braun ranked the worst for DGE in 32% of comparisons. Pairwise meta-analysis of retrocolic *vs* antecolic route of gastro-jejunostomy found increased risk of DGE with the retrocolic route (OR 2.1, 95% CrI; 0.92 - 4.7). Pairwise meta-analysis of Braun enteroenterostomy found a trend towards lower DGE rates with Braun compared to no Braun (OR 1.9, 95% CrI; 0.92 - 3.9). Having a Braun enteroenterostomy ranked the best in 96% of comparisons.

**Conclusion:** Based on existing RCT evidence, a pylorus-resecting, antecolic, Billroth II with Braun enteroenterostomy may be associated with the lowest rates of DGE.

## Introduction

Pancreaticoduodenectomy (PD) is commonly performed for benign and malignant pancreatic head and periampullary disease. There remains a high prevalence of postoperative delayed gastric emptying (DGE), affecting between 10-45% of patients ^1–5^. DGE is associated with poorer quality of life ^6^, increased hospital length of stay ^7^, readmissions ^8^ and healthcare costs ^9,10^. Technical approaches to gastric resection and reconstruction in PD are thought to impact the rates of DGE ^11^.

The recent pylorus resection or pylorus preservation (PROPP) RCT and several meta-analyses have shown comparable DGE rates between pylorus-preserving and pylorus-resecting PD ^3,12–14^. Pylorus resecting PD has been suggested to be favourable to pylorus preservation in reducing DGE rates as retaining the pylorus is thought to retain the propulsive activity of the stomach, but comparisons between the CW, PP and PR are limited ^3,12,15^. Previous meta-analyses also suggested antecolic gastro-jejunostomy to be the most effective route for minimising DGE ^16–18^, however, the evidence is conflicting ^19^. Similarly, it remains controversial whether Billroth II or Roux-en-Y reconstruction is favourable ^18,20–23^. The addition of a Braun enteroenterostomy, typically to prevent bilious reflux, has also been proposed to lower DGE rates ^24^.

Hence, it remains unclear which combination of gastric resection and anastomotic route and configuration is optimal for reducing DGE after PD. Traditional meta-analyses are limited to pairwise comparisons and large-scale surgical randomised controlled trials (RCTs) of all potential combinations are impractical. Network meta-analyses (NMA) allow comparison and ranking of a range of surgical approaches simultaneously, through direct and indirect comparisons. This NMA therefore aimed to identify the optimal combination of gastric resection, route of anastomosis, anastomotic configuration, and Braun enteroenterostomy on DGE and other complications after PD.

## Methods

This NMA was conducted per the PRISMA guidelines for NMAs ^25,26^. The study protocol was prospectively registered on PROSPERO (ID: xxxxx) before database searching.

### Literature search

A systematic search of MEDLINE (OVID), EMBASE (1980-2020), EMBASE Classic (1947-1973), and the Cochrane Controlled Register of Trials (CENTRAL) was conducted from their date of inception to December 2020. The following query terms were employed: the combined results of ‘pancreaticoduodenectomy’ OR ‘Whipple’ OR ‘gastroenteric’ OR ‘antecolic’ OR ‘retrocolic’ OR ‘Billroth’ OR ‘Roux-en-Y’ OR ‘pylorus-preserving’ or ‘antrectomy AND the combined results of ‘gastroparesis’ OR ‘delayed gastric emptying’ or ‘DGE’ AND the combined results of ‘trials’ OR ‘randomised’ OR ‘randomized controlled trial’. ‘Explode’ functions and Medical Subject Heading (MeSH) terms were used where available. There were no date or language restrictions. Article and review reference lists were also screened by two authors independently to identify additional potentially relevant RCTs. The full search strategy is available in **Appendix S1**.

### Inclusion and exclusion criteria

RCTs which described the gastric resection technique, route of gastro-jejunostomy (i.e. antecolic or retrocolic), the technique of reconstruction (i.e., Billroth II or Roux-en-Y), and addition of Braun enteroenterostomy in adults (>16 years old) undergoing PD were considered for inclusion. Only RCTs where DGE was either the primary endpoint or the study was adequately powered to detect a 20% difference in the rates of DGE between groups at 80% power and an alpha of 0.05 were eligible for inclusion (with a typical DGE incidence of ∼30% as per Hackert et al. ^3^). If aspects of the operative approach being investigated were unclear, individual authors were contacted for further details.

Studies were excluded if they were non-randomised, included paediatric populations (<16 years), were animal studies, or included operations other than PD. Only trials comparing surgical techniques were included. Trials comparing pharmacological interventions or methods of pancreatic anastomosis were excluded. Furthermore, trials where the surgeon could choose which method of gastric resection, route, or technique of anastomotic reconstruction and where these were not relatively homogenous, were excluded. Studies reporting duplicate outcomes from a previously published report were also excluded, to prevent multiple publication bias.

### Study selection and data extraction

Two authors screened titles and abstracts and reviewed full texts identified in the literature search for inclusion and extracted data onto a prespecified template independently. Discrepancies were discussed and resolved by mediation with a third independent reviewer. If there were missing data, the authors of the articles were contacted. We extracted the primary efficacy outcome data on rates of overall DGE as well as clinically relevant DGE (grade B/C defined as per the International Study Group of Pancreatic Surgery (ISGPS) ^27^). Secondary outcomes included the overall rates of postoperative pancreatic fistula (POPF) as well as clinically relevant POPF (grade B/C defined as per International Study Group of Pancreatic Fistula (ISGPF) ^28^), duration of operation, intraoperative blood loss, intra-abdominal abscess, wound infection, haemorrhage ^29^, hospital length of stay, reoperation, and mortality. The Cochrane Collaboration’s Risk-of-bias tool 2.0 ^30^ was used to assess study design.

### Terminology and definitions

Gastric resection was defined as Classic Whipple (CW), pylorus-resecting (PR) PD, and pylorus-preserving (PP) PD. CW included studies with antrectomy or 20-40% distal gastrectomy. PR PD (synonymous with subtotal stomach preserving PD) included studies where gastric resection occurred <3 cm proximal to the pyloric ring. This is physiologically rationalised by the proximal border of the terminal antral contraction being physiologically ~4-4.4 cm proximal to the pyloric ring ^31^. PP PD studies were defined based on gastric resection occurring at any distance distal to the pyloric ring. Route of anastomosis was defined as antecolic or retrocolic if the gastro-jejunostomy was anterior or posterior to the transverse colon, respectively. Reconstructions were defined as Billroth II if the pancreaticojejunostomy and hepatico-jejunostomy were proximal to an end-to-side gastro-junostomy in the absence of separate Roux and afferent limbs. The term Roux-en-Y reconstruction was used if a separate afferent and Roux limb were employed (e.g., to isolate pancreatic and hepatic outflows). Braun enteroenterostomy was defined as a jejunojejunostomy distal to the gastro-enteric anastomosis.

### Statistical analysis

A random-effects NMA was performed using gemtc in R ^32^ (R Foundation for Statistical Computing, Vienna, Austria); gemtc employs a Just Another Gibbs Sampler (JAGS) software to conduct arm-based calculations using a Bayesian framework and non-informative priors. Where studies reported medians, mean estimates were derived via the methods of Wan *et al*. and Luo *et al*. ^33,34^. Network maps were generated to visualise all direct comparisons made. Line thickness corresponded with the number of studies assessing a particular direct comparison and the size of nodes correlated with the number of participants receiving a particular intervention. A continuity correction of one was applied where a categorical outcome was achieved by none of the participants in a trial arm ^35^. Odds ratios (OR) were used for categorical outcome data, and mean differences (MD) for continuous data, both accompanied by 95 percent (%) credibility intervals (CrI). CW with antecolic gastrojejunostomy and Billroth II reconstruction without Braun enteroenterostomy was considered the comparator arm where applicable. Rankogram plots visualised the relative effectiveness of each intervention per outcome; they represent stacked bar plots of the probability of each intervention achieving each rank. Sum under the cumulative ranking (SUCRA) scores were used to rank interventions; where a score of 1 meant the intervention was the best ranked 100% of the time, and a score of 0 if it ranked as the worst intervention 100% of the time ^36^. Heterogeneity was assessed via the random-effects standard deviation as per Valkenhoef et al. ^37^. A node-splitting analysis of inconsistency was used to assess the comparability of indirect and direct comparisons. Transitivity was assessed by collecting and comparing the demographic data, surgical approach and co-interventions across direct comparisons. Comparison-adjusted funnel plots were constructed and visually inspected for asymmetry indicative of publication bias. A sensitivity analysis was performed by sequential removal of single studies to review the resulting discrepancies in rankings. A subgroup NMA and regression excluding studies that compared Braun *vs* no Braun enteroenterostomy was also performed. If 95% CrIs did not cross the no-effect line (0 for continuous outcomes and 1 for dichotomous outcomes), results were considered statistically significant.

## Results

Search results and study selection are summarised in **Figure 1**. Database search identified 1215 studies, of which 24 RCTs met the inclusion criteria for this NMA (**Appendix S2**). A total of 2647 patients were randomised, and 2526 patients were included in the analysis. These RCTs were published between 1999 and 2020 and compared 14 different combinations of gastric resection and anastomotic configurations after PD.

**Figure 1.**
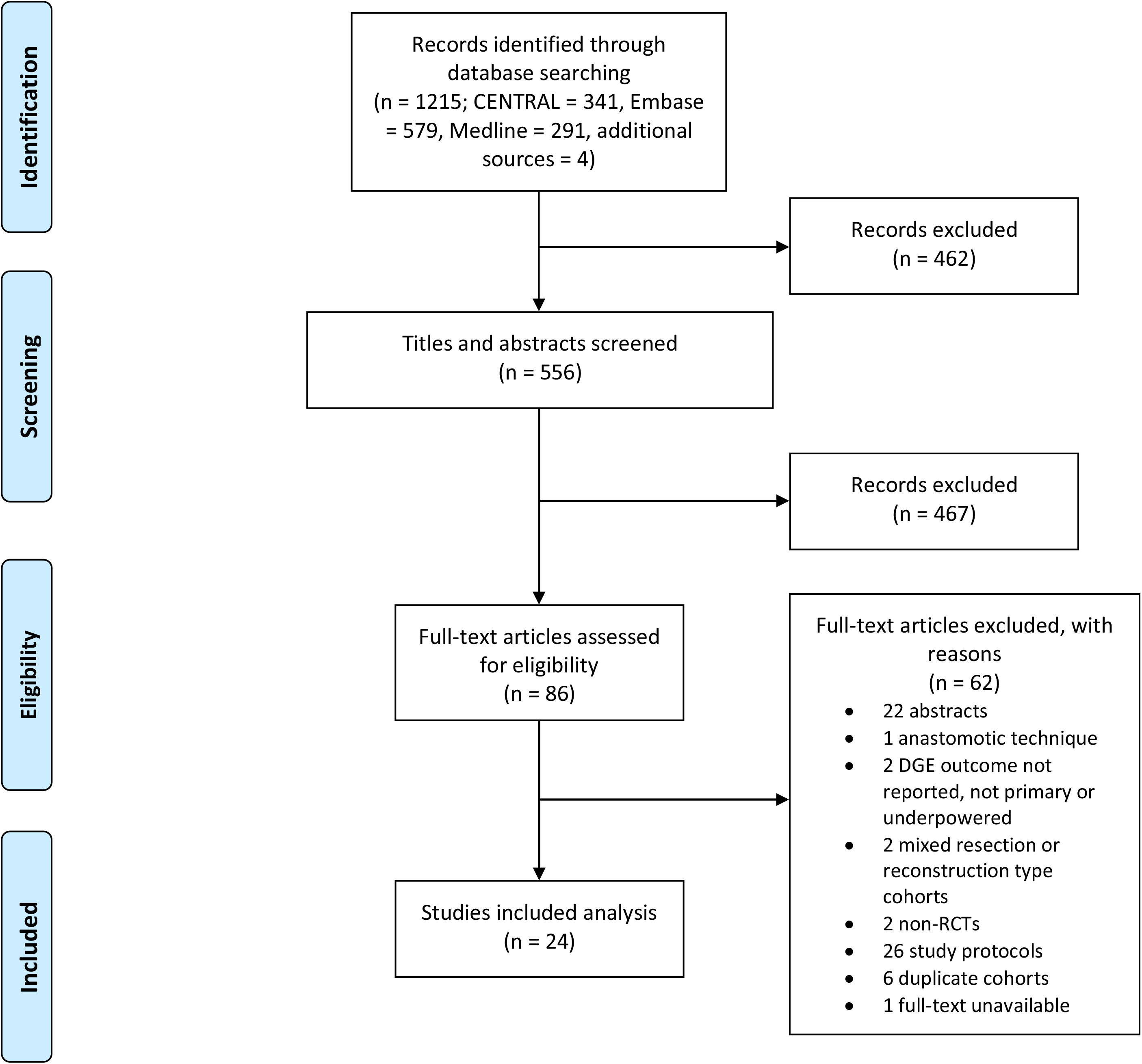
PRISMA flow diagram of study selection.

The direct comparisons from included RCTs are summarised in **Table 1**. The most common direct comparison (n = 4) was between PP PD with antecolic Billroth II reconstruction and PP PD with retrocolic Billroth II (both without Braun enteroenterostomy).

### Transitivity analysis

Definitions for the primary outcome of overall DGE were largely similar, however only 14 studies used the ISGPS definition. Similarly, 15 studies used the ISGPF definition for POPF. Only five studies reported the rates of prokinetic use after PD ^2–5,38^. Only three studies reported rates of parenteral nutrition (30-45%), with no differences between comparator arms ^3,39,40^. Ages, gender mix and BMI were generally comparable between direct comparisons, however, the single study comparing PR, retrocolic, Billroth II against PR, antecolic Billroth II had a male predominant cohort (>70%). Most studies were conducted in Asia (16/24; 67%) ^38,39,41–54^, 10 of which were in Japan ^41–43,46,47,49–53^. Study characteristics are summarised in **Table S1**.

### Risk of bias

Risk of bias assessments of these studies are shown in **Figure S1 and S2**. Four studies (16.7%) were considered to be at low risk-of-bias, with the remaining 20 (83.3%) considered to be at high risk-of-bias, predominantly due to the lack of blinding. Most studies demonstrated clear and efficacious randomisation, appropriate intention-to-treat analysis where possible, and transparent outcome reporting.

### Primary outcome measure

#### Overall delayed gastric emptying

Twenty-four trials comparing 14 combinations of PD in 2526 patients reported rates of overall DGE. The overall incidence of DGE was 25.6% (647/2526). The overall incidence of clinically relevant DGE was 14.9% (253/1698). **Table 2** summarises the results of the NMA from the direct comparisons of overall DGE rates. PP, antecolic, Billroth II *vs* PP, retrocolic, Billroth II had the most direct comparisons (**Figure 2**). Rankograms showed that PR, antecolic, Billroth II with Braun enteroenterostomy was associated with the lowest rates of overall DGE (**Figure 3**); this was followed by CW, antecolic, Billroth II with Braun enteroenterostomy. PR, antecolic, Billroth II with Braun enteroenterostomy ranked the best approach in 35% of comparisons. CW, retrocolic, Billroth II with Braun ranked the worst approach for overall DGE (in 32% of comparisons) (**Appendix S3**).

**Figure 2.**
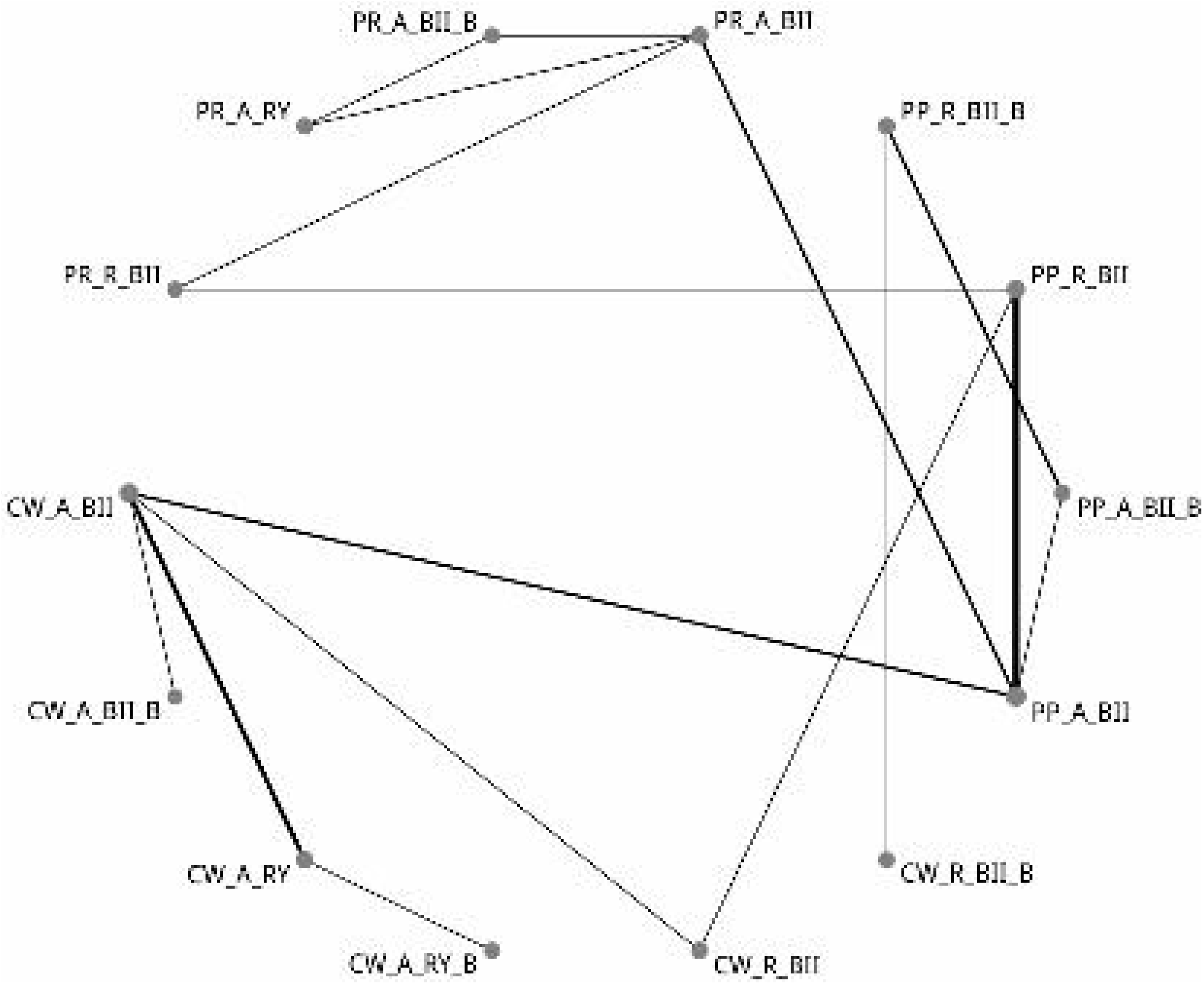
Network map for overall delayed gastric emptying. PP, pylorus preserving; CW, classic Whipple; PR, pylorus resecting; A, antecolic; R, retrocolic; BII, Billroth II; RY, Roux-en-Y; B, Braun enteroenterostomy.

**Figure 3.**
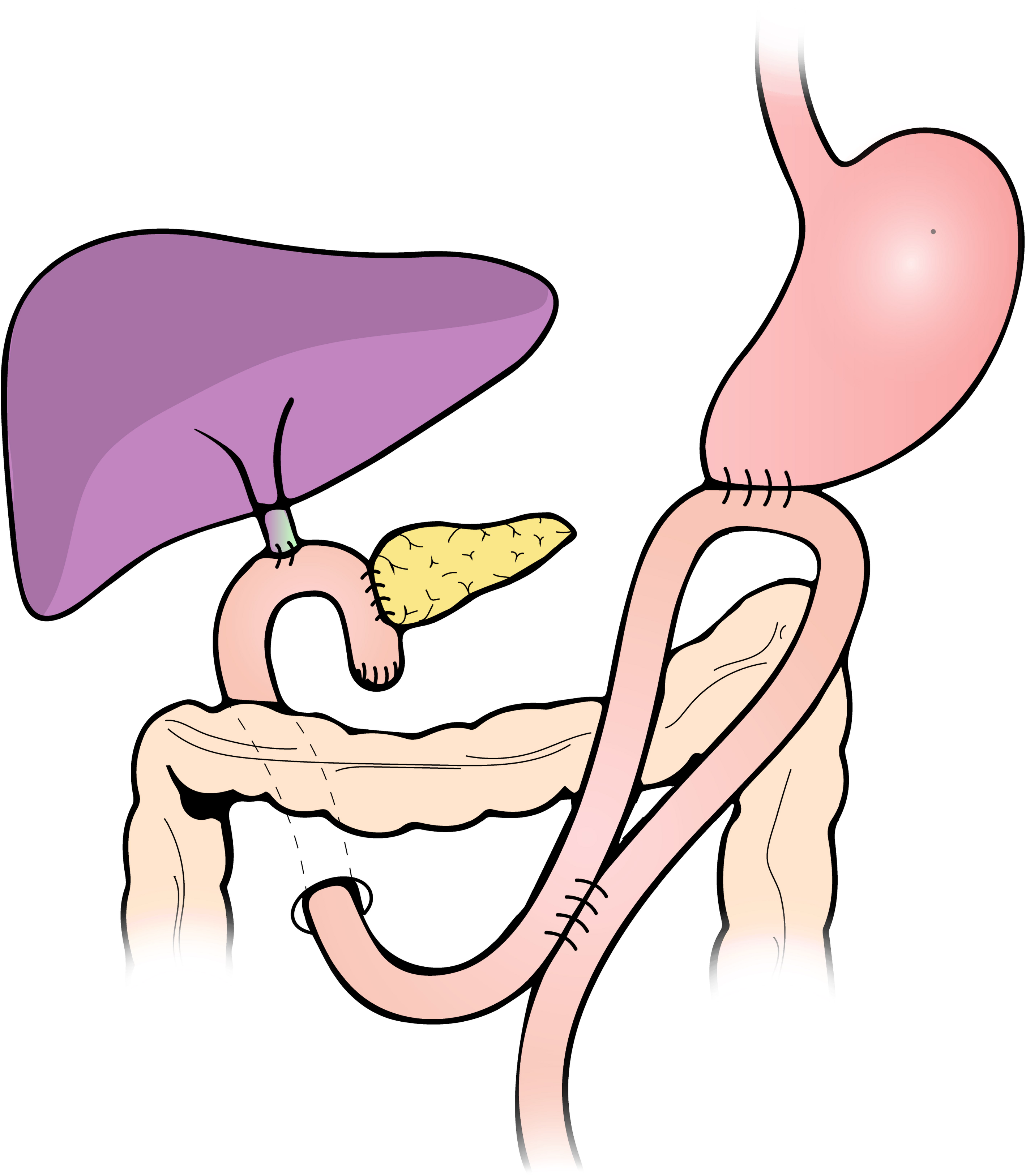
Schematic diagram of pylorus resecting, antecolic, Billroth II anastomosis with Braun enteroenterostomy.

### Secondary outcome measures

#### Overall postoperative pancreatic fistula

Overall POPF rates were reported in 22 trials comparing 13 approaches amongst 2472 patients. The overall incidence of POPF was 23.2% (571/2466). PP, retrocolic Billroth II vs PP antecolic Billroth II, both without Braun was the most frequent direct comparison. Rankograms showed that CW, retrocolic, Billroth II with Braun enteroenterostomy was associated with the lowest rates of POPF (in 79% of comparisons); this was followed by PR, antecolic, Roux-en-Y without Braun. However, CW, retrocolic, Billroth II with Braun was only reported in a single study ^55^. Hence, a sensitivity analysis excluding this study showed that PP, retrocolic, Billroth II with Braun was associated with the lowest POPF rates in 51% of comparisons, followed again by PR, antecolic, Roux-en-Y without Braun in 26% of comparisons. CW, antecolic, Roux-en-Y with Braun ranked the worst approach in 28% of comparisons and persisted as the worst ranked approach following sensitivity analysis (25% of comparisons) (**Appendix S3**).

#### Intra-abdominal abscess

Eighteen trials reported intra-abdominal abscess rates with direct comparisons of 11 approaches comprising 2105 patients. The overall incidence of intra-abdominal abscesses was 13.6% (285/2103). Billroth II *vs* Roux-en-Y configuration of CW with antecolic gastrojejunostomy and without Braun was the most frequent direct comparison. PR, antecolic, Roux-en-Y without Braun ranked best in 33% of direct comparisons followed by PP, retrocolic, Billroth II with Braun in 30% of direct comparisons. Because PR, antecolic, Roux-en-Y without Braun was only reported in a single study ^52^, a sensitivity analysis with this study removed resulted in PP, retrocolic, Billroth II with Braun ranking first in 33% of comparisons. PR, retrocolic, Billroth II without Braun was consistently associated with higher rates of intra-abdominal abscesses (63% of direct comparisons in both the overall and sensitivity analysis) and had significantly higher rates compared to PR, antecolic, Roux-en-Y without Braun (OR 9.3, 95% CrI; 1.1 to 136.5) and PR, antecolic, Billroth II without Braun (OR 6.0, 95% CrI; 1.1 to 65.8).

#### Postpancreatectomy haemorrhage (PPH)

Direct comparison of PPH rates were reported in 15 trials, comparing 11 approaches amongst 1689 patients. The overall incidence of PPH was 6.2% (105/1683). Rankograms showed comparable PPH rates with PP, retrocolic, Billroth II with Braun (ranked best in 29% of comparisons) and CW, antecolic, Billroth II with Braun (ranked best in 27% of comparisons). PR, antecolic Roux-en-Y ranked worst for PPH in 61% of comparisons; however, was only reported in one study. On sensitivity analysis, PP, retrocolic, Billroth II without Braun ranked worst in 27.6% of comparisons.

#### Duration of operation

Duration of operation was reported in 22 trials comprising 2252 patients and 13 approaches. Compared to CW, antecolic, Billroth II without Braun, both PP, retrocolic, Billroth II with Braun (MD -71 minutes, 95% CrI; -141 to -3.7) as PP, antecolic, Billroth II without Braun (MD - 30 minutes, 95% CrI; -58 to -4.4) were significantly quicker. PP, retrocolic, Billroth II with Braun also took significantly less time than both CW, retrocolic, Billroth II with Braun (MD - 66.2 minutes, 95% CrI; -120.5 to -12.3) and CW, antecolic, Roux-en-Y (MD -79.8 minutes, 95% CrI; -152.8 to -10.6). PP, retrocolic, Billroth II with Braun ranked best for operative duration in 82% of comparisons. CW, antecolic, Billroth II with Braun ranked worst in 32% of comparisons.

#### Intraoperative blood loss

Twenty trials reported comparative intraoperative blood loss across 13 approaches in 2214 patients. Antecolic *vs* retrocolic PP, Billroth II without Braun was the most common direct comparison. PP, retrocolic, Billroth II with Braun ranked the best for minimising intraoperative blood loss in 57% of all comparisons, followed by CW, antecolic, Billroth II with Braun, which ranked best in 21% of all comparisons. PR, retrocolic, Billroth II without Braun was associated with the largest volume of intraoperative blood loss in 46% of comparisons.

#### Mortality

Twenty-four trials reported mortality, with direct comparisons of all 14 approaches, including 2526 patients. The overall incidence of mortality was 2.1% (54/2526). Antecolic *vs* retrocolic, PP, Billroth II without Braun was the most frequent direct comparison. CW, antecolic, Billroth II with Braun had the lowest mortality rate in 27% of direct comparisons and CW, retrocolic, Billroth II with Braun had the worst mortality rate in 27% of direct comparisons.

#### Length of stay

Twenty trials reported length of stay (days) with direct comparisons of 12 approaches, including 2331 patients. Billroth II *vs* Roux-en-Y configuration for CW, antecolic, without Braun was the most frequent direct comparison. CW, retrocolic, Billroth II with Braun had the shortest length of stay in 63% of all comparisons, followed by PP, retrocolic, Billroth II with Braun in 25% of all comparisons. PR, antecolic, Roux-en-Y was associated with the longest length of stay in 36% of all comparisons. PR, antecolic, Roux-en-Y was associated with significantly longer length of stay compared to the best ranked approach: CW, retrocolic, Billroth II with Braun (MD 13.1 days, 95% CrI; 0.1 to 27).

Results of further secondary outcomes such as bile leak, wound infection, and reoperation rates can be found in **Appendix S3**.

### Subgroup analyses

#### Impact of Braun enteroenterostomy on DGE

Pairwise meta-analysis of Braun enteroenterostomy found a trend towards lower DGE rates with Braun compared to no Braun (OR 1.9, 95% CrI; 0.92 - 3.9). Having a Braun enteroenterostomy ranked the best approach in reducing DGE in 96% of comparisons. Results of a subgroup analysis excluding Braun comparisons showed PR, antecolic, Billroth II to rank the best in reducing DGE (**Appendix S3)**.

#### Impact of pyloric resection on DGE

When including studies which directly compared CW, PR and PP approaches to gastric resection in PD; no statistically significant differences in the rates of overall DGE were found. However, PR PD ranked as the best approach for reducing DGE in 71% of comparisons.

#### Impact of Antecolic vs retrocolic gastrojejunostomy on DGE

Pairwise meta-analysis of antecolic *vs* retrocolic route of gastro-jejunostomy found increased risk of DGE with the retrocolic compared to the antecolic route (OR 2.1, 95% CrI; 0.92 - 4.7). While this did not reach statistical significance, antecolic anastomosis ranked the best route for reducing DGE in 98% of comparisons.

#### Impact of Billroth II vs Roux-en-y reconstruction on DGE

Pairwise meta-analysis of Billroth II *vs* Roux-en-Y anastomotic configuration found no discernible differences in the rates of DGE (OR 1.2, 95% CrI; 0.59 - 2.3). Billroth II ranked the best configuration for reducing DGE in 68% of comparisons.

These results are congruent with PR, antecolic, Billroth II with Braun enteroenterostomy being the optimal combination of PD approaches to minimise DGE, demonstrating consistency with the NMA results.

### Risk of heterogeneity, inconsistency and publication bias

Some heterogeneity was found for the outcomes; rates of overall DGE, PPH, intraoperative blood loss, wound infection, bile leak, and in the separated NMA assessing gastric resection comparisons only. No significant heterogeneity was identified in the other outcome data. Node-splitting analysis models demonstrated no inconsistency in most outcomes except length of stay. This outcome should therefore be interpreted with caution. Heterogeneity and inconsistency results are summarised in **Appendix S4**. Comparison adjusted funnel plots showed an even distribution of studies adjacent to the pooled estimate line for most outcomes (**Appendix S3**). However, some funnel plot asymmetry, indicative of publication bias, was evident in trials comparing antecolic *vs* retrocolic CW, Billroth II without Braun and antecolic *vs* retrocolic PP, Billroth II without Braun for the primary outcome of overall DGE incidence.

## Discussion

The present NMA compared the impact of gastric resection, route and configuration of enteric anastomosis, and addition of Braun enteroenterostomy on DGE following PD. DGE occurred in nearly a quarter of patients in the included RCTs. PR, antecolic, Billroth II with Braun enteroenterostomy ranked best for minimising DGE, in both the overall NMA as well as in a subgroup analyses of techniques comparing each aspect of PD independently (i.e., gastric resection, CW *vs* PR *vs* PP; route of anastomosis, antecolic vs retrocolic; anastomotic configuration, Billroth II vs Roux-en-Y; and the presence *vs* absence of Braun enteroenterostomy). PP, retrocolic, Billroth II with Braun enteroenterostomy ranked best on sensitivity analysis for minimising complication rates such as POPF, intra-abdominal abscesses, bile leak, PPH, duration of operation and intraoperative blood loss. Postoperative complications such as POPF and intra-abdominal abscesses are known risk factors for DGE ^56^, and optimal approaches to PD should ideally minimise all of these complications. CW, retrocolic, Billroth II, with Braun ranked worst for DGE and mortality, while PR, retrocolic, Billroth II without Braun ranked worst for intra-abdominal abscess, bile leak, intraoperative blood loss and reoperation.

The mechanisms of DGE after PD remain unresolved but are multifactorial. Physiologically, gastric accommodation and the resultant common cavity pressure gradient between stomach and small bowel play a key role in the emptying of liquids (the ‘pressure pump’), whereas antral contractions are more important in the emptying of solids (the ‘peristaltic pump’) ^57^. Myenteric factors (circular smooth muscle and interstitial cells of Cajal (ICC)), vagal innervation, antropyloric coordination, and hormonal influences (inhibitors of GE: cholecystokinin, GLP-1; promoters of GE: motilin and ghrelin ^58^) all play essential contributing and/or regulatory roles ^59^. The combination of partial vagotomy, distal gastric resection, gastro-jejunostomy in PD predisposes patients to delayed gastric emptying ^60^. Additionally, reduced motilin arising from the duodenal resection likely further contributes to retention through suppression of the migrating motor complex ^61^.

The distal gastric antrum is electrically and functionally distinct to the rest of the stomach, with a specific bioelectrical activity that coordinates the ‘terminal antral contraction’, being important for trituration and solid gastric emptying ^31,62^. CW, with resection of the distal ∼4cm of the stomach, could therefore result in impaired solid gastric emptying due to loss of the terminal antral contraction ^31^. Alternatively, potential denervation of the antroduodenal complex as in PR PD could contribute to retention of a relatively hypo-motile distal antral remnant that could be counter-productive for gastric emptying, and some authors have advocated that complete antropyloric resection (as in CW) may be preferable for reducing DGE, as supported by Sun *et al* and Yeo *et al* ^11,63^. This proposed advantage of CW was not evident in three other RCTs ^40,48,64^ nor our results, with the highest ranked technique associated with reduced DGE being a PR PD technique. Experimental evidence suggests coordination of contractions between antrum and pylorus, and pylorus and duodenum is less important for gastric emptying compared to the depth of antral contractions (important for trituration) and receptive relaxation of the small bowel ^65^. In PR PD, these requirements are relatively maintained in comparison with CW.

The route of gastro-or duodenojejunostomy (antecolic *vs* retrocolic), anastomotic reconfiguration method (Billroth II *vs* Roux-en-Y), and addition of Braun enteroenterostomy have been commonly investigated for their role in DGE ^18,66^, but there is a paucity of physiological literature justifying specific mechanisms. Some hypothesise excess torsion or poor angulation of the gastro-enteric anastomosis may contribute to DGE through mechanical obstruction, but this requires further exploration in future studies ^67^. The poor performance of approaches that use retrocolic gastro-enteric anastomosis for DGE in our study may support this hypothesis, potentially due to the possibility of mechanical obstruction through a tight transmesocolic window ^47^. Furthermore, bile reflux through gastric mucosal irritation also likely contributes to DGE ^68^; the addition of a Braun enteroenterostomy allows for bilious diversion, provides two paths for gastric contents and stabilises the gastro-/duodeno-enteric anastomosis reducing torsion and risk of gastroenteric obstruction. Braun likely also reduces pressure on the small bowel side of the anastomosis, which is important for permitting gastric emptying ^65^. These mechanisms may explain why Braun was favourable for DGE and other complications in our NMA. These hypotheses require further investigation. A recent report also raised the possibility of aberrant conduction pathways arising from interstitial cells of Cajal (ICC) regrowth in small-bowel to stomach anastomosis ^69^, potentially contributing to DGE regardless of choice of anastomotic configuration, but further clinical data is needed to assess whether this is a factor after PD.

Another key consideration in mitigating DGE after PD is the incidence of postoperative complications. Complications, in particular POPF, have been well described as a major risk factor for DGE after PD ^8,56,70–73^. Therefore, an approach to PD which also minimises other complications should also be considered important when aiming to mitigate DGE. In this network meta-analysis, PP, retrocolic, Billroth II with Braun enteroenterostomy was consistently shown to rank the best in minimising rates of POPF, intra-abdominal abscesses, bile leak, PPH, and intraoperative blood loss. Interestingly, this contrasted with the optimal approach for minimising DGE (PR, antecolic, Billroth II with Braun enteroenterostomy). One potential explanation for the preference of the antecolic route in DGE could be the separation from the pancreas afforded by the transverse colon; protecting against the negative local inflammatory consequences of POPF on DGE ^19^. Importantly, the severity of complications have also correlated with DGE ^74^, hence future trials should conform to standardised reporting guidelines set out by the Clavien Dindo system, ISGPS and ISGPF, to allow for comparisons between trial cohorts ^27,28^. Furthermore, the included RCT’s in the present NMA were powered to detect a difference in DGE rates rather than POPF rates.

When interpreting these results and the above discussion points, it is necessary to reflect on PD as a complex procedure with many steps that could influence the likelihood of developing DGE. Previous literature has attempted to ascertain an approach at each step of PD that is optimal for minimising DGE. A few meta-analyses have suggested antecolic, Billroth II to be favourable for DGE ^16–18^. Of meta-analyses that specifically investigated the resection of the pylorus; Klaiber et al. found no difference between PR and PP PD in a subgroup analysis of RCTs ^12^, further confirmed by this NMA. However, when these authors pooled non-randomized studies, in a similar approach to Huang et al., both found PR PD may be preferred for minimising DGE ^12,15^. Wu et al. compared PR, CW and PP PD in a series of subgroup analyses and found PR, but not CW, to improve DGE compared to PP ^13^. However, inclusion of non-RCT data risks significant confounding and selection bias necessitating confirmation of these results using RCT data. Furthermore, where more than two operative options exist at each stage of PD, pairwise meta-analysis is unable to provide a suitable answer. The present study employed a Bayesian NMA of RCTs to compare 14 different approaches to gastric resection and enteric anastomotic route and configuration in PD. This methodology is advantageous as it allowed simultaneous comparison of all randomised data available on this topic, while maintaining randomisation during pooled analysis ^75^. Direct and indirect comparisons from the available RCT literature allowed for accurate NMA as evidenced by the lack of inconsistency in all outcomes except length of stay. This is especially relevant given the difficulty of conducting large-scale, high-quality RCTs comparing all combinatorial approaches to PD.

This review has several limitations. Most studies were deemed to be at high-risk of bias, however this was predominantly due to limited blinding of surgeons and investigators collecting outcome data. While the majority of RCTs employed randomisation intraoperatively prior to the procedure step under investigation, there remains a risk of selection and detection bias due to the limited blinding. Otherwise, there were minimal causes of concern as most studies conducted intention-to-treat analysis, demonstrated adequate and transparent randomisation, patient blinding, and reported outcomes in-full, with little missing data. Indications for PD, aspects of postoperative care (in particular the use of prokinetics), Clavien Dindo complication grades, and the rates of postoperative enteral nutrition were variably and poorly reported in the included trials. These factors may be a potential modifier of treatment effects, thus contributing to unquantified transitivity in this NMA ^29,76^. These outcomes should therefore be more consistently reported in future RCTs, in particular those investigating DGE. Analysis of clinically relevant DGE (defined as Grade B or C ^27^), could not be performed due to insufficient studies. Three studies had some degree of cross-over between PP and CW arms, in particular in Eshuis et al ^2,5,63^. One study that would otherwise have met the inclusion criteria for this NMA was excluded due to patients being stratified to either CW or PP in both arms (comparing antecolic *vs* retrocolic routes), with breakdown of data unavailable upon contacting authors. While many trials focused on comparing a single aspect of PD; adequate consistency between trial arms pertaining to other aspects of PD allowed for such an NMA to be performed. This highlights a strength of the NMA methodology to perform comparisons not feasible in a trial setting. The influence of each combination of approaches to PD on DGE was able to be assessed to an extent not previously attempted. This NMA did not include approaches to the pancreatico-enteric anastomosis. Gastric and enteric resection and anastomotic factors were prioritised as they more directly impact on DGE. Some publication bias and heterogeneity, but no inconsistency was seen in the primary outcome of overall DGE. Furthermore, several direct comparisons were only informed by single RCTs, which may have introduced some bias.

In conclusion based on existing RCT evidence, a pylorus resecting, antecolic, Billroth II anastomosis with Braun enteroenterostomy is associated with the lowest rates of DGE after pancreatoduodenectomy.

## Data Availability

Data is available on request.

## Tables and Figures

**Table 1. Summary of included trials**

*Values are mean (SD). PP, pylorus preserving; CW, classic Whipple; PR, pylorus resecting; A, antecolic; R, retrocolic; BII, Billroth II; RY, Roux-en-Y; B, Braun enteroenterostomy.

**Table 2. Network meta-analysis for overall delayed gastric emptying rates**

CW, Classic Whipple; PP, pylorus-preserving; PR, pylorus-resecting; A, antecolic; R, retrocolic; BII, Billroth II; RY, Roux-en-Y; B, Braun enteroenterostomy.

**Table 3. Rank probability of being the best ranked approach for pancreaticoduodenectomy in direct and indirect comparison in the network meta-analysis**

*Results of sensitivity analysis performed by omitting trial with single comparison. †PD approach with the highest probability of ranking first.

