## Appendix for "Impact of gastric resection and enteric anastomotic configuration on delayed gastric emptying after pancreaticoduodenectomy: a network meta-analysis of randomized trials"

**Table of Contents**

***Appendix S1: Search Strategies	2***

***Appendix S2: References of Included RCTs	4***

***Appendix S3: NMA All Results	7***

***Appendix S4: Heterogeneity and inconsistency	61***

***Figure S1: Risk of bias summary graph	63***

***Figure S2: Risk of bias figure	63***

### Appendix S1: Search Strategies

**The search was conducted in December 2020.**

**Ovid MEDLINE(R) Epub Ahead of Print, In Process & Other Non-Indexed Citations, Ovid MEDLINE (R) Daily, and Ovid MEDLINE (R) 1946-Present**

1	exp Pancreaticoduodenectomy/ or exp Pancreatic Neoplasms/

2	pancreaticoduodenectom*.mp.

3	pancreatoduodenectomy.mp.

4	duodenopancreatectomy.mp.

5	pancreaticoduodenal resection.mp.

6	PPPD.mp.

7	(subtotal and stomach and preserving).mp. [mp=title, abstract, original title, name of substance word, subject heading word, floating sub-heading word, keyword heading word, organism supplementary concept word, protocol supplementary concept word, rare disease supplementary concept word, unique identifier, synonyms]

8	exp Pancreaticojejunostomy/ or pancreaticogastrostomy.mp.

9	pancreaticojejunostom*.mp.

10	(pancreaticoenteric anastomosis or (gastroenteric or gastro-enteric)).mp. [mp=title, abstract, original title, name of substance word, subject heading word, floating sub-heading word, keyword heading word, organism supplementary concept word, protocol supplementary concept word, rare disease supplementary concept word, unique identifier, synonyms]

11	(antecolic or ante-colic or retrocolic or retro-colic or Roux-en-Y or RY).mp. [mp=title, abstract, original title, name of substance word, subject heading word, floating sub-heading word, keyword heading word, organism supplementary concept word, protocol supplementary concept word, rare disease supplementary concept word, unique identifier, synonyms]

12	(standard and pancreaticoduodenectomy).mp. [mp=title, abstract, original title, name of substance word, subject heading word, floating sub-heading word, keyword heading word, organism supplementary concept word, protocol supplementary concept word, rare disease supplementary concept word, unique identifier, synonyms]

13	(pylorus preserving or pylorus-preserving).mp.

14	gastrojejunostomy.mp.

15	duodenojejunostomy.mp.

16	pancreaticogastrostomy.mp.

17	(pylorus resect* or pylorus-resect*).mp. [mp=title, abstract, original title, name of substance word, subject heading word, floating sub-heading word, keyword heading word, organism supplementary concept word, protocol supplementary concept word, rare disease supplementary concept word, unique identifier, synonyms]

18	antrectomy.mp.

19	(Whipple* and (surgery or procedure or resect*)).mp. [mp=title, abstract, original title, name of substance word, subject heading word, floating sub-heading word, keyword heading word, organism supplementary concept word, protocol supplementary concept word, rare disease supplementary concept word, unique identifier, synonyms]

20	gastrojejunostomy.mp.

21	exp Gastroenterostomy/ or gastroduodenostomy.mp.

22	enterostomy.mp. or exp Enterostomy/

23	(entero-enteric or enteroenteric).mp. [mp=title, abstract, original title, name of substance word, subject heading word, floating sub-heading word, keyword heading word, organism supplementary concept word, protocol supplementary concept word, rare disease supplementary concept word, unique identifier, synonyms]

24	duodenojejunostomy.mp.

25	jejunoduodenostomy.mp.

26	Billroth.mp.

27	braun.mp.

28	20 or 21 or 22 or 23 or 24 or 25 or 26 or 27

29	exp Gastroparesis/ or delayed gastric emptying.mp.

30	DGE.mp.

31	29 or 30

32	1 or 2 or 3 or 4 or 5 or 6 or 7 or 8 or 9 or 10 or 11 or 12 or 13 or 14 or 15 or 16 or 17 or 18 or 19

33	28 or 32

34	randomized controlled trial.pt.

35	controlled clinical trial.pt.

36	randomi?ed.ab.

37	randomi?ed.ab.

38	clinical trials as topic.sh.

39	randomly.ab. or random*.mp. [mp=title, abstract, original title, name of substance word, subject heading word, floating sub-heading word, keyword heading word, organism supplementary concept word, protocol supplementary concept word, rare disease supplementary concept word, unique identifier, synonyms]

40	trial.ti.

41	34 or 35 or 36 or 37 or 38 or 39 or 40

34	33 and 31 and 41

**The exact same search was adapted for EMBASE through the OVID platform. The following databases were searched:**

Embase <1980 to 2020 Week 51>

Embase Classic <1947 to 1973>

**Search terms for CENTRAL:**

pancreaticoduodenectom* or duodenopancreatectom* or pancreaticoduodenal resection or PPPD or ((subtotal or sub-total) and stomach and preserving) or Pancreaticojejunostom* or pancreaticogastrostom* or pancreaticoenteric or gastroenteric or (antecolic or ante-colic or retrocolic or retro-colic or Roux-en-Y or RY) or standard and pancreaticoduodenectomy or pylorus preserving or pylorus-preserving or gastrojejunostomy or duodenojejunostomy or pancreaticogastrostomy or pylorus resect* or pylorus-resect* or antrectom* or (Whipple* and (surgery or procedure or resect*)) or gastrojejunostomy or Gastroenterostomy or gastroduodenostomy or enterostomy or entero-enteric or enteroenteric or duodenojejunostomy or jejunoduodenostomy or Billroth or braun

AND

gastroparesis OR delayed gastric emptying OR DGE

### Appendix S2: References of Included RCTs

### Appendix S3: NMA All Results

**Main Analysis**

DGE Overall

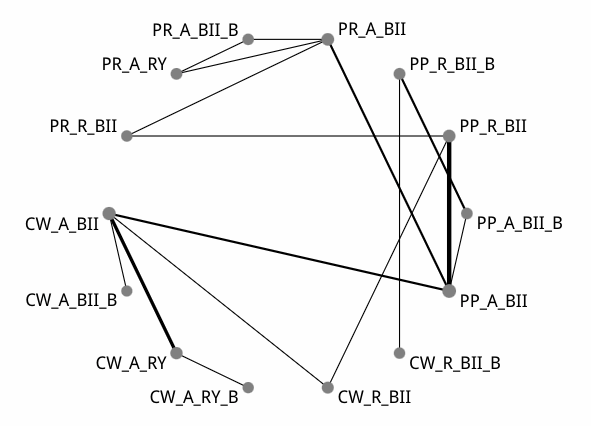

| Comparison of the included interventions: odds ratio (95% CrI). Each cell gives the effect of the column-defining intervention relative to the row-defining intervention. | | | | | | | | | | | | | |
| --- | --- | --- | --- | --- | --- | --- | --- | --- | --- | --- | --- | --- | --- |
| CW_A_BII | 0.559 ( 0.029, 10.810) | 0.821 ( 0.219, 2.873) | 0.723 ( 0.053, 9.255) | 3.053 ( 0.555, 21.929) | 2.467 ( 0.053, 146.790) | 1.169 ( 0.275, 6.128) | 0.485 ( 0.031, 8.600) | 1.993 ( 0.394, 13.807) | 1.306 ( 0.054, 44.590) | 0.626 ( 0.075, 4.906) | 0.295 ( 0.017, 5.200) | 1.003 ( 0.056, 16.617) | 1.588 ( 0.150, 19.773) |
|  | CW_A_BII_B | 1.494 ( 0.057, 36.730) | 1.294 ( 0.025, 65.694) | 5.455 ( 0.192, 185.490) | 4.304 ( 0.039, 712.728) | 2.105 ( 0.082, 63.156) | 0.846 ( 0.015, 54.549) | 3.642 ( 0.133, 125.914) | 2.340 ( 0.032, 227.102) | 1.107 ( 0.030, 40.117) | 0.526 ( 0.009, 32.382) | 1.755 ( 0.031, 106.740) | 2.822 ( 0.070, 122.768) |
|  |  | CW_A_RY | 0.883 ( 0.090, 7.932) | 3.677 ( 0.439, 40.956) | 2.938 ( 0.055, 248.837) | 1.429 ( 0.203, 11.895) | 0.581 ( 0.029, 13.918) | 2.431 ( 0.325, 25.524) | 1.574 ( 0.053, 66.374) | 0.768 ( 0.060, 9.082) | 0.358 ( 0.016, 9.049) | 1.239 ( 0.052, 30.554) | 1.884 ( 0.130, 33.023) |
|  |  |  | CW_A_RY_B | 4.214 ( 0.202, 122.217) | 3.362 ( 0.038, 469.139) | 1.639 ( 0.086, 36.694) | 0.658 ( 0.015, 32.214) | 2.813 ( 0.141, 76.846) | 1.805 ( 0.032, 150.100) | 0.887 ( 0.031, 25.795) | 0.418 ( 0.008, 21.700) | 1.396 ( 0.027, 75.196) | 2.217 ( 0.065, 80.022) |
|  |  |  |  | CW_R_BII | 0.814 ( 0.012, 51.367) | 0.387 ( 0.050, 2.489) | 0.158 ( 0.006, 2.926) | 0.667 ( 0.095, 4.332) | 0.432 ( 0.012, 15.229) | 0.206 ( 0.015, 1.852) | 0.099 ( 0.003, 1.895) | 0.326 ( 0.013, 6.187) | 0.524 ( 0.034, 6.380) |
|  |  |  |  |  | CW_R_BII_B | 0.481 ( 0.011, 17.509) | 0.194 ( 0.011, 3.185) | 0.821 ( 0.018, 39.453) | 0.535 ( 0.059, 4.617) | 0.258 ( 0.004, 10.686) | 0.122 ( 0.001, 8.070) | 0.408 ( 0.004, 29.032) | 0.639 ( 0.009, 40.161) |
|  |  |  |  |  |  | PP_A_BII | 0.406 ( 0.039, 4.396) | 1.699 ( 0.597, 5.631) | 1.123 ( 0.061, 23.194) | 0.528 ( 0.115, 2.081) | 0.248 ( 0.020, 2.609) | 0.845 ( 0.071, 9.767) | 1.327 ( 0.182, 9.863) |
|  |  |  |  |  |  |  | PP_A_BII_B | 4.175 ( 0.306, 62.985) | 2.758 ( 0.466, 17.945) | 1.314 ( 0.071, 17.020) | 0.618 ( 0.019, 17.574) | 2.140 ( 0.061, 55.042) | 3.318 ( 0.144, 69.020) |
|  |  |  |  |  |  |  |  | PP_R_BII | 0.656 ( 0.027, 16.473) | 0.308 ( 0.047, 1.514) | 0.149 ( 0.009, 1.716) | 0.500 ( 0.033, 5.881) | 0.783 ( 0.107, 4.873) |
|  |  |  |  |  |  |  |  |  | PP_R_BII_B | 0.484 ( 0.015, 10.864) | 0.229 ( 0.004, 9.083) | 0.769 ( 0.015, 30.211) | 1.216 ( 0.030, 37.200) |
|  |  |  |  |  |  |  |  |  |  | PR_A_BII | 0.474 ( 0.069, 3.401) | 1.620 ( 0.235, 10.708) | 2.538 ( 0.389, 18.506) |
|  |  |  |  |  |  |  |  |  |  |  | PR_A_BII_B | 3.371 ( 0.499, 25.460) | 5.346 ( 0.374, 90.315) |
|  |  |  |  |  |  |  |  |  |  |  |  | PR_A_RY | 1.582 ( 0.120, 26.143) |
|  |  |  |  |  |  |  |  |  |  |  |  |  | PR_R_BII |

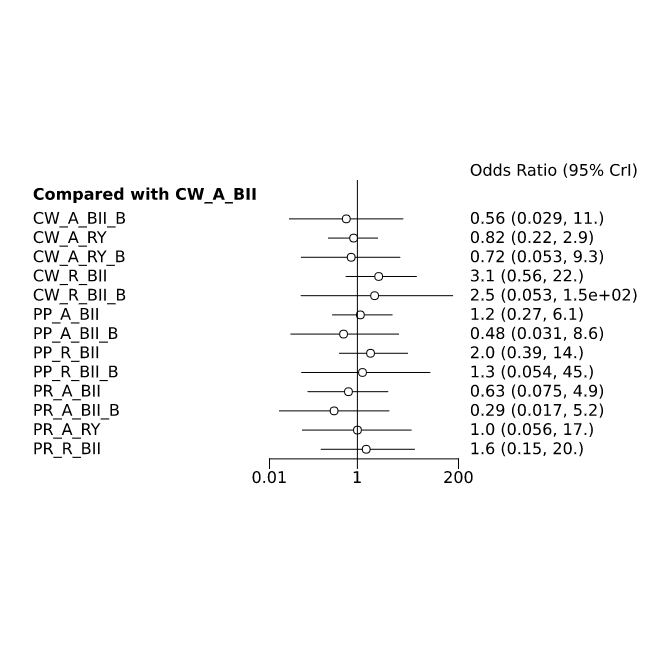

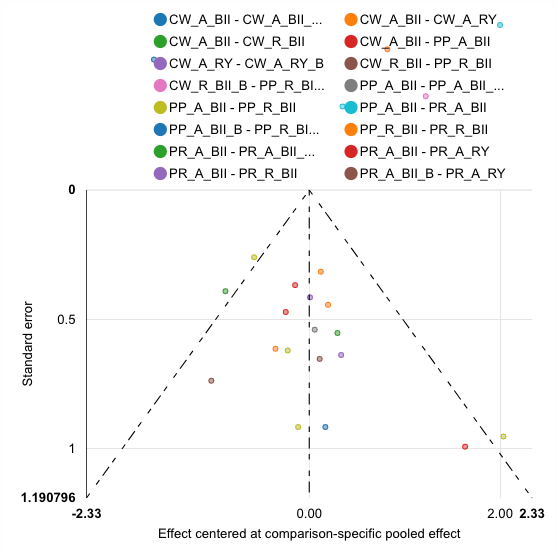

| Rank probabilities table | | | | | | | | | | | | | | |
| --- | --- | --- | --- | --- | --- | --- | --- | --- | --- | --- | --- | --- | --- | --- |
|  | **Rank 1** | **Rank 2** | **Rank 3** | **Rank 4** | **Rank 5** | **Rank 6** | **Rank 7** | **Rank 8** | **Rank 9** | **Rank 10** | **Rank 11** | **Rank 12** | **Rank 13** | **Rank 14** |
| CW_A_BII | 0.005 | 0.019 | 0.053 | 0.081 | 0.109 | 0.125 | 0.124 | 0.126 | 0.116 | 0.096 | 0.080 | 0.043 | 0.019 | 0.003 |
| CW_A_BII_B | 0.215 | 0.120 | 0.079 | 0.075 | 0.069 | 0.061 | 0.048 | 0.042 | 0.043 | 0.046 | 0.046 | 0.049 | 0.050 | 0.059 |
| CW_A_RY | 0.027 | 0.076 | 0.103 | 0.105 | 0.111 | 0.103 | 0.092 | 0.092 | 0.081 | 0.070 | 0.059 | 0.043 | 0.027 | 0.010 |
| CW_A_RY_B | 0.123 | 0.118 | 0.097 | 0.081 | 0.077 | 0.068 | 0.060 | 0.056 | 0.051 | 0.055 | 0.055 | 0.050 | 0.055 | 0.055 |
| CW_R_BII | 0.002 | 0.004 | 0.007 | 0.009 | 0.018 | 0.020 | 0.029 | 0.038 | 0.053 | 0.075 | 0.109 | 0.159 | 0.200 | 0.279 |
| CW_R_BII_B | 0.032 | 0.038 | 0.047 | 0.045 | 0.042 | 0.044 | 0.039 | 0.039 | 0.043 | 0.046 | 0.060 | 0.080 | 0.130 | 0.317 |
| PP_A_BII | 0.001 | 0.006 | 0.013 | 0.037 | 0.066 | 0.107 | 0.154 | 0.170 | 0.165 | 0.137 | 0.083 | 0.043 | 0.016 | 0.003 |
| PP_A_BII_B | 0.170 | 0.162 | 0.137 | 0.110 | 0.086 | 0.070 | 0.061 | 0.049 | 0.046 | 0.036 | 0.033 | 0.032 | 0.007 | 0.003 |
| PP_R_BII | 0.001 | 0.002 | 0.005 | 0.009 | 0.020 | 0.029 | 0.046 | 0.067 | 0.105 | 0.146 | 0.180 | 0.185 | 0.145 | 0.062 |
| PP_R_BII_B | 0.027 | 0.062 | 0.076 | 0.066 | 0.067 | 0.063 | 0.063 | 0.061 | 0.059 | 0.069 | 0.073 | 0.099 | 0.165 | 0.050 |
| PR_A_BII | 0.020 | 0.102 | 0.156 | 0.158 | 0.137 | 0.118 | 0.099 | 0.078 | 0.056 | 0.036 | 0.023 | 0.011 | 0.005 | 0.001 |
| PR_A_BII_B | 0.346 | 0.193 | 0.112 | 0.088 | 0.063 | 0.050 | 0.039 | 0.029 | 0.021 | 0.018 | 0.016 | 0.014 | 0.009 | 0.004 |
| PR_A_RY | 0.022 | 0.076 | 0.081 | 0.087 | 0.081 | 0.076 | 0.078 | 0.075 | 0.074 | 0.075 | 0.072 | 0.074 | 0.067 | 0.061 |
| PR_R_BII | 0.010 | 0.023 | 0.033 | 0.050 | 0.056 | 0.066 | 0.069 | 0.078 | 0.088 | 0.095 | 0.112 | 0.118 | 0.107 | 0.095 |

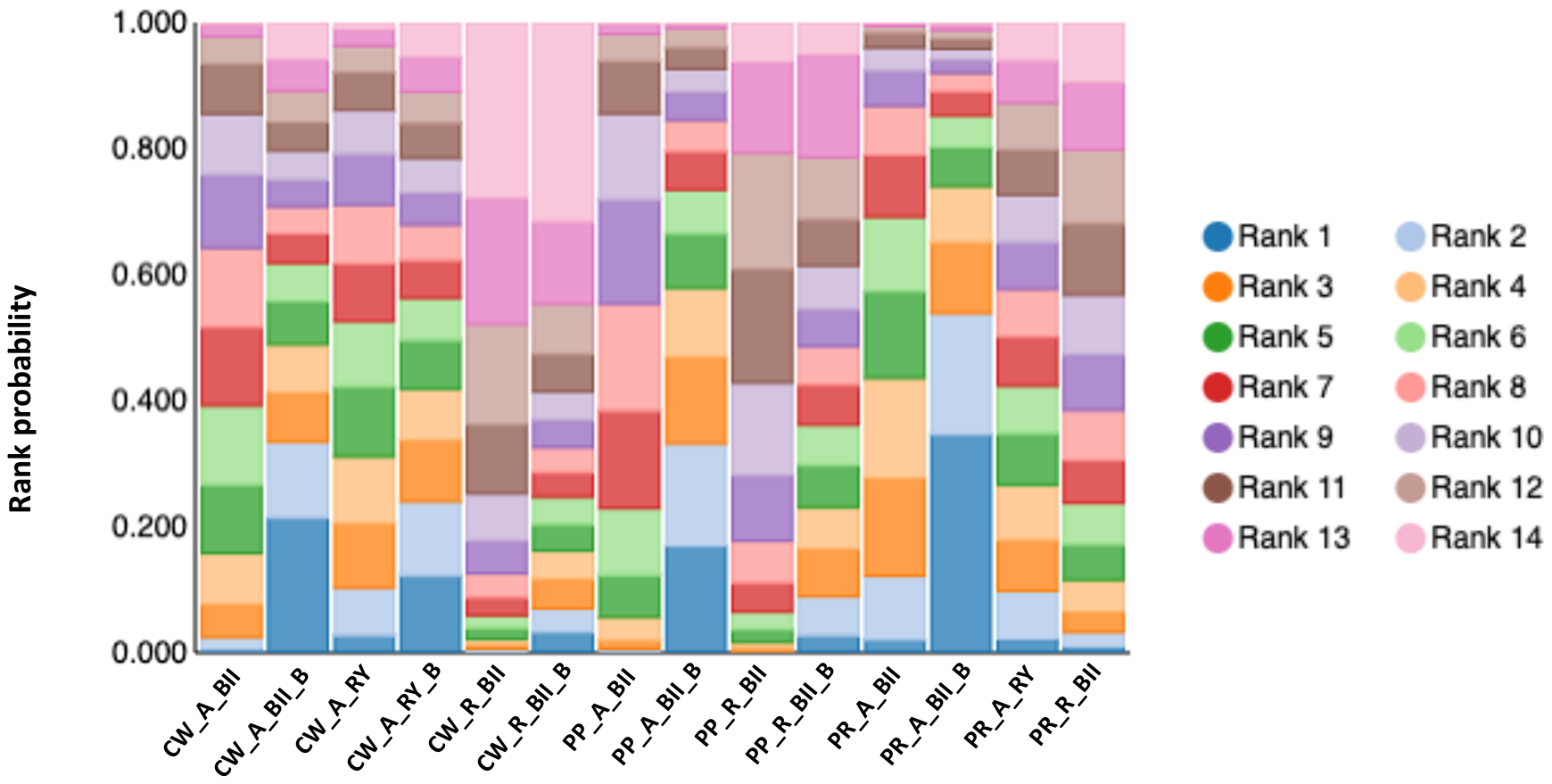

DGE Grade B/C:

Of studies reporting the DGE Grade B/C outcome no direct comparisons existed between studies employing Classic Whipple (CW) and pylorus-resecting (PR)/pylorus-preserving (PP) pancreaticoduodenectomies (PD). Hence, two separate analyses were done to compare the optimal combination approach for clinically significant DGE (ISGPS Grade B/C).

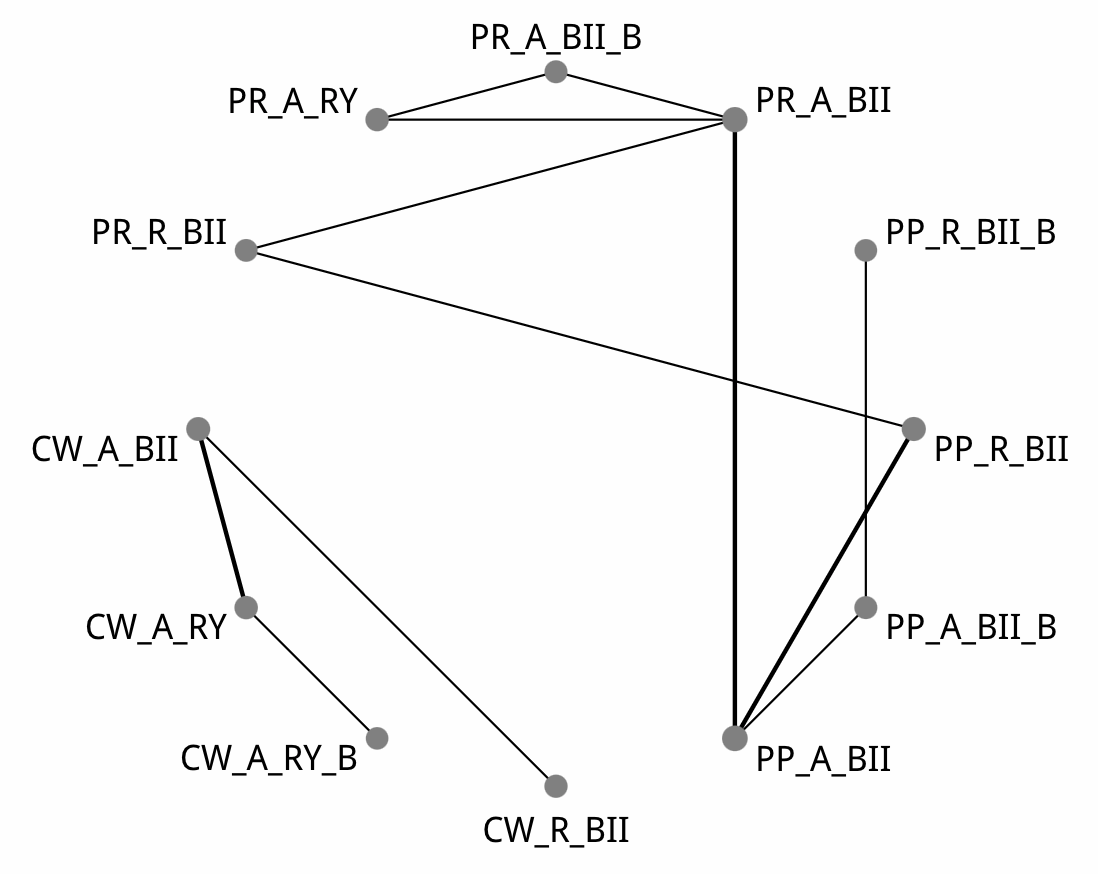

CW DGE Grade B/C:

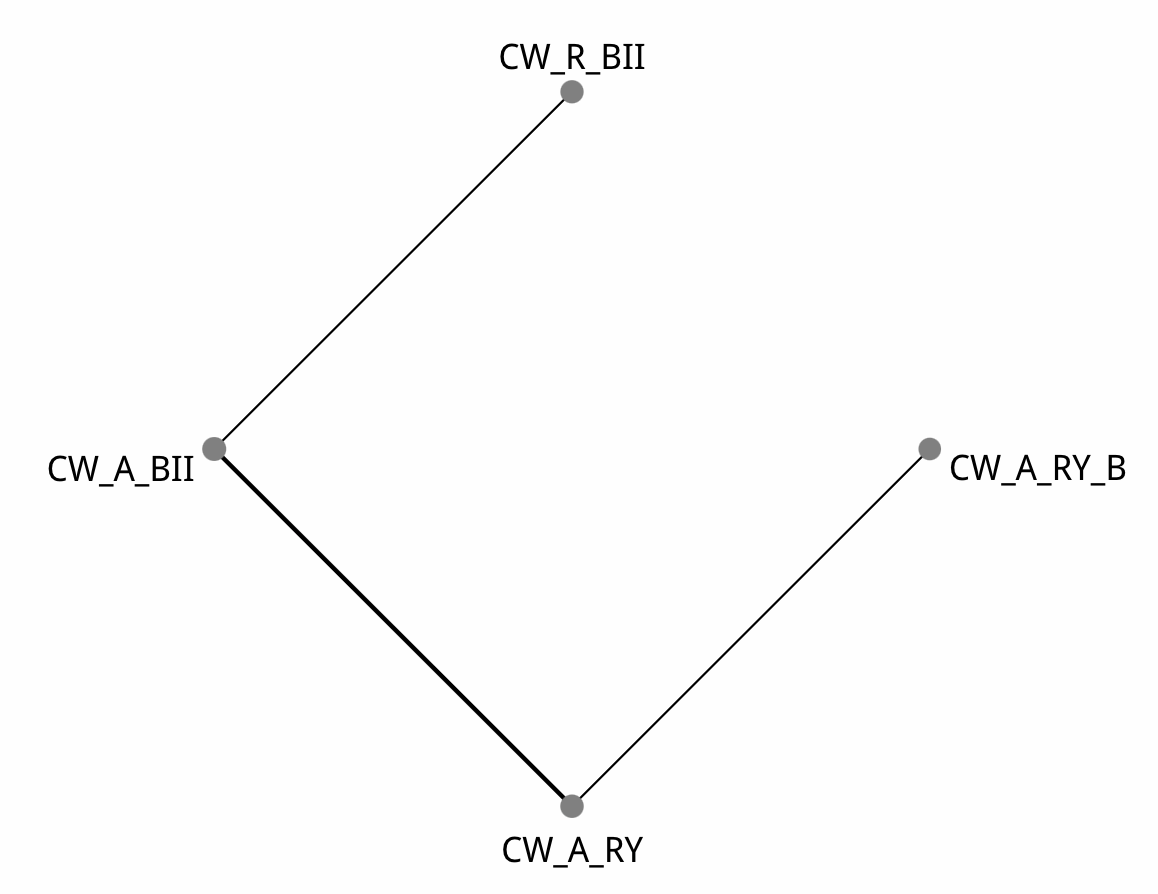

| Comparison of the included interventions: odds ratio (95% CrI). Each cell gives the effect of the column-defining intervention relative to the row-defining intervention. | | | |
| --- | --- | --- | --- |
| CW_A_BII | 1.254 ( 0.458, 3.430) | 1.203 ( 0.197, 6.991) | 1.379 ( 0.479, 4.002) |
|  | CW_A_RY | 0.962 ( 0.222, 3.977) | 1.107 ( 0.259, 4.683) |
|  |  | CW_A_RY_B | 1.147 ( 0.151, 9.251) |
|  |  |  | CW_R_BII |

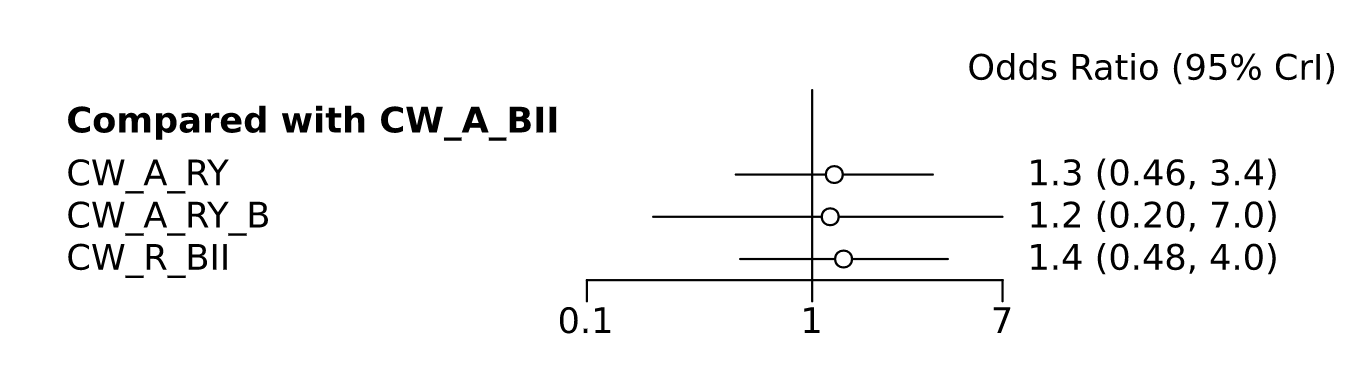

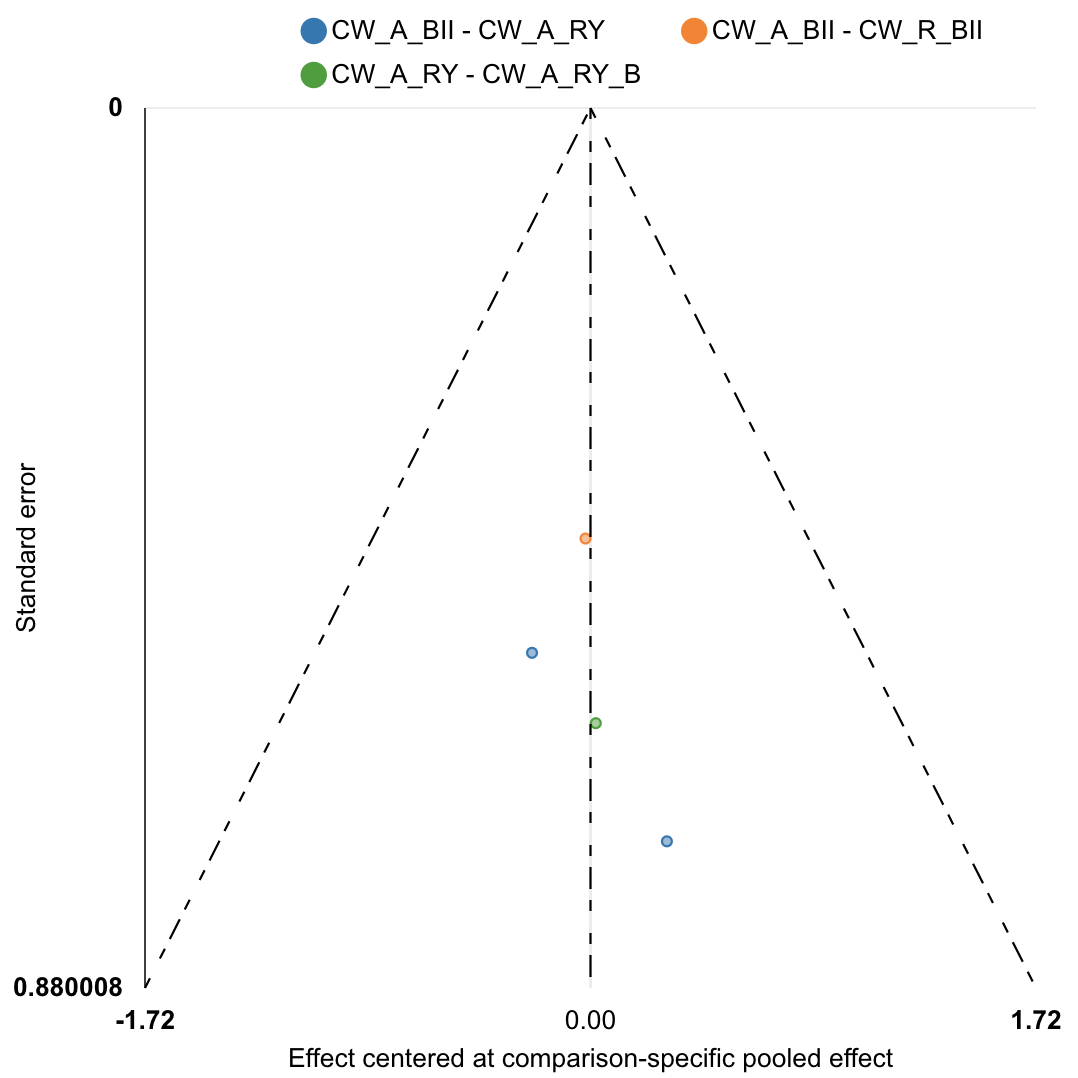

| Rank probabilities table | | | | |
| --- | --- | --- | --- | --- |
|  | **Rank 1** | **Rank 2** | **Rank 3** | **Rank 4** |
| CW_A_BII | 0.351 | 0.352 | 0.230 | 0.066 |
| CW_A_RY | 0.141 | 0.288 | 0.364 | 0.207 |
| CW_A_RY_B | 0.336 | 0.149 | 0.186 | 0.329 |
| CW_R_BII | 0.172 | 0.211 | 0.220 | 0.398 |

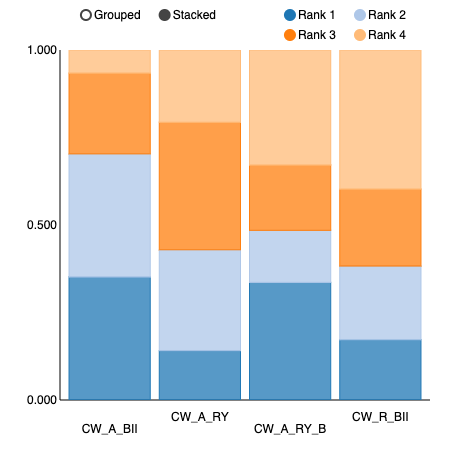

PP and PR DGE Grade B/C:

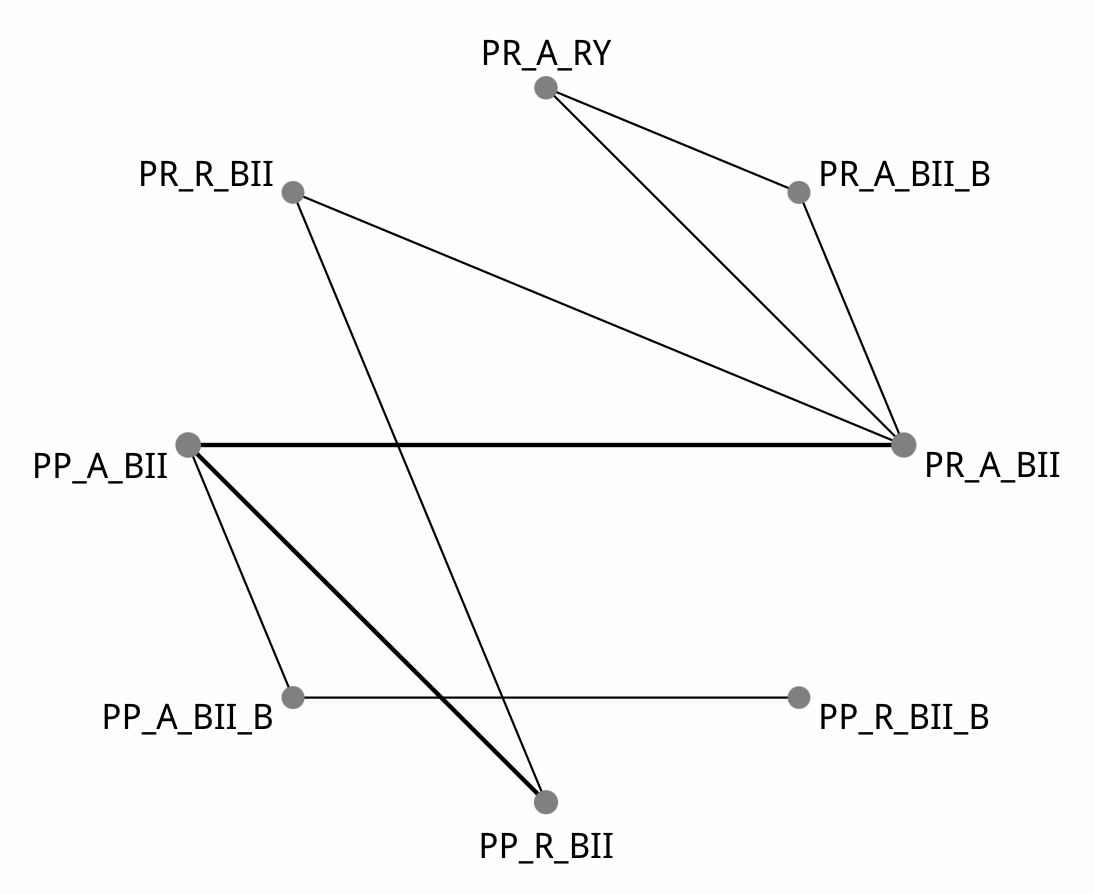

| Comparison of the included interventions: odds ratio (95% CrI). Each cell gives the effect of the column-defining intervention relative to the row-defining intervention. | | | | | | | |
| --- | --- | --- | --- | --- | --- | --- | --- |
| PP_A_BII | 0.077 ( 0.001, 1.653) | 1.679 ( 0.316, 11.579) | 0.163 ( 0.002, 10.993) | 0.648 ( 0.108, 2.955) | 0.427 ( 0.025, 5.534) | 2.107 ( 0.134, 34.073) | 1.610 ( 0.170, 20.035) |
|  | PP_A_BII_B | 22.628 ( 0.632, 1,846.413) | 2.292 ( 0.155, 35.304) | 8.399 ( 0.218, 588.396) | 5.659 ( 0.091, 564.082) | 28.315 ( 0.406, 3,090.228) | 22.776 ( 0.451, 2,027.381) |
|  |  | PP_R_BII | 0.097 ( 0.001, 8.801) | 0.386 ( 0.035, 2.656) | 0.254 ( 0.010, 4.350) | 1.269 ( 0.050, 23.278) | 0.967 ( 0.117, 8.382) |
|  |  |  | PP_R_BII_B | 3.814 ( 0.040, 483.814) | 2.591 ( 0.016, 480.823) | 12.614 ( 0.073, 2,971.137) | 10.453 ( 0.078, 1,650.610) |
|  |  |  |  | PR_A_BII | 0.641 ( 0.083, 5.817) | 3.199 ( 0.377, 32.835) | 2.542 ( 0.304, 33.859) |
|  |  |  |  |  | PR_A_BII_B | 5.070 ( 0.598, 47.499) | 3.797 ( 0.208, 108.701) |
|  |  |  |  |  |  | PR_A_RY | 0.768 ( 0.036, 23.220) |
|  |  |  |  |  |  |  | PR_R_BII |

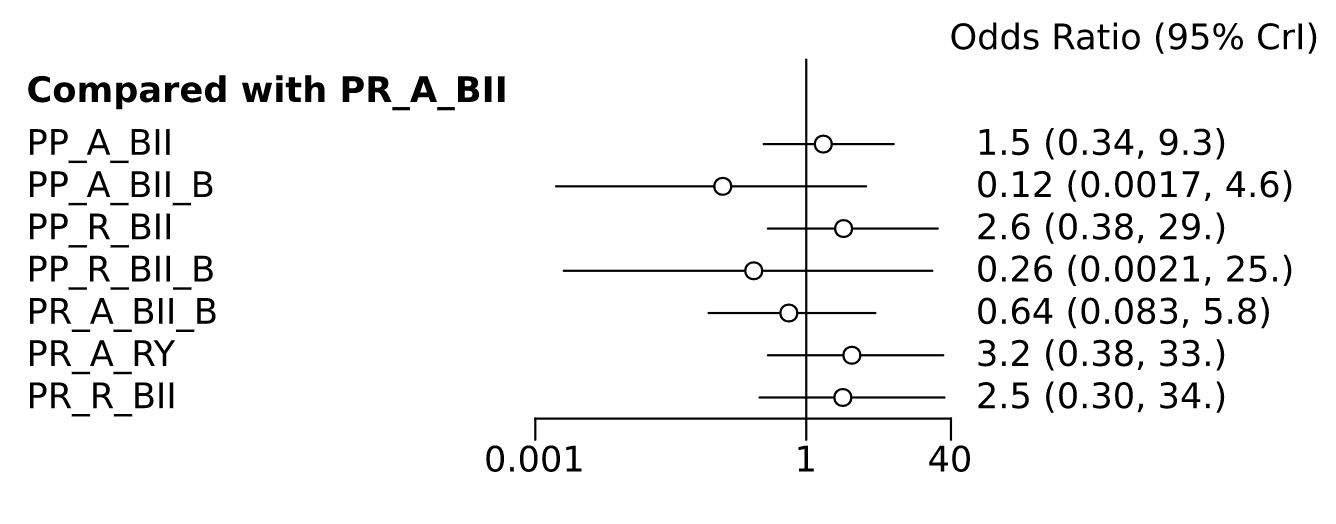

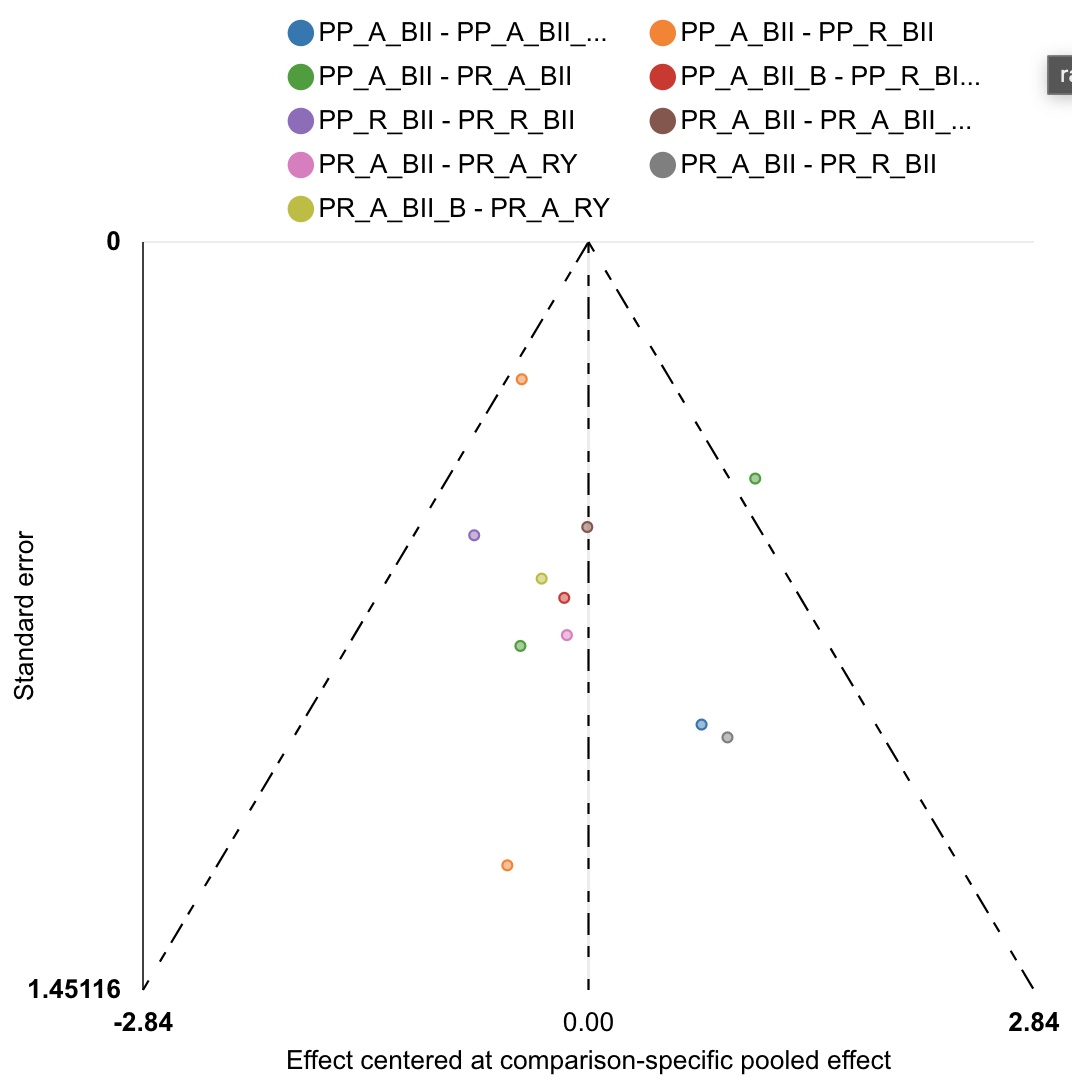

| Rank probabilities table | | | | | | | | |
| --- | --- | --- | --- | --- | --- | --- | --- | --- |
|  | **Rank 1** | **Rank 2** | **Rank 3** | **Rank 4** | **Rank 5** | **Rank 6** | **Rank 7** | **Rank 8** |
| PP_A_BII | 0.004 | 0.022 | 0.119 | 0.185 | 0.278 | 0.230 | 0.123 | 0.039 |
| PP_A_BII_B | 0.580 | 0.271 | 0.061 | 0.035 | 0.024 | 0.014 | 0.013 | 0.003 |
| PP_R_BII | 0.007 | 0.014 | 0.049 | 0.074 | 0.119 | 0.227 | 0.295 | 0.216 |
| PP_R_BII_B | 0.217 | 0.413 | 0.099 | 0.058 | 0.054 | 0.043 | 0.049 | 0.069 |
| PR_A_BII | 0.025 | 0.093 | 0.221 | 0.330 | 0.194 | 0.097 | 0.036 | 0.004 |
| PR_A_BII_B | 0.147 | 0.145 | 0.335 | 0.140 | 0.094 | 0.067 | 0.061 | 0.013 |
| PR_A_RY | 0.007 | 0.019 | 0.046 | 0.084 | 0.110 | 0.145 | 0.170 | 0.419 |
| PR_R_BII | 0.014 | 0.024 | 0.071 | 0.095 | 0.129 | 0.177 | 0.253 | 0.238 |

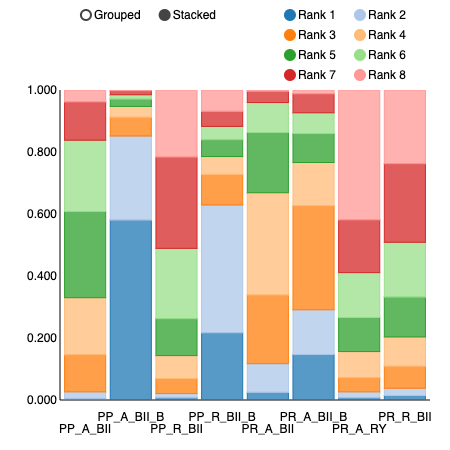

POPF Overall

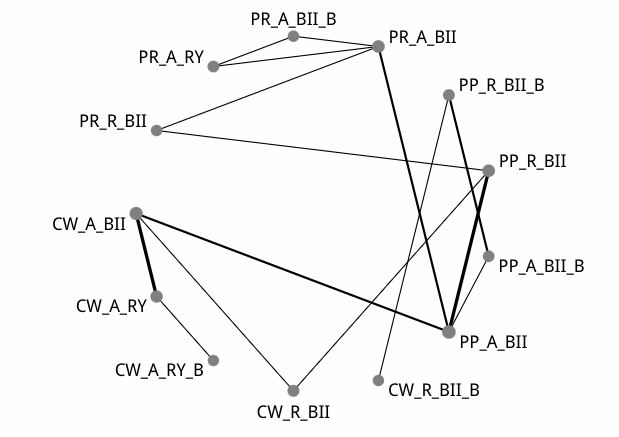

| Comparison of the included interventions: odds ratio (95% CrI). Each cell gives the effect of the column-defining intervention relative to the row-defining intervention. | | | | | | | | | | | | |
| --- | --- | --- | --- | --- | --- | --- | --- | --- | --- | --- | --- | --- |
| CW.A.BII | 0.991 ( 0.517, 1.925) | 0.939 ( 0.284, 3.285) | 0.934 ( 0.420, 2.034) | 0.058 ( 0.001, 1.192) | 0.655 ( 0.267, 1.585) | 0.336 ( 0.066, 1.783) | 0.696 ( 0.249, 1.766) | 0.232 ( 0.034, 1.564) | 0.432 ( 0.147, 1.269) | 0.513 ( 0.114, 2.162) | 0.326 ( 0.077, 1.220) | 0.744 ( 0.190, 2.861) |
|  | CW.A.RY | 0.950 ( 0.338, 2.721) | 0.946 ( 0.327, 2.580) | 0.058 ( 0.001, 1.249) | 0.658 ( 0.208, 1.943) | 0.339 ( 0.056, 1.969) | 0.689 ( 0.205, 2.167) | 0.228 ( 0.030, 1.801) | 0.436 ( 0.124, 1.527) | 0.516 ( 0.098, 2.540) | 0.325 ( 0.067, 1.416) | 0.742 ( 0.174, 3.208) |
|  |  | CW.A.RY.B | 0.990 ( 0.231, 4.121) | 0.060 ( 0.001, 1.641) | 0.704 ( 0.145, 3.084) | 0.360 ( 0.045, 2.754) | 0.737 ( 0.143, 3.455) | 0.245 ( 0.024, 2.380) | 0.465 ( 0.085, 2.268) | 0.559 ( 0.074, 3.616) | 0.352 ( 0.048, 2.048) | 0.781 ( 0.125, 5.089) |
|  |  |  | CW.R.BII | 0.063 ( 0.001, 1.308) | 0.707 ( 0.260, 1.879) | 0.368 ( 0.065, 1.955) | 0.741 ( 0.257, 1.988) | 0.250 ( 0.034, 1.759) | 0.465 ( 0.142, 1.497) | 0.548 ( 0.117, 2.623) | 0.350 ( 0.079, 1.425) | 0.796 ( 0.203, 3.110) |
|  |  |  |  | CW.R.BII.B | 11.081 ( 0.621, 487.310) | 5.759 ( 0.469, 193.892) | 11.824 ( 0.600, 524.896) | 3.953 ( 0.401, 136.115) | 7.377 ( 0.367, 346.367) | 9.142 ( 0.372, 490.635) | 5.659 ( 0.242, 285.203) | 12.527 ( 0.572, 626.783) |
|  |  |  |  |  | PP.A.BII | 0.520 ( 0.126, 2.069) | 1.054 ( 0.539, 1.962) | 0.356 ( 0.064, 1.949) | 0.666 ( 0.343, 1.253) | 0.795 ( 0.232, 2.533) | 0.500 ( 0.161, 1.361) | 1.125 ( 0.380, 3.416) |
|  |  |  |  |  |  | PP.A.BII.B | 2.017 ( 0.439, 9.361) | 0.694 ( 0.265, 1.784) | 1.266 ( 0.275, 6.274) | 1.509 ( 0.235, 9.519) | 0.943 ( 0.154, 5.518) | 2.140 ( 0.391, 12.528) |
|  |  |  |  |  |  |  | PP.R.BII | 0.339 ( 0.056, 2.133) | 0.631 ( 0.268, 1.528) | 0.750 ( 0.199, 2.828) | 0.470 ( 0.137, 1.515) | 1.070 ( 0.400, 3.074) |
|  |  |  |  |  |  |  |  | PP.R.BII.B | 1.832 ( 0.300, 11.934) | 2.218 ( 0.261, 16.511) | 1.350 ( 0.165, 10.198) | 3.105 ( 0.415, 23.677) |
|  |  |  |  |  |  |  |  |  | PR.A.BII | 1.191 ( 0.429, 3.188) | 0.746 ( 0.306, 1.712) | 1.711 ( 0.553, 5.442) |
|  |  |  |  |  |  |  |  |  |  | PR.A.BII.B | 0.630 ( 0.231, 1.650) | 1.429 ( 0.331, 6.646) |
|  |  |  |  |  |  |  |  |  |  |  | PR.A.RY | 2.294 ( 0.536, 9.467) |
|  |  |  |  |  |  |  |  |  |  |  |  | PR.R.BII |

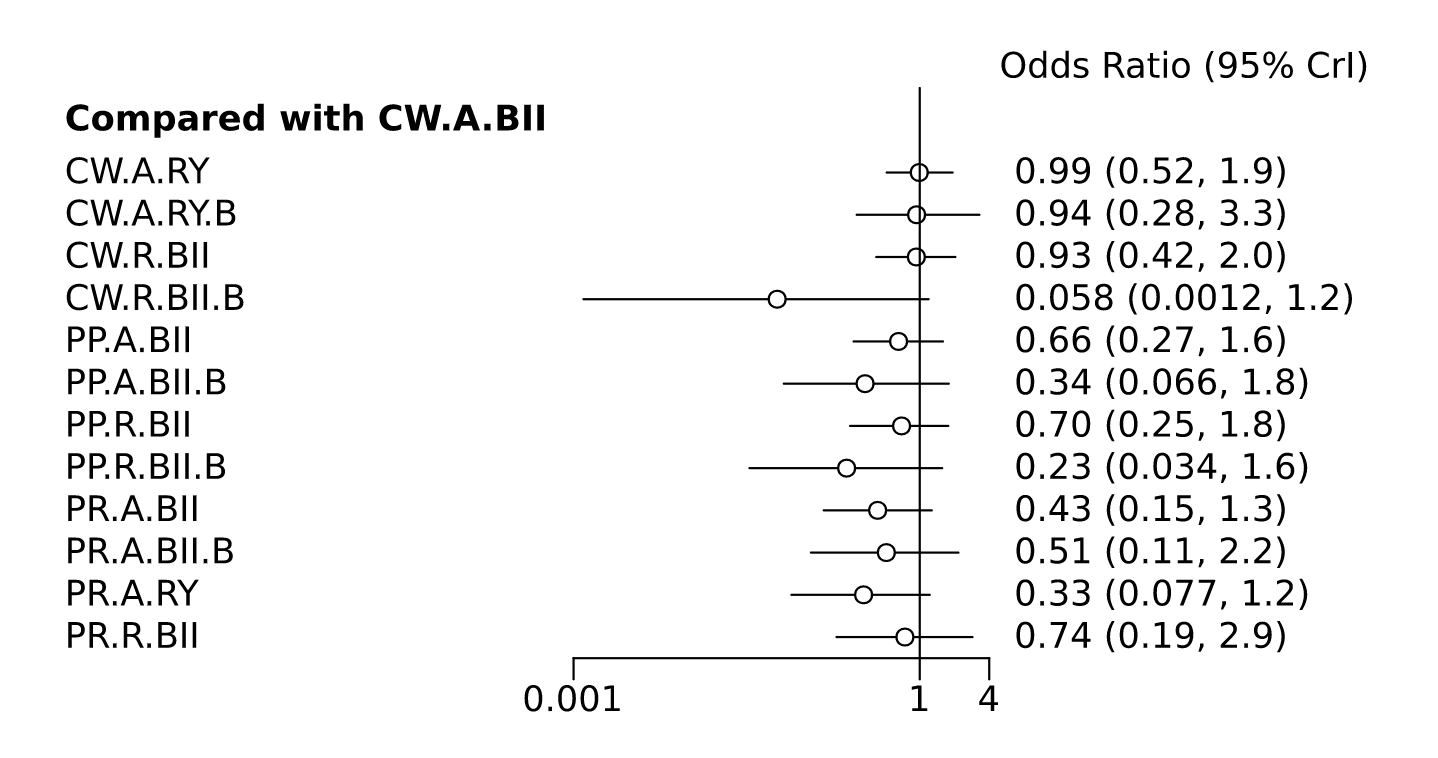

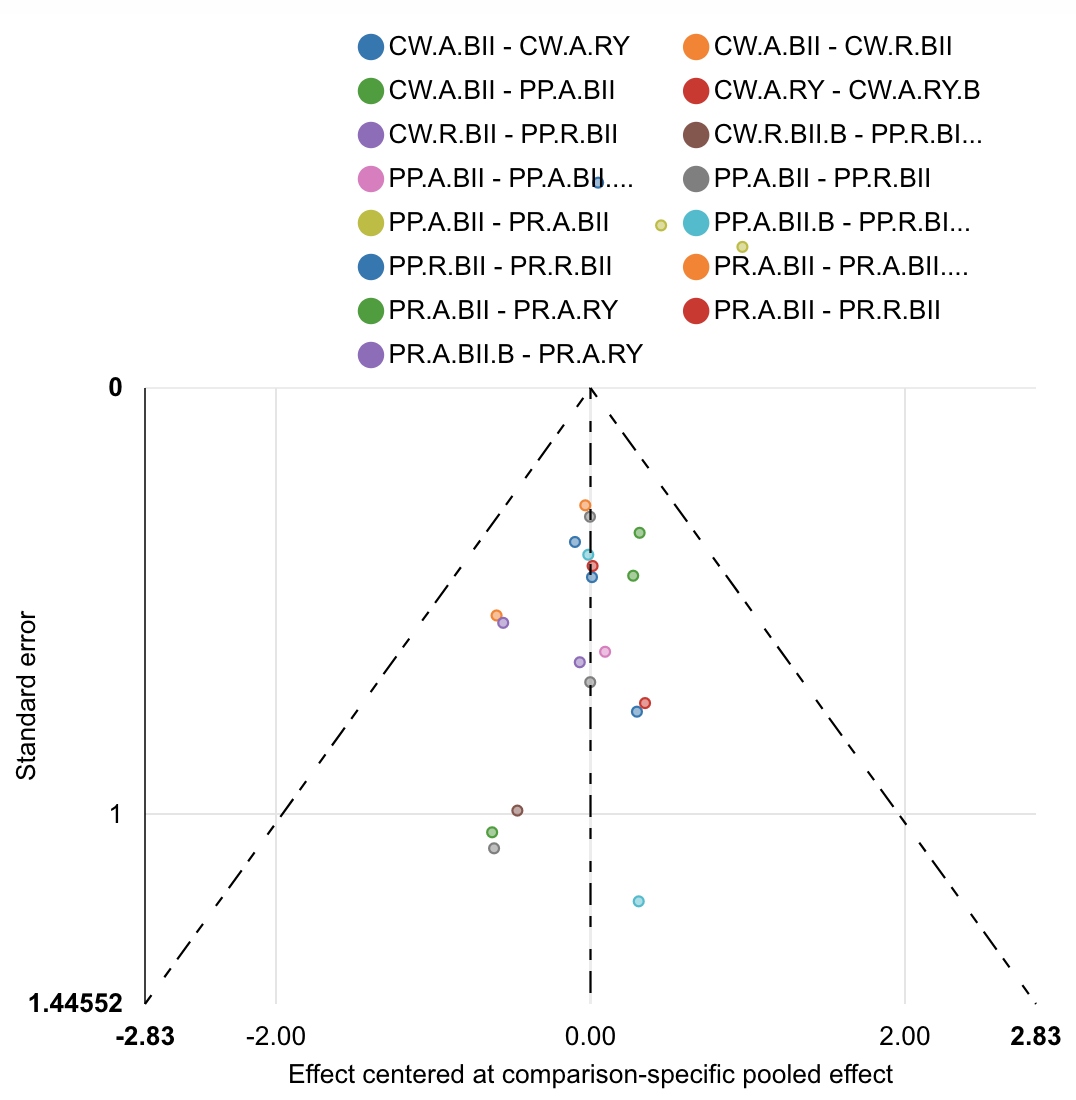

| Rank probabilities table | | | | | | | | | | | | | |
| --- | --- | --- | --- | --- | --- | --- | --- | --- | --- | --- | --- | --- | --- |
|  | **Rank 1** | **Rank 2** | **Rank 3** | **Rank 4** | **Rank 5** | **Rank 6** | **Rank 7** | **Rank 8** | **Rank 9** | **Rank 10** | **Rank 11** | **Rank 12** | **Rank 13** |
| CW.A.BII | 0.001 | 0.001 | 0.003 | 0.007 | 0.016 | 0.025 | 0.038 | 0.057 | 0.099 | 0.179 | 0.242 | 0.212 | 0.122 |
| CW.A.RY | 0.001 | 0.004 | 0.011 | 0.020 | 0.025 | 0.038 | 0.050 | 0.067 | 0.086 | 0.133 | 0.194 | 0.229 | 0.142 |
| CW.A.RY.B | 0.010 | 0.024 | 0.033 | 0.046 | 0.047 | 0.053 | 0.058 | 0.067 | 0.076 | 0.104 | 0.088 | 0.118 | 0.276 |
| CW.R.BII | 0.002 | 0.006 | 0.011 | 0.023 | 0.033 | 0.041 | 0.060 | 0.078 | 0.107 | 0.179 | 0.159 | 0.149 | 0.153 |
| CW.R.BII.B | 0.786 | 0.074 | 0.039 | 0.022 | 0.012 | 0.013 | 0.009 | 0.007 | 0.006 | 0.007 | 0.007 | 0.006 | 0.013 |
| PP.A.BII | 0.000 | 0.001 | 0.004 | 0.021 | 0.053 | 0.119 | 0.244 | 0.216 | 0.158 | 0.090 | 0.051 | 0.032 | 0.011 |
| PP.A.BII.B | 0.014 | 0.092 | 0.338 | 0.139 | 0.098 | 0.089 | 0.052 | 0.045 | 0.043 | 0.022 | 0.022 | 0.021 | 0.025 |
| PP.R.BII | 0.002 | 0.006 | 0.015 | 0.037 | 0.062 | 0.098 | 0.141 | 0.193 | 0.173 | 0.109 | 0.074 | 0.061 | 0.029 |
| PP.R.BII.B | 0.074 | 0.465 | 0.144 | 0.079 | 0.060 | 0.043 | 0.031 | 0.022 | 0.016 | 0.018 | 0.016 | 0.021 | 0.012 |
| PR.A.BII | 0.008 | 0.041 | 0.102 | 0.188 | 0.286 | 0.185 | 0.082 | 0.046 | 0.027 | 0.018 | 0.010 | 0.005 | 0.002 |
| PR.A.BII.B | 0.015 | 0.059 | 0.089 | 0.110 | 0.128 | 0.152 | 0.101 | 0.075 | 0.069 | 0.047 | 0.050 | 0.048 | 0.058 |
| PR.A.RY | 0.081 | 0.208 | 0.175 | 0.240 | 0.115 | 0.063 | 0.034 | 0.028 | 0.025 | 0.011 | 0.009 | 0.008 | 0.004 |
| PR.R.BII | 0.006 | 0.018 | 0.035 | 0.067 | 0.067 | 0.082 | 0.101 | 0.098 | 0.117 | 0.084 | 0.079 | 0.092 | 0.154 |

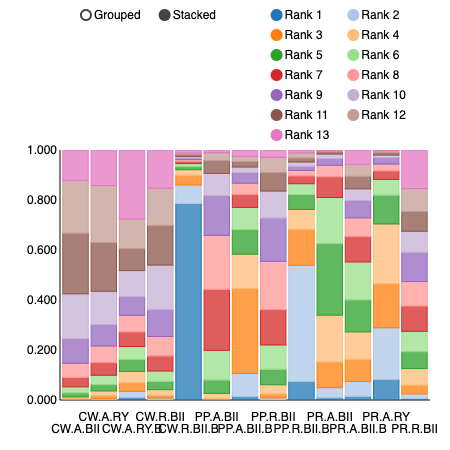

POPF Overall sensitivity analysis with removal of Seiler et al. 2005 (single study with comparison of CW_R_BII_B).

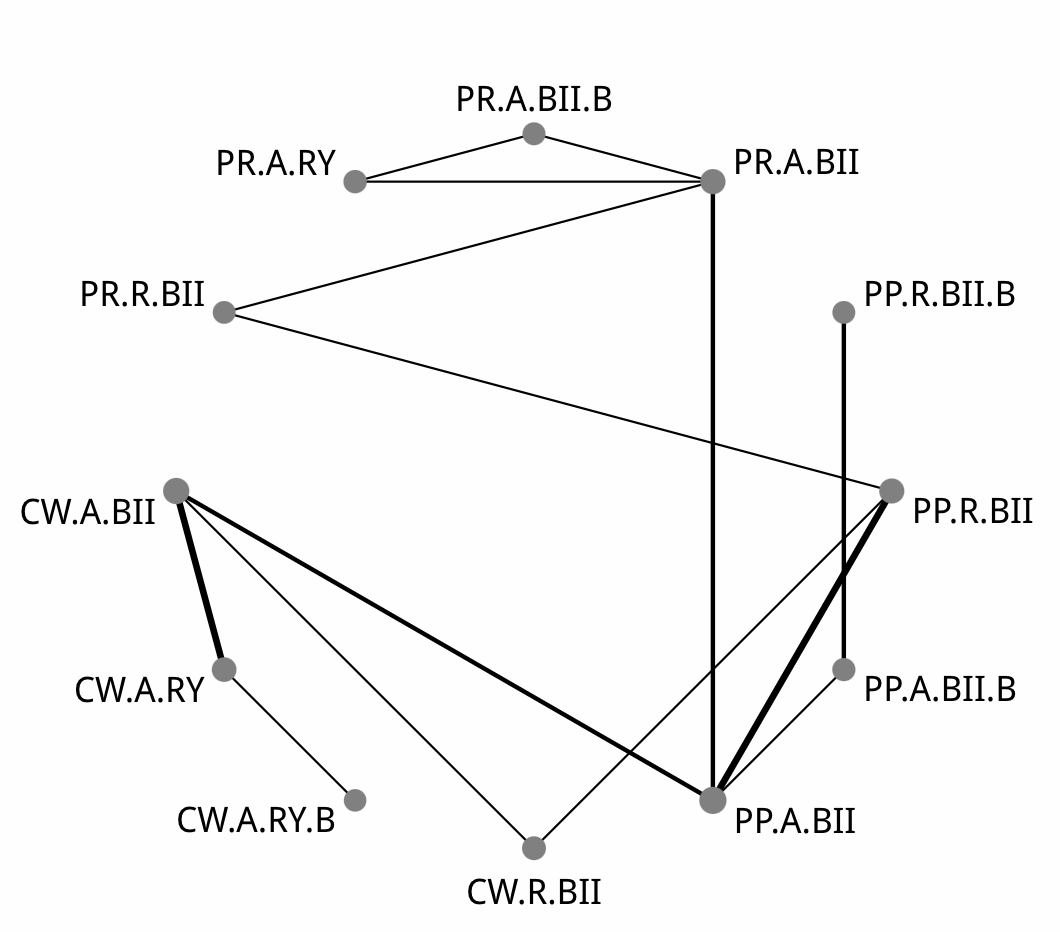

| Comparison of the included interventions: odds ratio (95% CrI). Each cell gives the effect of the column-defining intervention relative to the row-defining intervention. | | | | | | | | | | | |
| --- | --- | --- | --- | --- | --- | --- | --- | --- | --- | --- | --- |
| CW.A.BII | 0.981 ( 0.515, 1.944) | 0.923 ( 0.265, 3.253) | 0.934 ( 0.433, 2.023) | 0.658 ( 0.262, 1.569) | 0.350 ( 0.061, 1.760) | 0.684 ( 0.246, 1.833) | 0.243 ( 0.033, 1.536) | 0.443 ( 0.140, 1.252) | 0.530 ( 0.117, 2.139) | 0.337 ( 0.074, 1.178) | 0.761 ( 0.197, 2.859) |
|  | CW.A.RY | 0.931 ( 0.325, 2.691) | 0.942 ( 0.332, 2.562) | 0.675 ( 0.213, 1.916) | 0.359 ( 0.056, 1.947) | 0.694 ( 0.199, 2.186) | 0.248 ( 0.031, 1.699) | 0.454 ( 0.116, 1.504) | 0.536 ( 0.102, 2.458) | 0.338 ( 0.063, 1.346) | 0.775 ( 0.171, 3.336) |
|  |  | CW.A.RY.B | 1.007 ( 0.241, 4.371) | 0.724 ( 0.158, 3.139) | 0.385 ( 0.048, 2.797) | 0.754 ( 0.148, 3.397) | 0.273 ( 0.026, 2.306) | 0.486 ( 0.088, 2.368) | 0.580 ( 0.083, 3.914) | 0.364 ( 0.051, 2.067) | 0.829 ( 0.136, 4.826) |
|  |  |  | CW.R.BII | 0.707 ( 0.250, 1.939) | 0.371 ( 0.060, 2.034) | 0.740 ( 0.245, 1.990) | 0.262 ( 0.035, 1.761) | 0.475 ( 0.135, 1.483) | 0.576 ( 0.114, 2.428) | 0.360 ( 0.072, 1.404) | 0.810 ( 0.192, 3.194) |
|  |  |  |  | PP.A.BII | 0.525 ( 0.123, 2.126) | 1.038 ( 0.540, 1.953) | 0.367 ( 0.065, 1.928) | 0.673 ( 0.344, 1.273) | 0.804 ( 0.241, 2.571) | 0.503 ( 0.164, 1.383) | 1.134 ( 0.400, 3.418) |
|  |  |  |  |  | PP.A.BII.B | 1.974 ( 0.436, 9.530) | 0.695 ( 0.269, 1.765) | 1.263 ( 0.262, 6.190) | 1.472 ( 0.247, 9.474) | 0.944 ( 0.163, 5.659) | 2.180 ( 0.373, 13.410) |
|  |  |  |  |  |  | PP.R.BII | 0.349 ( 0.058, 2.104) | 0.644 ( 0.270, 1.512) | 0.769 ( 0.210, 2.854) | 0.486 ( 0.142, 1.541) | 1.109 ( 0.420, 3.071) |
|  |  |  |  |  |  |  | PP.R.BII.B | 1.840 ( 0.293, 11.146) | 2.096 ( 0.285, 18.256) | 1.352 ( 0.181, 10.171) | 3.175 ( 0.429, 23.850) |
|  |  |  |  |  |  |  |  | PR.A.BII | 1.170 ( 0.459, 3.162) | 0.753 ( 0.315, 1.689) | 1.712 ( 0.576, 5.337) |
|  |  |  |  |  |  |  |  |  | PR.A.BII.B | 0.645 ( 0.220, 1.617) | 1.455 ( 0.332, 6.311) |
|  |  |  |  |  |  |  |  |  |  | PR.A.RY | 2.292 ( 0.572, 9.683) |
|  |  |  |  |  |  |  |  |  |  |  | PR.R.BII |

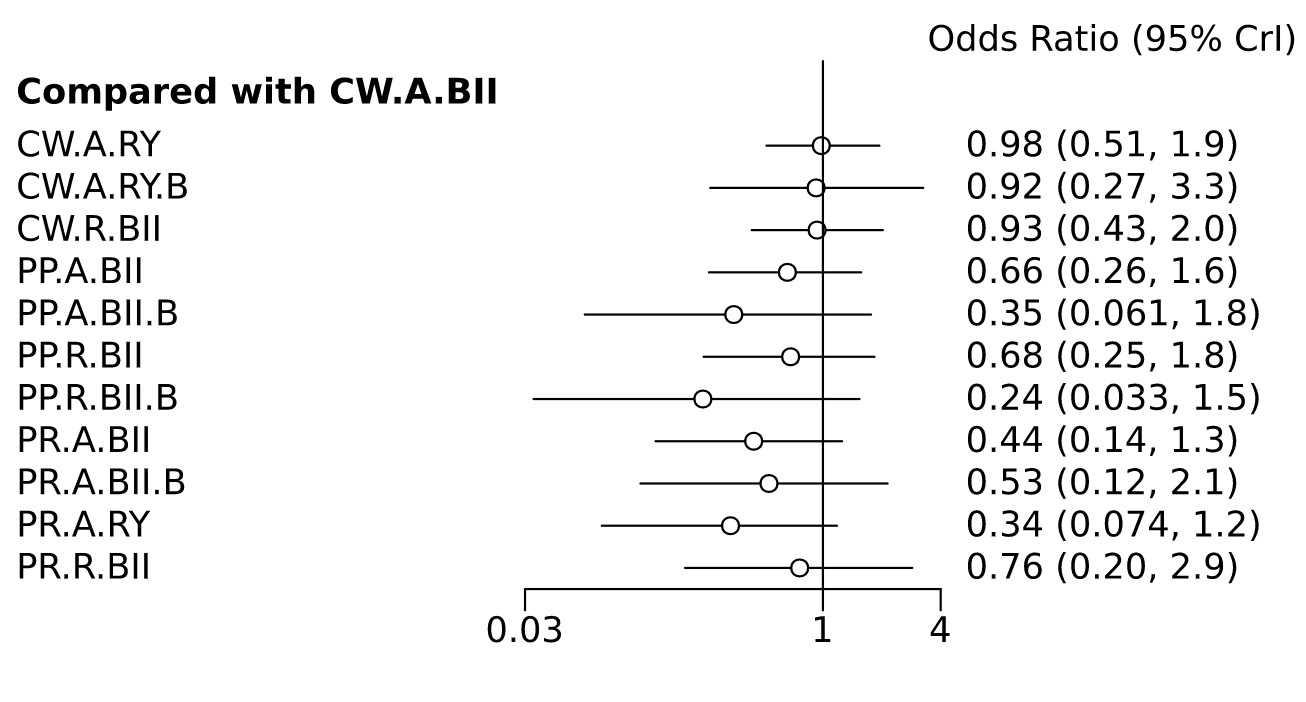

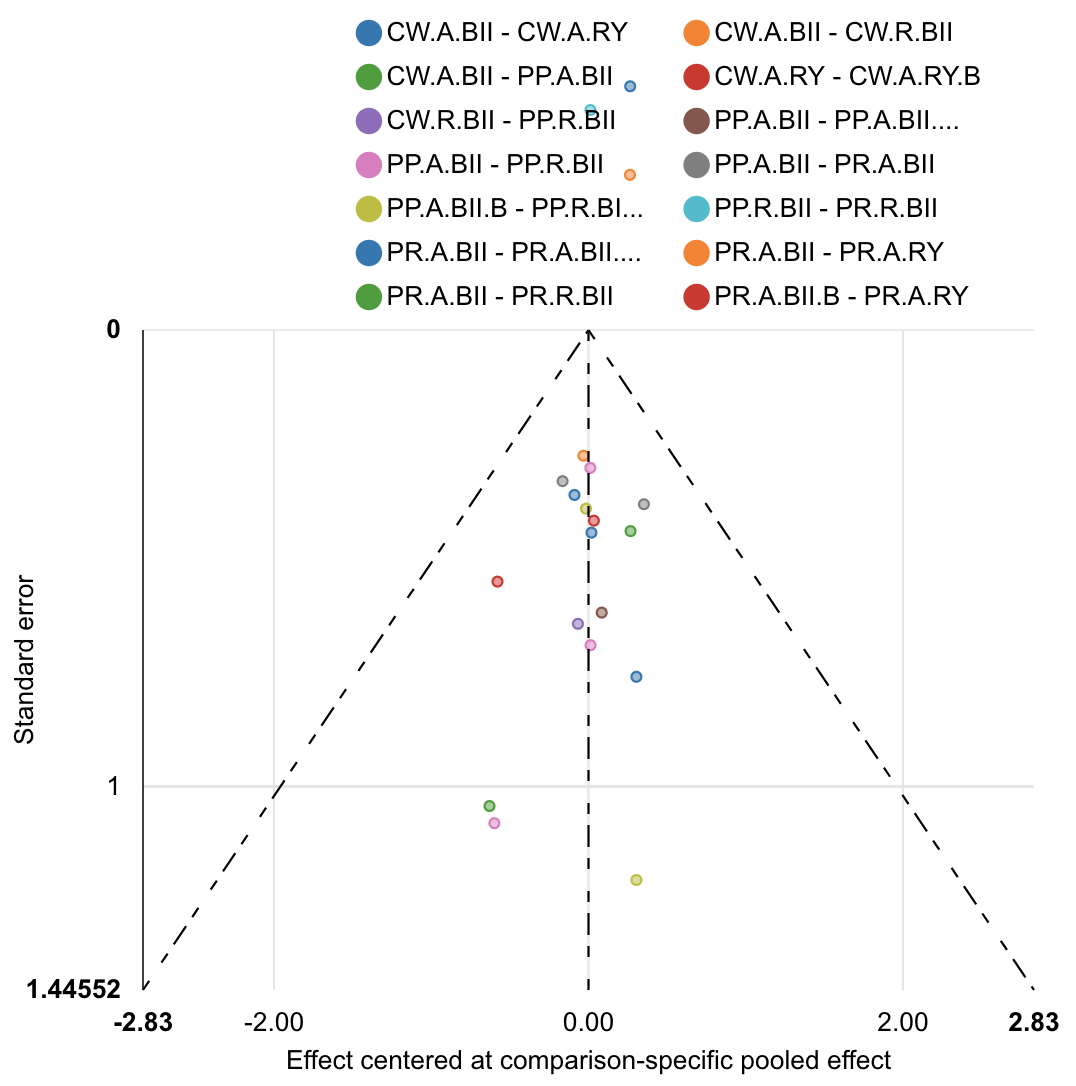

| Rank probabilities table | | | | | | | | | | | | |
| --- | --- | --- | --- | --- | --- | --- | --- | --- | --- | --- | --- | --- |
|  | **Rank 1** | **Rank 2** | **Rank 3** | **Rank 4** | **Rank 5** | **Rank 6** | **Rank 7** | **Rank 8** | **Rank 9** | **Rank 10** | **Rank 11** | **Rank 12** |
| CW.A.BII | 0.001 | 0.003 | 0.006 | 0.014 | 0.028 | 0.038 | 0.057 | 0.088 | 0.167 | 0.248 | 0.233 | 0.118 |
| CW.A.RY | 0.004 | 0.009 | 0.020 | 0.029 | 0.039 | 0.050 | 0.065 | 0.087 | 0.138 | 0.196 | 0.216 | 0.149 |
| CW.A.RY.B | 0.026 | 0.031 | 0.053 | 0.052 | 0.053 | 0.068 | 0.063 | 0.074 | 0.104 | 0.100 | 0.126 | 0.250 |
| CW.R.BII | 0.005 | 0.011 | 0.022 | 0.031 | 0.042 | 0.058 | 0.080 | 0.115 | 0.176 | 0.152 | 0.142 | 0.166 |
| PP.A.BII | 0.001 | 0.003 | 0.024 | 0.057 | 0.120 | 0.218 | 0.211 | 0.171 | 0.097 | 0.055 | 0.032 | 0.012 |
| PP.A.BII.B | 0.092 | 0.344 | 0.141 | 0.092 | 0.087 | 0.061 | 0.048 | 0.039 | 0.026 | 0.023 | 0.025 | 0.024 |
| PP.R.BII | 0.005 | 0.013 | 0.040 | 0.074 | 0.104 | 0.149 | 0.189 | 0.155 | 0.103 | 0.078 | 0.063 | 0.030 |
| PP.R.BII.B | 0.510 | 0.166 | 0.077 | 0.062 | 0.040 | 0.033 | 0.024 | 0.024 | 0.018 | 0.013 | 0.016 | 0.016 |
| PR.A.BII | 0.031 | 0.108 | 0.189 | 0.271 | 0.209 | 0.083 | 0.047 | 0.030 | 0.016 | 0.010 | 0.005 | 0.001 |
| PR.A.BII.B | 0.047 | 0.092 | 0.127 | 0.139 | 0.134 | 0.097 | 0.084 | 0.075 | 0.057 | 0.041 | 0.050 | 0.058 |
| PR.A.RY | 0.257 | 0.194 | 0.252 | 0.117 | 0.066 | 0.036 | 0.028 | 0.021 | 0.012 | 0.010 | 0.007 | 0.002 |
| PR.R.BII | 0.020 | 0.029 | 0.051 | 0.063 | 0.078 | 0.111 | 0.104 | 0.123 | 0.086 | 0.074 | 0.087 | 0.174 |

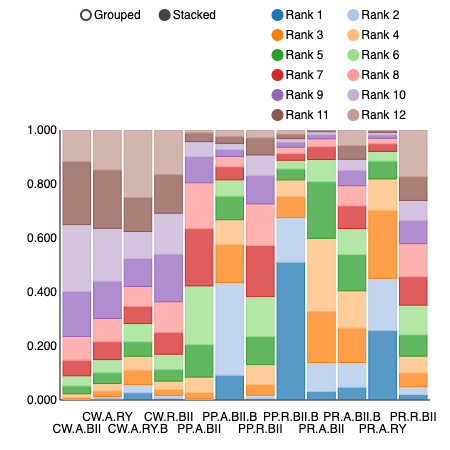

POPF Grade B/C

Of studies reporting the POPF Grade B/C outcome no direct comparisons existed between studies employing Classic Whipple (CW) and pylorus-resecting (PR)/pylorus-preserving (PP) pancreaticoduodenectomies (PD). Hence, two separate analyses were done to compare the optimal combination approach for clinically significant DGE (ISGPF Grade B/C).

CW POPF Grade B/C

| Comparison of the included interventions: odds ratio (95% CrI). Each cell gives the effect of the column-defining intervention relative to the row-defining intervention. | | | |
| --- | --- | --- | --- |
| CW_A_BII | 0.629 ( 0.248, 1.663) | 0.740 ( 0.098, 5.843) | 0.766 ( 0.211, 2.794) |
|  | CW_A_RY | 1.172 ( 0.190, 7.388) | 1.232 ( 0.236, 5.926) |
|  |  | CW_A_RY_B | 1.053 ( 0.092, 11.175) |
|  |  |  | CW_R_BII |

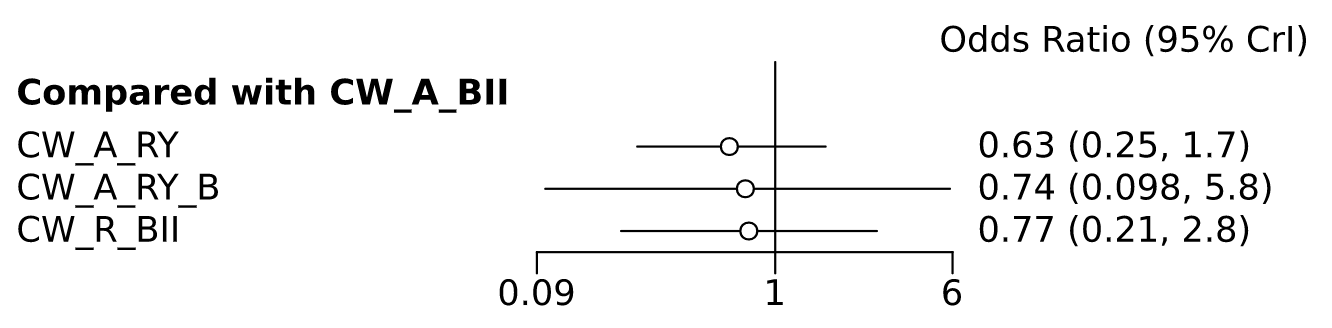

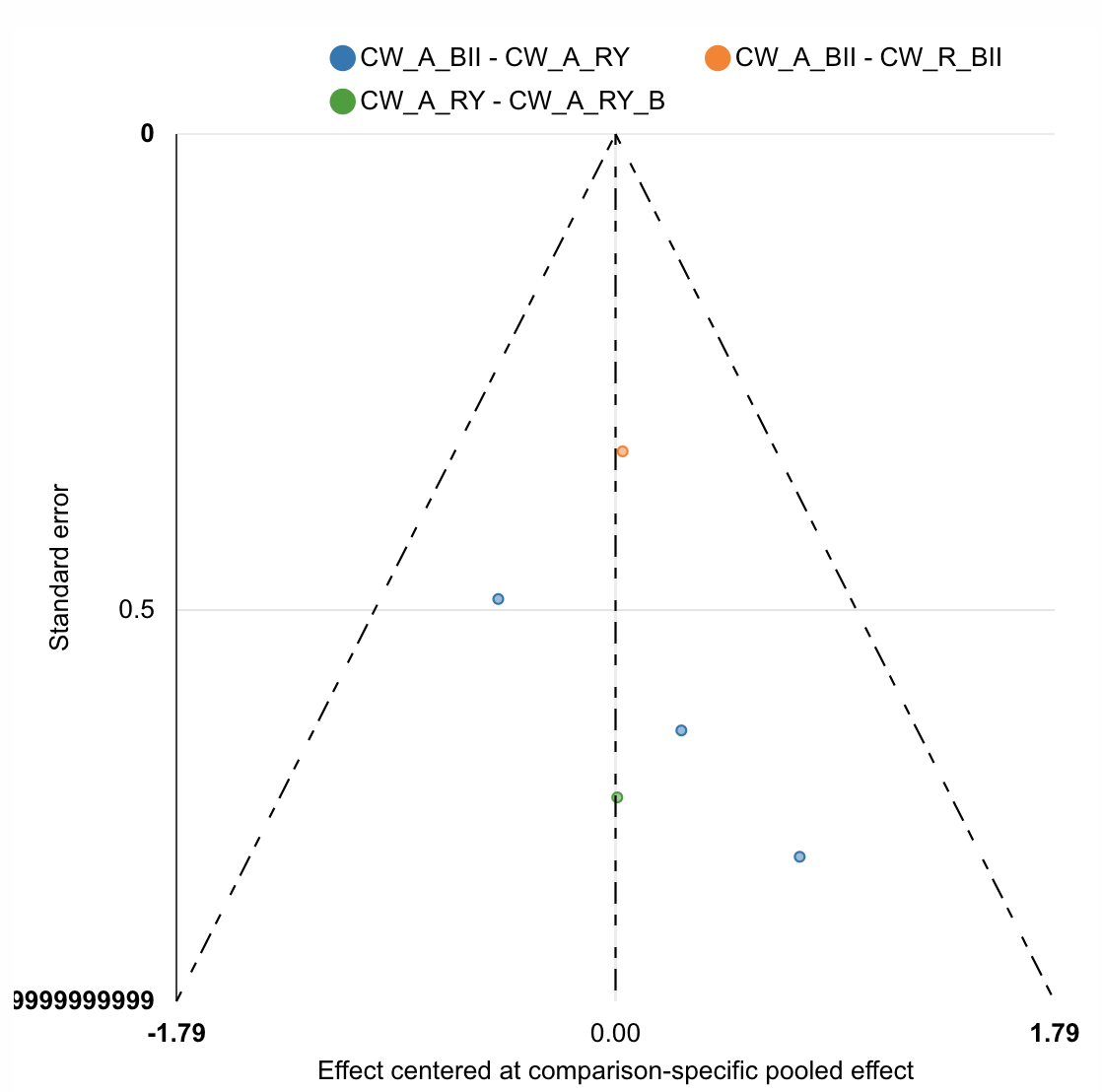

| Rank probabilities table | | | | |
| --- | --- | --- | --- | --- |
|  | **Rank 1** | **Rank 2** | **Rank 3** | **Rank 4** |
| CW_A_BII | 0.036 | 0.166 | 0.406 | 0.393 |
| CW_A_RY | 0.331 | 0.424 | 0.192 | 0.054 |
| CW_A_RY_B | 0.355 | 0.185 | 0.133 | 0.327 |
| CW_R_BII | 0.279 | 0.225 | 0.270 | 0.227 |

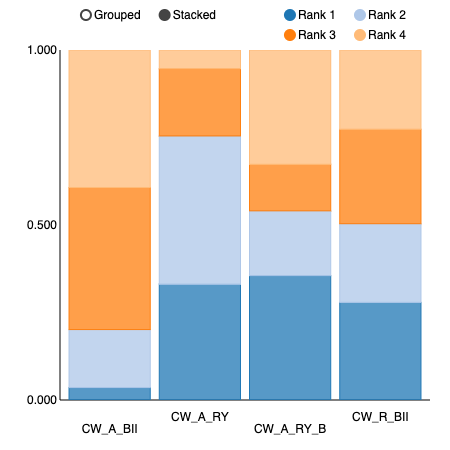

PP and PR POPF Grade B/C

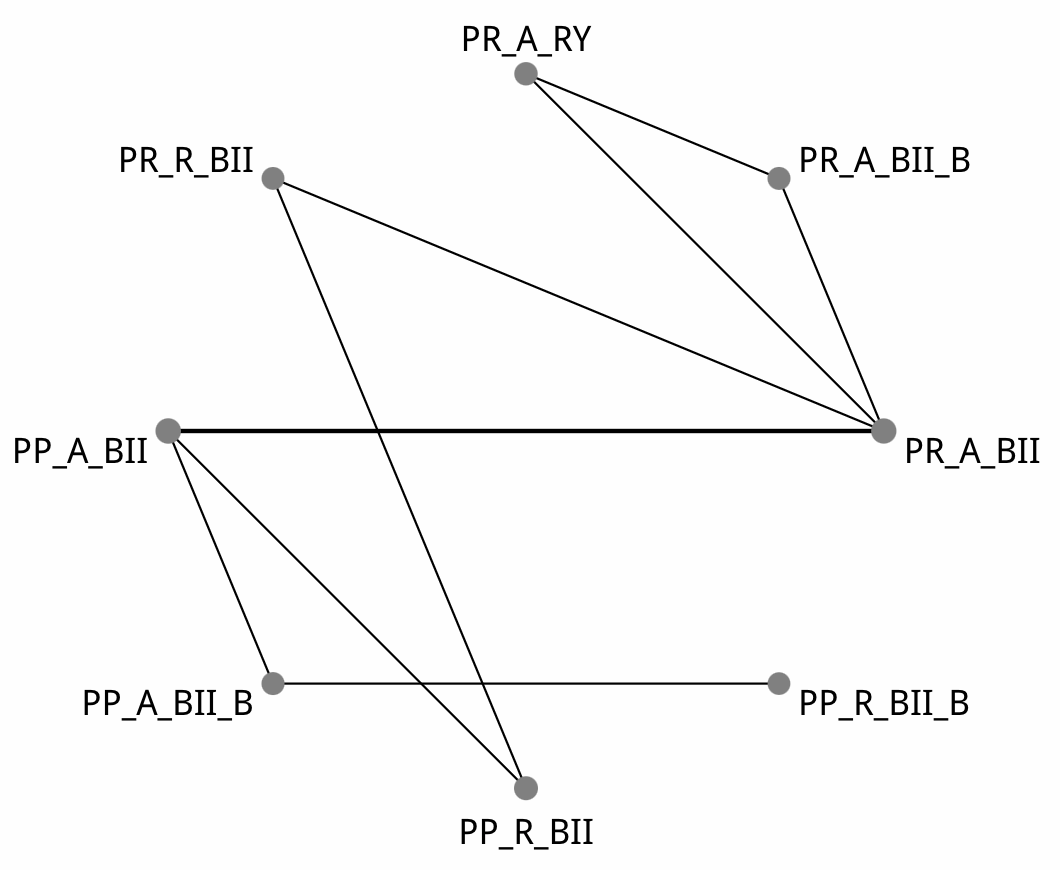

| Comparison of the included interventions: odds ratio (95% CrI). Each cell gives the effect of the column-defining intervention relative to the row-defining intervention. | | | | | | | |
| --- | --- | --- | --- | --- | --- | --- | --- |
| PP_A_BII | 2.473 ( 0.143, 103.348) | 1.191 ( 0.337, 4.350) | 2.865 ( 0.110, 148.369) | 0.611 ( 0.217, 1.759) | 0.730 ( 0.136, 4.035) | 0.461 ( 0.085, 2.287) | 1.150 ( 0.217, 6.495) |
|  | PP_A_BII_B | 0.487 ( 0.010, 10.792) | 1.146 ( 0.239, 5.216) | 0.248 ( 0.006, 5.142) | 0.287 ( 0.005, 8.589) | 0.179 ( 0.004, 4.929) | 0.460 ( 0.009, 12.117) |
|  |  | PP_R_BII | 2.382 ( 0.073, 145.576) | 0.510 ( 0.107, 2.248) | 0.597 ( 0.077, 4.520) | 0.383 ( 0.049, 2.710) | 0.960 ( 0.234, 3.838) |
|  |  |  | PP_R_BII_B | 0.214 ( 0.004, 6.877) | 0.246 ( 0.004, 10.546) | 0.154 ( 0.002, 6.395) | 0.402 ( 0.006, 16.540) |
|  |  |  |  | PR_A_BII | 1.178 ( 0.323, 4.489) | 0.748 ( 0.200, 2.576) | 1.878 ( 0.345, 11.080) |
|  |  |  |  |  | PR_A_BII_B | 0.629 ( 0.160, 2.260) | 1.567 ( 0.188, 15.189) |
|  |  |  |  |  |  | PR_A_RY | 2.522 ( 0.298, 22.608) |
|  |  |  |  |  |  |  | PR_R_BII |

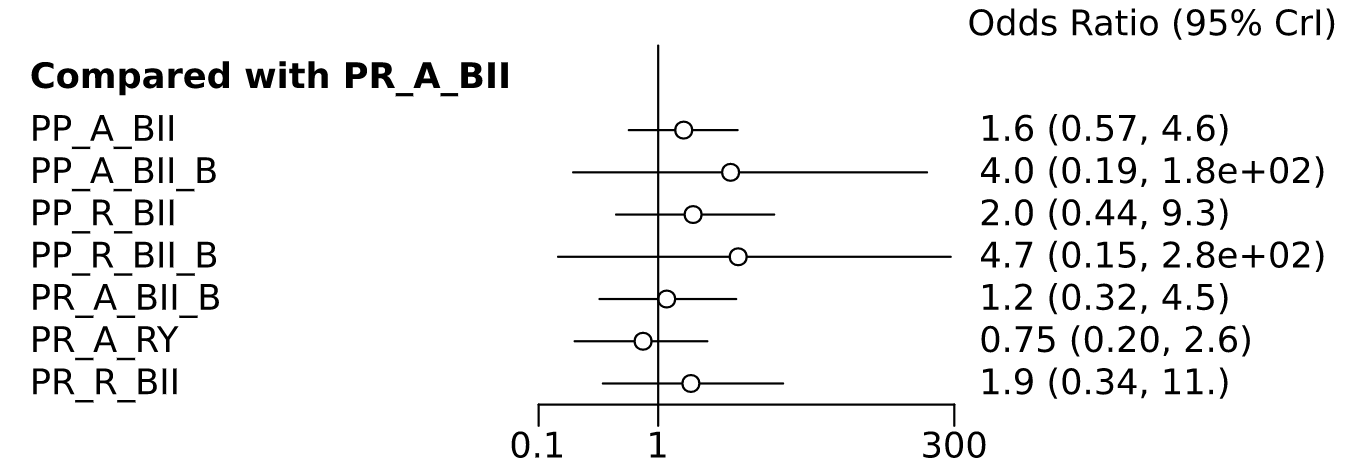

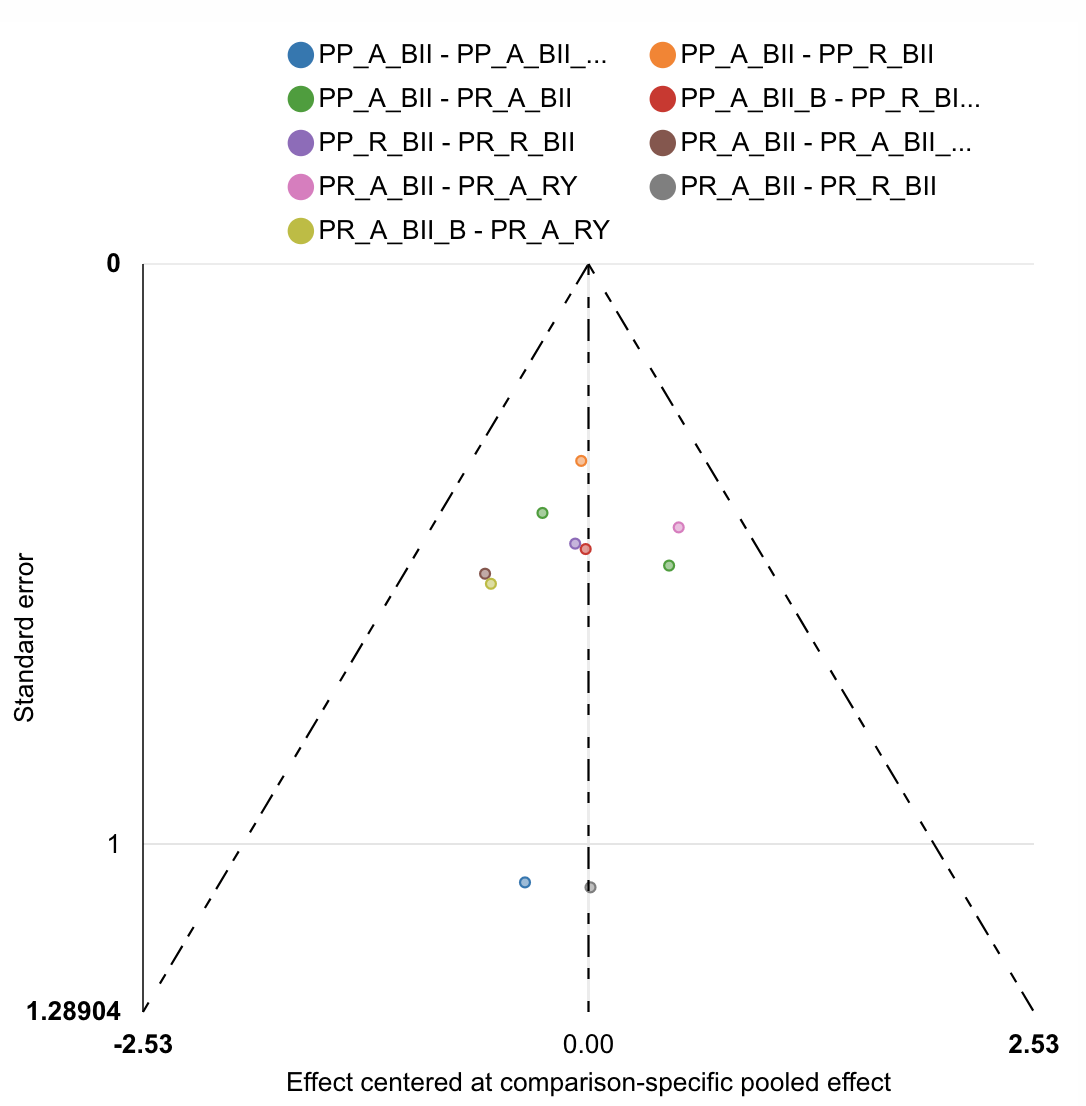

| Rank probabilities table | | | | | | | | |
| --- | --- | --- | --- | --- | --- | --- | --- | --- |
|  | **Rank 1** | **Rank 2** | **Rank 3** | **Rank 4** | **Rank 5** | **Rank 6** | **Rank 7** | **Rank 8** |
| PP_A_BII | 0.025 | 0.054 | 0.119 | 0.270 | 0.230 | 0.202 | 0.067 | 0.034 |
| PP_A_BII_B | 0.059 | 0.070 | 0.047 | 0.055 | 0.067 | 0.078 | 0.377 | 0.248 |
| PP_R_BII | 0.042 | 0.054 | 0.073 | 0.133 | 0.236 | 0.230 | 0.131 | 0.102 |
| PP_R_BII_B | 0.082 | 0.063 | 0.037 | 0.044 | 0.057 | 0.068 | 0.234 | 0.416 |
| PR_A_BII | 0.130 | 0.282 | 0.295 | 0.150 | 0.085 | 0.040 | 0.014 | 0.005 |
| PR_A_BII_B | 0.113 | 0.201 | 0.200 | 0.136 | 0.114 | 0.127 | 0.053 | 0.056 |
| PR_A_RY | 0.468 | 0.199 | 0.134 | 0.075 | 0.055 | 0.043 | 0.019 | 0.008 |
| PR_R_BII | 0.081 | 0.077 | 0.097 | 0.138 | 0.156 | 0.214 | 0.106 | 0.132 |

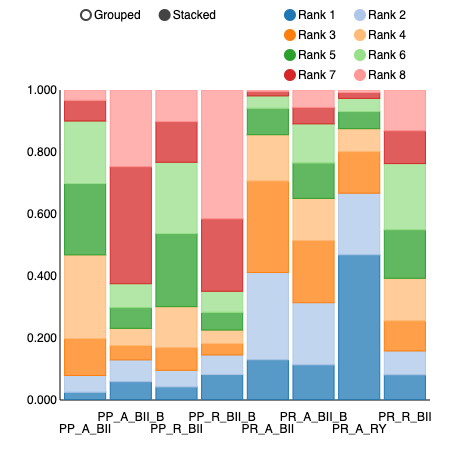

Postpancreatectomy Haemorrhage

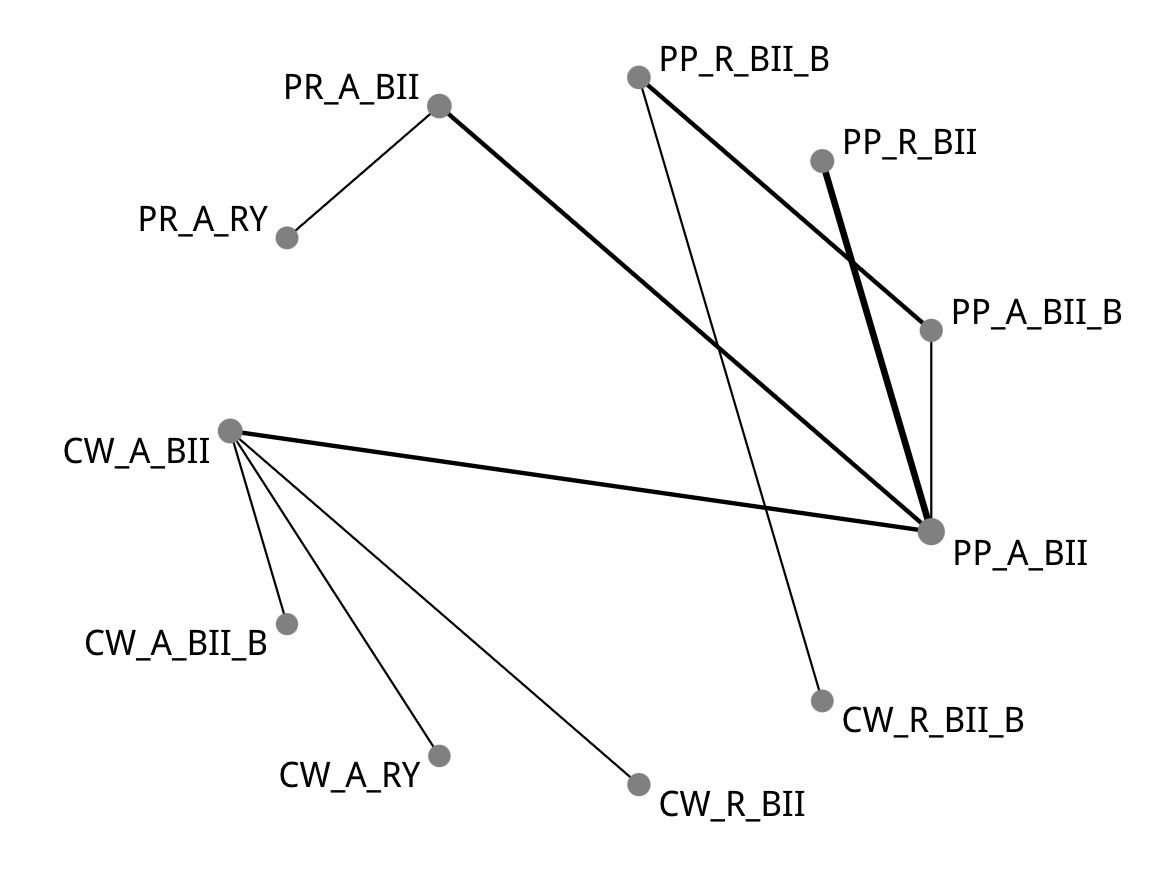

| Comparison of the included interventions: odds ratio (95% CrI). Each cell gives the effect of the column-defining intervention relative to the row-defining intervention. | | | | | | | | | | |
| --- | --- | --- | --- | --- | --- | --- | --- | --- | --- | --- |
| CW_A_BII | 0.373 ( 0.011, 6.868) | 0.539 ( 0.074, 3.443) | 0.935 ( 0.197, 4.357) | 0.470 ( 0.004, 32.809) | 0.761 ( 0.184, 3.001) | 0.298 ( 0.006, 7.489) | 1.365 ( 0.230, 7.689) | 0.214 ( 0.003, 7.354) | 0.905 ( 0.129, 5.833) | 4.947 ( 0.211, 203.588) |
|  | CW_A_BII_B | 1.438 ( 0.037, 76.028) | 2.551 ( 0.095, 113.591) | 1.232 ( 0.005, 285.574) | 2.020 ( 0.088, 91.964) | 0.753 ( 0.006, 89.104) | 3.691 ( 0.128, 188.387) | 0.537 ( 0.004, 77.285) | 2.394 ( 0.081, 134.909) | 13.754 ( 0.195, 2,181.134) |
|  |  | CW_A_RY | 1.741 ( 0.153, 20.493) | 0.838 ( 0.006, 91.469) | 1.394 ( 0.130, 15.794) | 0.560 ( 0.007, 22.558) | 2.519 ( 0.186, 34.220) | 0.400 ( 0.004, 24.444) | 1.670 ( 0.116, 26.353) | 9.370 ( 0.235, 598.903) |
|  |  |  | CW_R_BII | 0.492 ( 0.004, 44.674) | 0.804 ( 0.099, 6.027) | 0.313 ( 0.005, 11.085) | 1.487 ( 0.136, 14.701) | 0.223 ( 0.002, 10.962) | 0.951 ( 0.078, 10.456) | 5.221 ( 0.175, 274.184) |
|  |  |  |  | CW_R_BII_B | 1.657 ( 0.028, 138.269) | 0.634 ( 0.038, 9.660) | 2.945 ( 0.044, 309.792) | 0.447 ( 0.045, 3.554) | 1.967 ( 0.025, 203.405) | 10.853 ( 0.080, 3,132.542) |
|  |  |  |  |  | PP_A_BII | 0.390 ( 0.010, 6.818) | 1.802 ( 0.594, 5.623) | 0.275 ( 0.005, 7.418) | 1.191 ( 0.328, 4.350) | 6.010 ( 0.426, 206.108) |
|  |  |  |  |  |  | PP_A_BII_B | 4.596 ( 0.201, 246.830) | 0.718 ( 0.133, 3.542) | 3.062 ( 0.128, 141.062) | 16.790 ( 0.306, 2,553.704) |
|  |  |  |  |  |  |  | PP_R_BII | 0.154 ( 0.002, 5.219) | 0.650 ( 0.112, 3.628) | 3.426 ( 0.193, 131.947) |
|  |  |  |  |  |  |  |  | PP_R_BII_B | 4.175 ( 0.123, 312.061) | 23.874 ( 0.301, 5,022.585) |
|  |  |  |  |  |  |  |  |  | PR_A_BII | 5.213 ( 0.490, 143.022) |
|  |  |  |  |  |  |  |  |  |  | PR_A_RY |

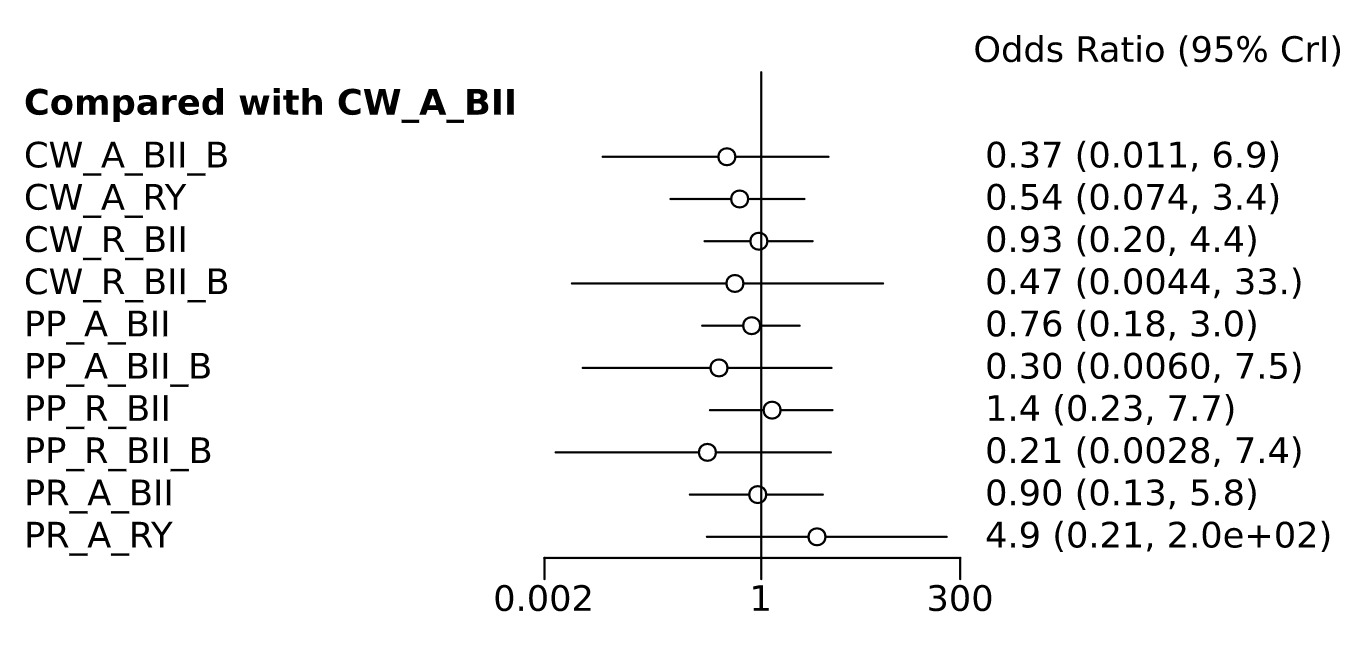

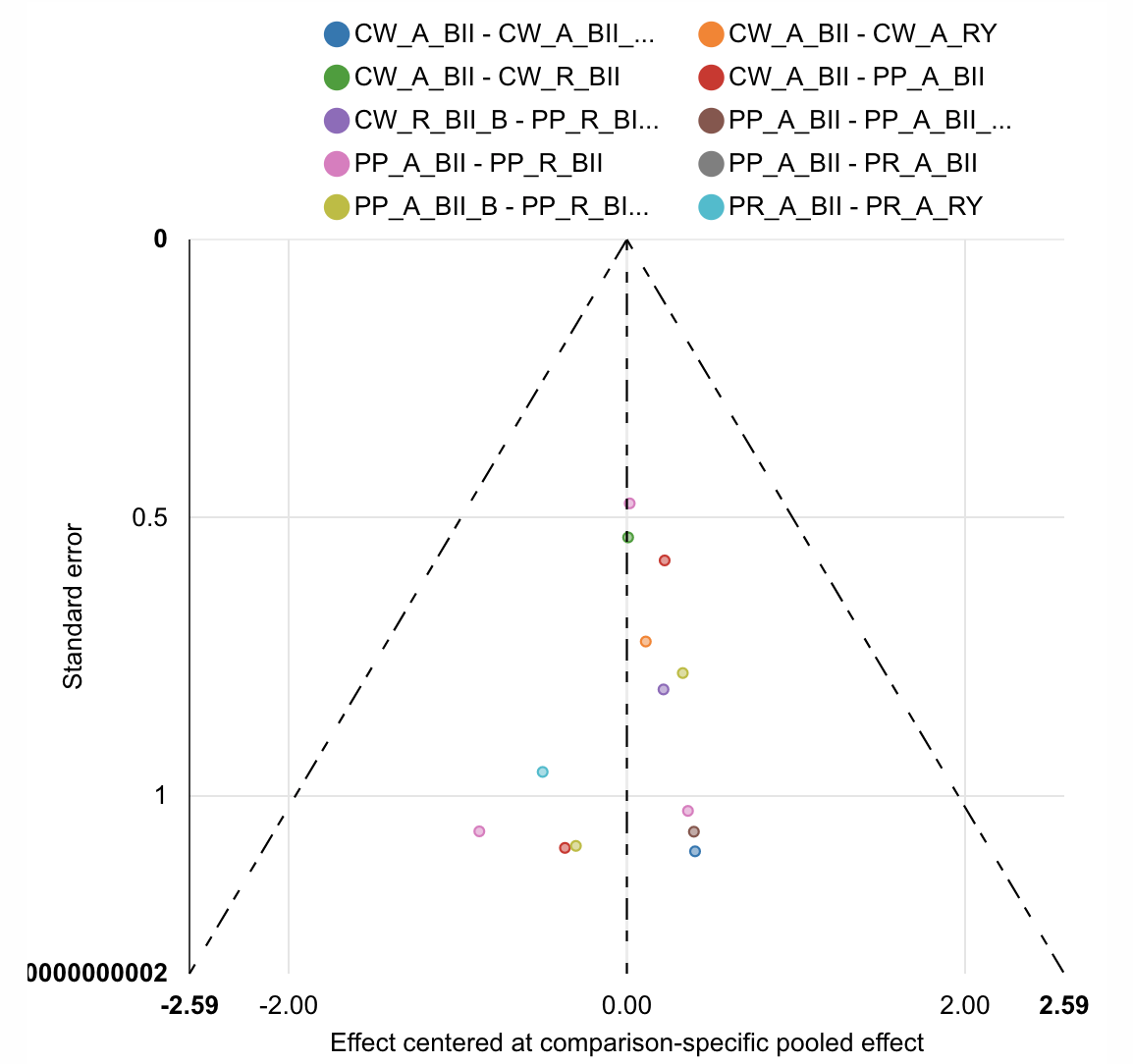

| Rank probabilities table | | | | | | | | | | | |
| --- | --- | --- | --- | --- | --- | --- | --- | --- | --- | --- | --- |
|  | **Rank 1** | **Rank 2** | **Rank 3** | **Rank 4** | **Rank 5** | **Rank 6** | **Rank 7** | **Rank 8** | **Rank 9** | **Rank 10** | **Rank 11** |
| CW_A_BII | 0.001 | 0.015 | 0.049 | 0.069 | 0.105 | 0.155 | 0.174 | 0.181 | 0.148 | 0.081 | 0.022 |
| CW_A_BII_B | 0.273 | 0.094 | 0.082 | 0.132 | 0.076 | 0.055 | 0.049 | 0.050 | 0.058 | 0.079 | 0.051 |
| CW_A_RY | 0.120 | 0.125 | 0.097 | 0.138 | 0.128 | 0.085 | 0.074 | 0.070 | 0.070 | 0.061 | 0.032 |
| CW_R_BII | 0.030 | 0.059 | 0.067 | 0.094 | 0.105 | 0.115 | 0.102 | 0.111 | 0.131 | 0.123 | 0.064 |
| CW_R_BII_B | 0.118 | 0.138 | 0.172 | 0.094 | 0.057 | 0.050 | 0.047 | 0.047 | 0.059 | 0.105 | 0.114 |
| PP_A_BII | 0.010 | 0.033 | 0.064 | 0.130 | 0.205 | 0.205 | 0.168 | 0.123 | 0.051 | 0.011 | 0.001 |
| PP_A_BII_B | 0.115 | 0.217 | 0.237 | 0.112 | 0.065 | 0.052 | 0.050 | 0.052 | 0.055 | 0.034 | 0.011 |
| PP_R_BII | 0.006 | 0.014 | 0.020 | 0.042 | 0.063 | 0.091 | 0.132 | 0.137 | 0.186 | 0.233 | 0.078 |
| PP_R_BII_B | 0.293 | 0.248 | 0.133 | 0.071 | 0.045 | 0.040 | 0.038 | 0.045 | 0.048 | 0.033 | 0.007 |
| PR_A_BII | 0.025 | 0.045 | 0.065 | 0.100 | 0.127 | 0.127 | 0.132 | 0.129 | 0.135 | 0.105 | 0.009 |
| PR_A_RY | 0.009 | 0.010 | 0.014 | 0.018 | 0.025 | 0.026 | 0.035 | 0.054 | 0.061 | 0.137 | 0.610 |

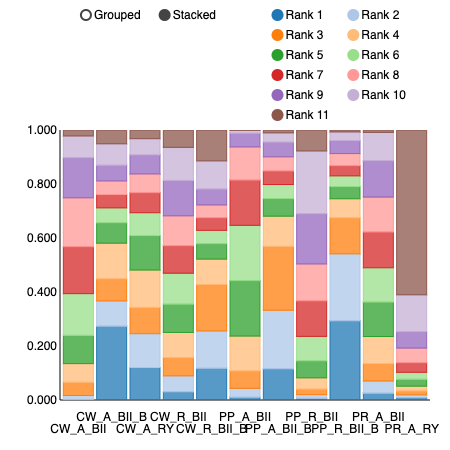

Intraoperative Blood Loss (mL)

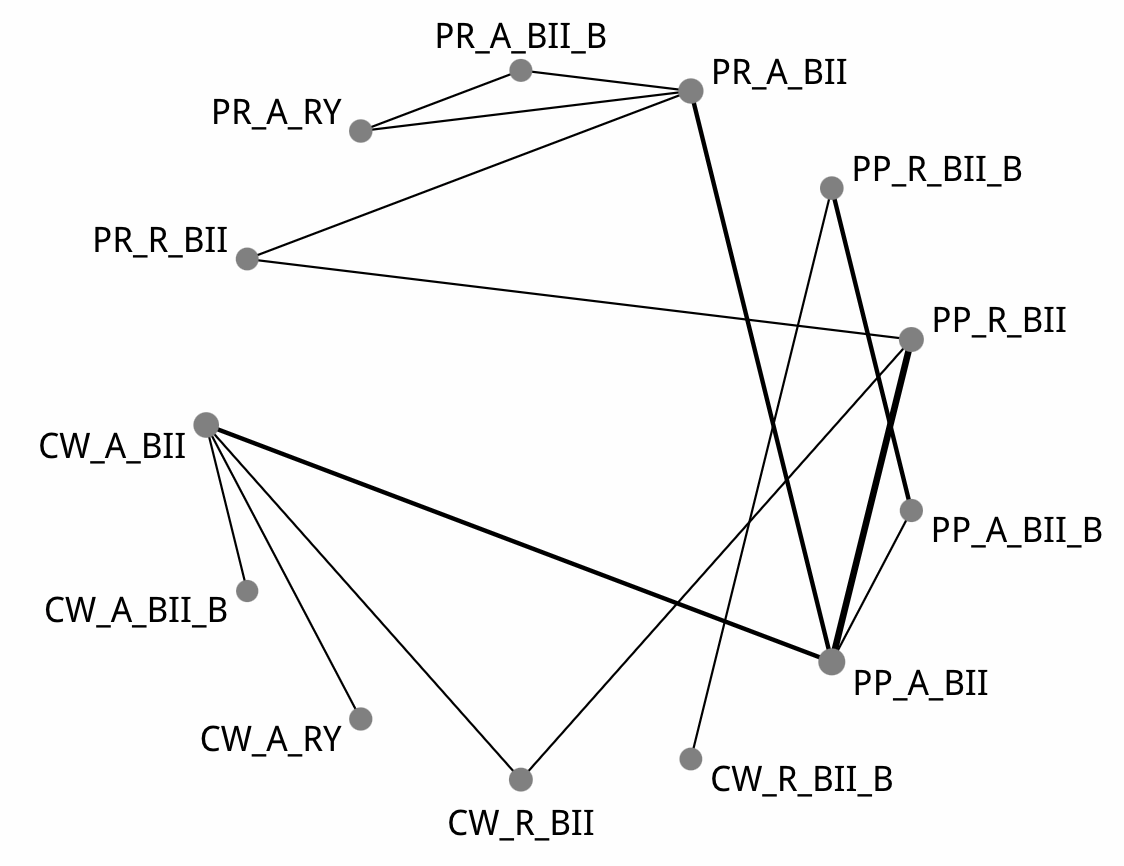

| Comparison of the included interventions: mean difference (95% CrI). Each cell gives the effect of the column-defining intervention relative to the row-defining intervention. | | | | | | | | | | | | |
| --- | --- | --- | --- | --- | --- | --- | --- | --- | --- | --- | --- | --- |
| CW_A_BII | -126.820 ( -325.920, 73.152) | -28.724 ( -201.740, 154.210) | -2.821 ( -193.390, 201.560) | 160.030 ( -470.050, 799.060) | -46.103 ( -297.040, 216.860) | -24.818 ( -374.700, 345.180) | -20.788 ( -271.200, 243.250) | -248.970 ( -753.350, 269.200) | -52.084 ( -352.970, 289.160) | -14.388 ( -388.030, 408.530) | -6.449 ( -380.980, 409.380) | 218.000 ( -139.210, 573.630) |
|  | CW_A_BII_B | 97.195 ( -164.590, 369.730) | 125.810 ( -155.220, 405.090) | 287.020 ( -392.890, 944.050) | 79.613 ( -237.250, 412.670) | 103.120 ( -297.870, 518.640) | 103.680 ( -207.570, 431.660) | -118.020 ( -682.790, 432.940) | 74.955 ( -286.840, 463.430) | 113.660 ( -310.670, 577.980) | 119.740 ( -295.470, 579.220) | 345.810 ( -52.778, 747.180) |
|  |  | CW_A_RY | 26.345 ( -232.020, 294.020) | 185.640 ( -462.350, 845.840) | -19.872 ( -322.700, 299.720) | 4.568 ( -399.800, 404.780) | 8.442 ( -285.890, 316.320) | -217.840 ( -761.170, 325.830) | -26.667 ( -360.810, 359.290) | 13.897 ( -396.900, 472.480) | 17.229 ( -378.590, 479.160) | 246.420 ( -143.570, 633.540) |
|  |  |  | CW_R_BII | 160.430 ( -466.860, 791.300) | -44.211 ( -276.540, 193.670) | -22.791 ( -359.760, 331.350) | -16.325 ( -238.840, 206.890) | -247.780 ( -756.770, 255.220) | -48.214 ( -342.920, 265.550) | -11.688 ( -390.650, 391.960) | -4.304 ( -370.360, 392.690) | 217.950 ( -117.780, 556.180) |
|  |  |  |  | CW_R_BII_B | -201.840 ( -778.150, 367.320) | -179.640 ( -698.930, 334.030) | -174.820 ( -768.040, 409.780) | -405.730 ( -796.680, -15.615) | -207.000 ( -805.200, 394.160) | -173.800 ( -801.090, 503.200) | -168.270 ( -800.870, 491.240) | 65.845 ( -582.880, 694.610) |
|  |  |  |  |  | PP_A_BII | 24.635 ( -226.550, 282.520) | 25.018 ( -99.874, 163.500) | -201.850 ( -655.860, 244.750) | -3.982 ( -182.970, 198.650) | 32.216 ( -262.460, 370.530) | 37.991 ( -242.200, 349.730) | 265.090 ( -25.371, 546.310) |
|  |  |  |  |  |  | PP_A_BII_B | 2.826 ( -305.520, 286.100) | -228.020 ( -597.310, 145.250) | -26.958 ( -335.740, 300.200) | 8.751 ( -378.900, 431.480) | 15.319 ( -362.100, 408.780) | 236.470 ( -137.940, 620.370) |
|  |  |  |  |  |  |  | PP_R_BII | -228.890 ( -700.510, 243.960) | -29.199 ( -241.930, 196.610) | 7.944 ( -305.920, 351.060) | 14.019 ( -292.120, 336.690) | 237.470 ( -35.298, 498.180) |
|  |  |  |  |  |  |  |  | PP_R_BII_B | 201.190 ( -274.320, 686.580) | 235.440 ( -289.680, 790.750) | 244.270 ( -277.220, 775.590) | 467.550 ( -64.138, 996.960) |
|  |  |  |  |  |  |  |  |  | PR_A_BII | 35.288 ( -204.980, 300.540) | 44.684 ( -180.530, 277.750) | 271.370 ( -45.794, 561.500) |
|  |  |  |  |  |  |  |  |  |  | PR_A_BII_B | 6.904 ( -212.800, 214.990) | 230.940 ( -167.100, 592.840) |
|  |  |  |  |  |  |  |  |  |  |  | PR_A_RY | 227.590 ( -163.640, 583.920) |
|  |  |  |  |  |  |  |  |  |  |  |  | PR_R_BII |

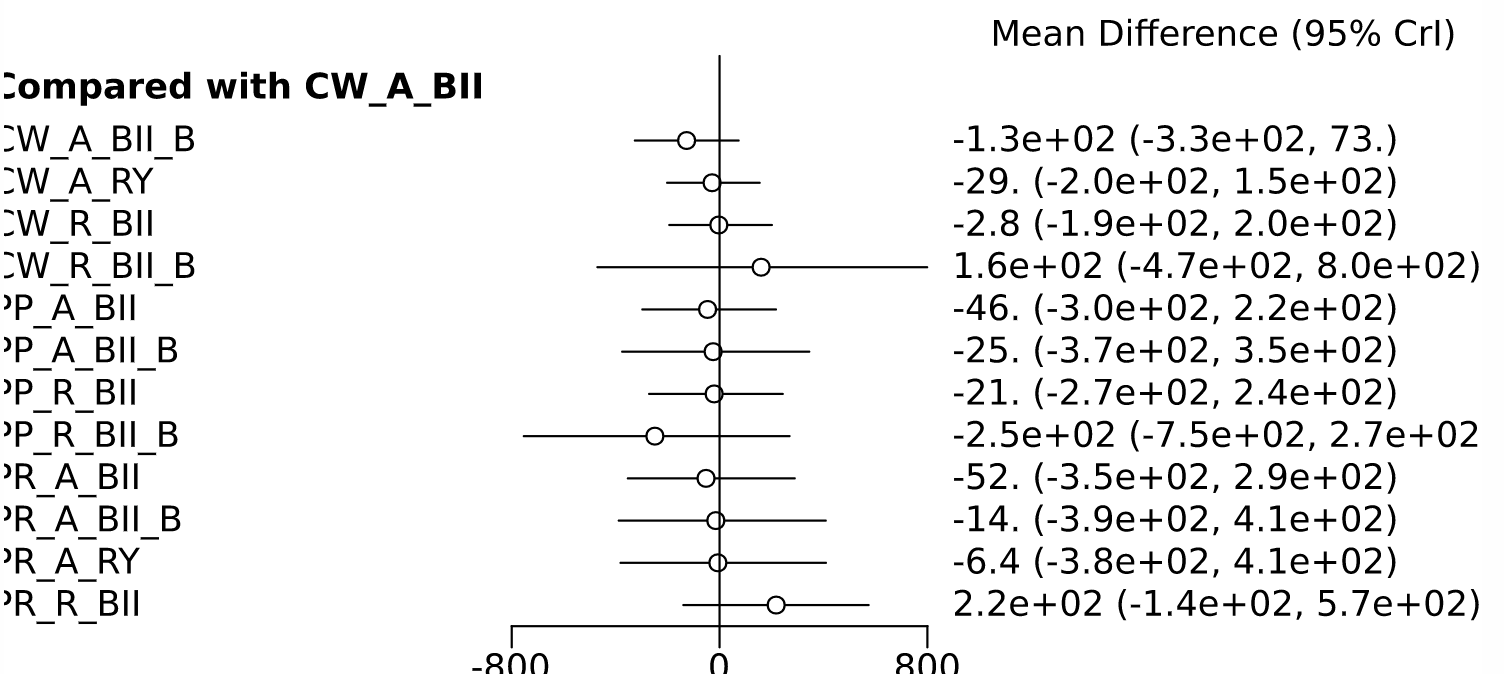

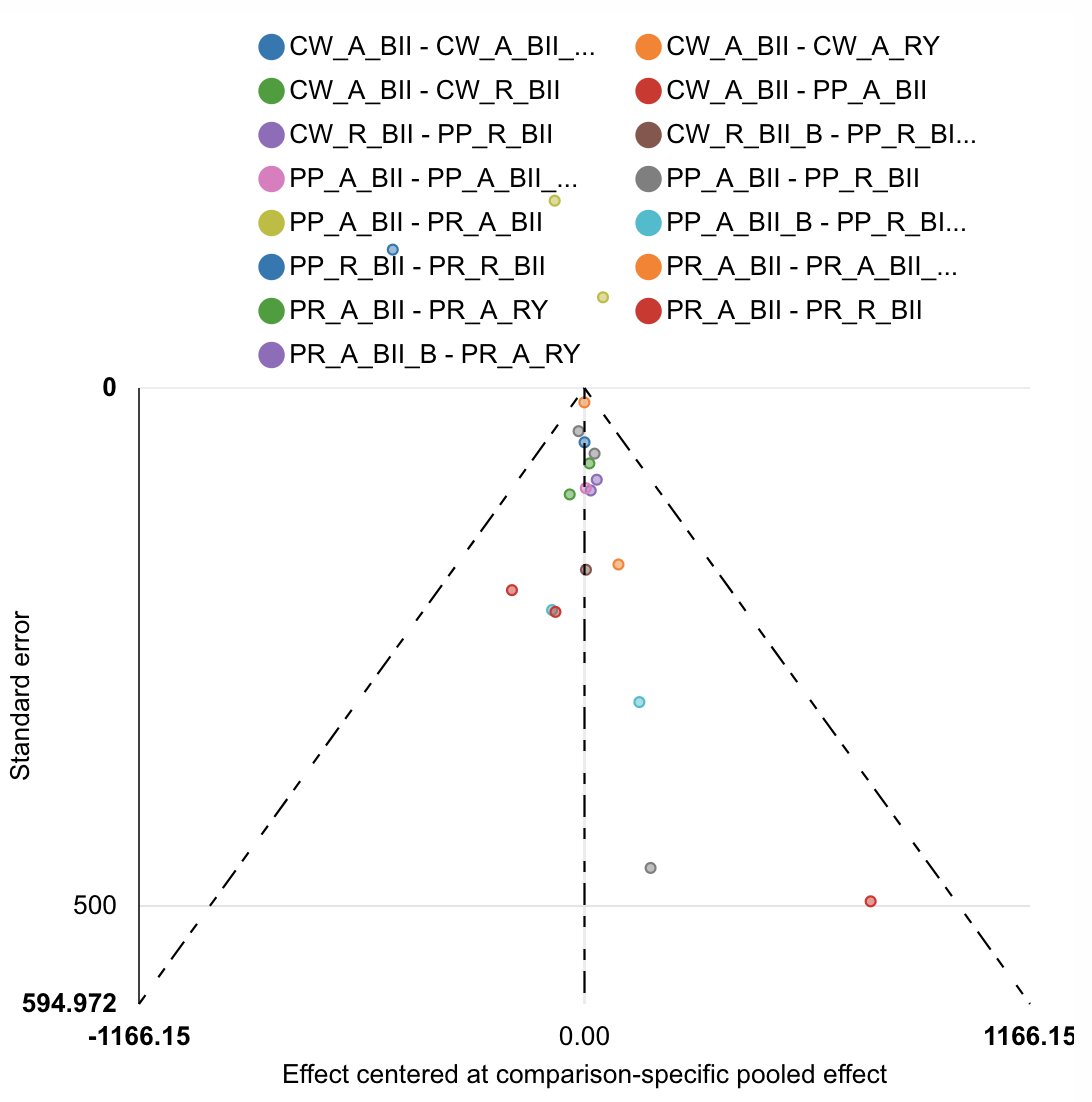

| Rank probabilities table | | | | | | | | | | | | | |
| --- | --- | --- | --- | --- | --- | --- | --- | --- | --- | --- | --- | --- | --- |
|  | **Rank 1** | **Rank 2** | **Rank 3** | **Rank 4** | **Rank 5** | **Rank 6** | **Rank 7** | **Rank 8** | **Rank 9** | **Rank 10** | **Rank 11** | **Rank 12** | **Rank 13** |
| CW_A_BII | 0.003 | 0.020 | 0.060 | 0.100 | 0.081 | 0.079 | 0.082 | 0.103 | 0.109 | 0.133 | 0.122 | 0.087 | 0.022 |
| CW_A_BII_B | 0.216 | 0.224 | 0.131 | 0.086 | 0.066 | 0.059 | 0.054 | 0.048 | 0.050 | 0.029 | 0.020 | 0.012 | 0.006 |
| CW_A_RY | 0.029 | 0.089 | 0.110 | 0.090 | 0.086 | 0.082 | 0.084 | 0.086 | 0.101 | 0.091 | 0.081 | 0.050 | 0.023 |
| CW_R_BII | 0.010 | 0.033 | 0.055 | 0.077 | 0.095 | 0.092 | 0.097 | 0.111 | 0.118 | 0.118 | 0.107 | 0.071 | 0.017 |
| CW_R_BII_B | 0.013 | 0.104 | 0.045 | 0.032 | 0.024 | 0.028 | 0.030 | 0.027 | 0.034 | 0.045 | 0.053 | 0.176 | 0.390 |
| PP_A_BII | 0.009 | 0.046 | 0.092 | 0.130 | 0.162 | 0.161 | 0.137 | 0.111 | 0.085 | 0.045 | 0.019 | 0.005 | 0.001 |
| PP_A_BII_B | 0.014 | 0.099 | 0.103 | 0.084 | 0.082 | 0.083 | 0.082 | 0.080 | 0.079 | 0.091 | 0.118 | 0.074 | 0.012 |
| PP_R_BII | 0.010 | 0.026 | 0.058 | 0.072 | 0.106 | 0.130 | 0.148 | 0.147 | 0.120 | 0.092 | 0.065 | 0.026 | 0.001 |
| PP_R_BII_B | 0.574 | 0.116 | 0.055 | 0.043 | 0.032 | 0.026 | 0.027 | 0.022 | 0.025 | 0.037 | 0.025 | 0.019 | 0.000 |
| PR_A_BII | 0.034 | 0.083 | 0.114 | 0.130 | 0.121 | 0.114 | 0.089 | 0.094 | 0.090 | 0.076 | 0.038 | 0.016 | 0.002 |
| PR_A_BII_B | 0.054 | 0.090 | 0.088 | 0.073 | 0.066 | 0.067 | 0.073 | 0.070 | 0.078 | 0.096 | 0.113 | 0.100 | 0.034 |
| PR_A_RY | 0.033 | 0.069 | 0.086 | 0.077 | 0.074 | 0.069 | 0.083 | 0.077 | 0.080 | 0.099 | 0.139 | 0.086 | 0.030 |
| PR_R_BII | 0.001 | 0.003 | 0.005 | 0.007 | 0.007 | 0.011 | 0.017 | 0.024 | 0.033 | 0.050 | 0.101 | 0.281 | 0.461 |

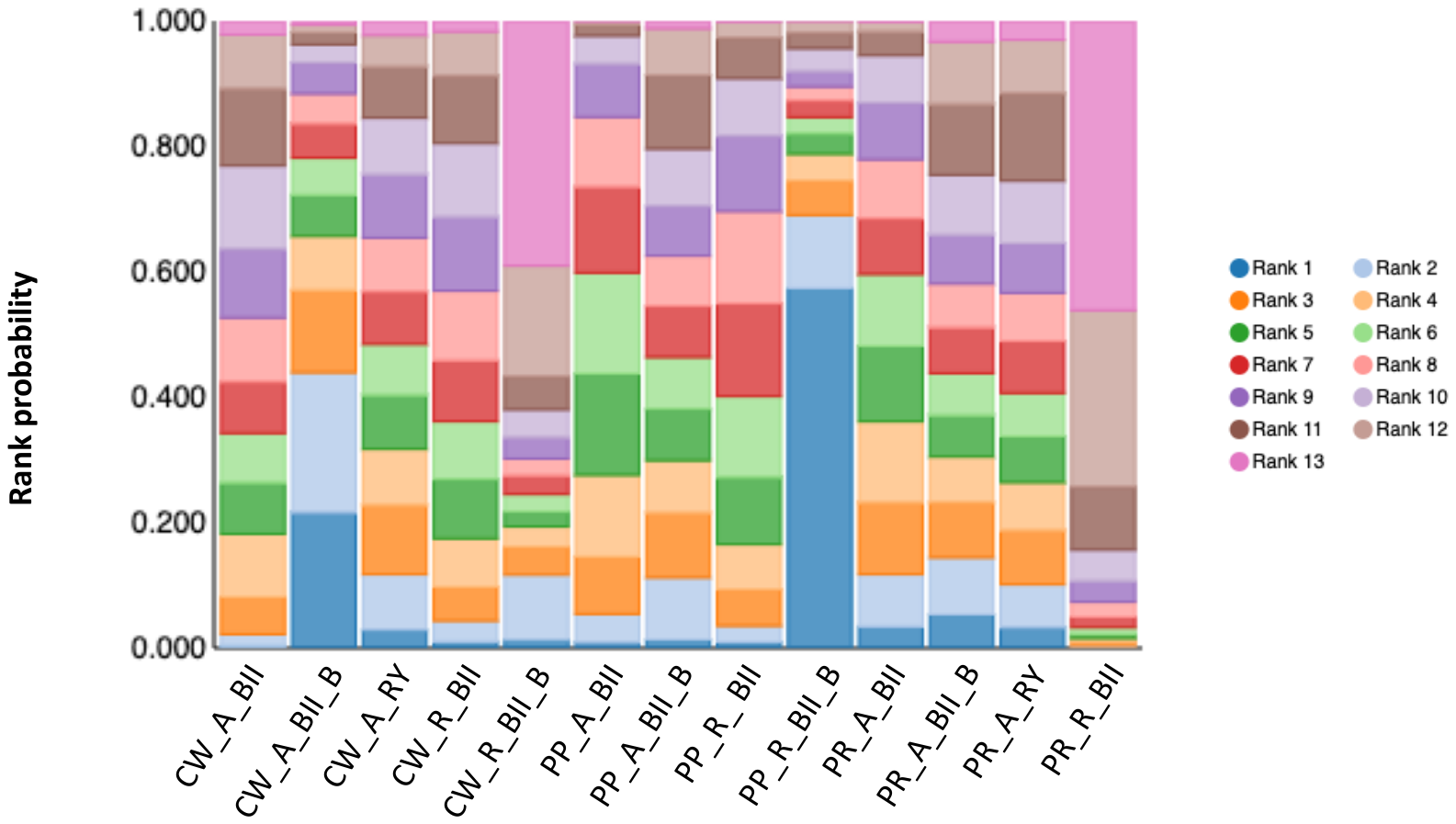

Duration of operation (minutes)

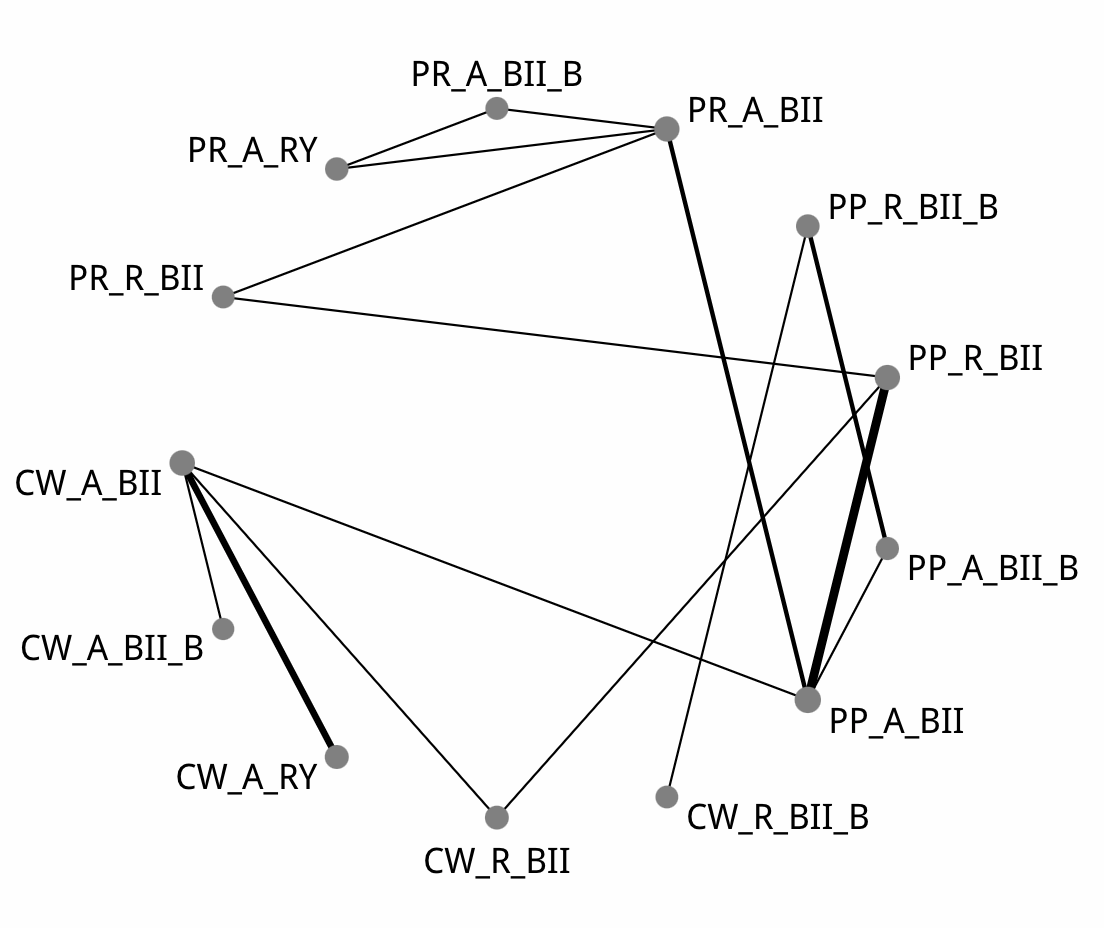

| Comparison of the included interventions: mean difference (95% CrI). Each cell gives the effect of the column-defining intervention relative to the row-defining intervention. | | | | | | | | | | | | |
| --- | --- | --- | --- | --- | --- | --- | --- | --- | --- | --- | --- | --- |
| CW_A_BII | 11.472 ( -68.462, 86.518) | 8.337 ( -6.413, 27.715) | -5.212 ( -37.290, 26.121) | -4.711 ( -93.361, 80.330) | **-30.059 ( -58.293, -4.430)** | -36.941 ( -96.799, 21.968) | -24.971 ( -56.266, 2.289) | **-70.853 ( -140.810, -3.678)** | -25.964 ( -61.298, 7.303) | -1.734 ( -55.182, 47.490) | 1.836 ( -47.118, 49.483) | 8.333 ( -48.780, 62.452) |
|  | CW_A_BII_B | -1.929 ( -79.348, 78.688) | -16.990 ( -95.480, 67.139) | -14.604 ( -128.770, 101.170) | -41.516 ( -120.560, 41.117) | -47.528 ( -142.740, 50.723) | -36.330 ( -115.730, 46.417) | -82.180 ( -180.690, 24.144) | -37.075 ( -116.190, 46.212) | -13.397 ( -100.900, 76.291) | -9.361 ( -97.808, 81.526) | -3.426 ( -94.627, 89.632) |
|  |  | CW_A_RY | -14.097 ( -51.374, 21.040) | -13.524 ( -103.520, 73.494) | **-38.603 ( -72.776, -8.816)** | -45.691 ( -109.390, 15.257) | **-33.765 ( -71.592, -2.730)** | **-79.819 ( -152.820, -10.593)** | -34.737 ( -75.271, 0.729) | -10.109 ( -68.057, 40.232) | -6.841 ( -60.249, 42.063) | -0.470 ( -62.571, 55.425) |
|  |  |  | CW_R_BII | 0.638 ( -87.906, 87.690) | -24.817 ( -57.324, 8.292) | -32.063 ( -91.113, 27.457) | -19.809 ( -51.922, 11.066) | -65.064 ( -137.510, 4.284) | -20.473 ( -59.739, 19.054) | 3.068 ( -51.123, 55.906) | 6.698 ( -44.154, 56.820) | 13.341 ( -46.429, 70.077) |
|  |  |  |  | CW_R_BII_B | -25.256 ( -107.590, 56.956) | -32.174 ( -98.625, 34.090) | -20.813 ( -105.650, 63.587) | **-66.152 ( -120.490, -12.284)** | -21.279 ( -105.890, 63.261) | 2.678 ( -90.503, 95.132) | 6.413 ( -84.043, 95.317) | 11.389 ( -82.967, 110.000) |
|  |  |  |  |  | PP_A_BII | -6.722 ( -58.293, 45.460) | 4.986 ( -13.049, 20.956) | -40.555 ( -105.260, 21.608) | 4.016 ( -17.966, 25.769) | 27.804 ( -17.523, 71.359) | 31.452 ( -9.300, 71.366) | 38.511 ( -14.722, 87.664) |
|  |  |  |  |  |  | PP_A_BII_B | 11.628 ( -42.868, 65.330) | -33.990 ( -71.985, 5.524) | 10.614 ( -43.351, 66.904) | 34.108 ( -35.925, 101.840) | 38.208 ( -26.746, 101.830) | 44.453 ( -26.333, 115.080) |
|  |  |  |  |  |  |  | PP_R_BII | -44.954 ( -110.220, 20.202) | -1.123 ( -26.652, 27.138) | 23.067 ( -23.438, 69.486) | 26.488 ( -15.935, 70.125) | 33.778 ( -17.660, 82.250) |
|  |  |  |  |  |  |  |  | PP_R_BII_B | 43.732 ( -21.090, 111.940) | 68.019 ( -7.356, 146.520) | 71.635 ( -1.823, 147.100) | 78.720 ( -2.945, 158.910) |
|  |  |  |  |  |  |  |  |  | PR_A_BII | 23.843 ( -14.708, 61.149) | 27.051 ( -5.792, 60.735) | 34.026 ( -18.807, 83.167) |
|  |  |  |  |  |  |  |  |  |  | PR_A_BII_B | 3.574 ( -28.571, 35.663) | 10.247 ( -51.214, 73.548) |
|  |  |  |  |  |  |  |  |  |  |  | PR_A_RY | 6.503 ( -55.048, 65.548) |
|  |  |  |  |  |  |  |  |  |  |  |  | PR_R_BII |

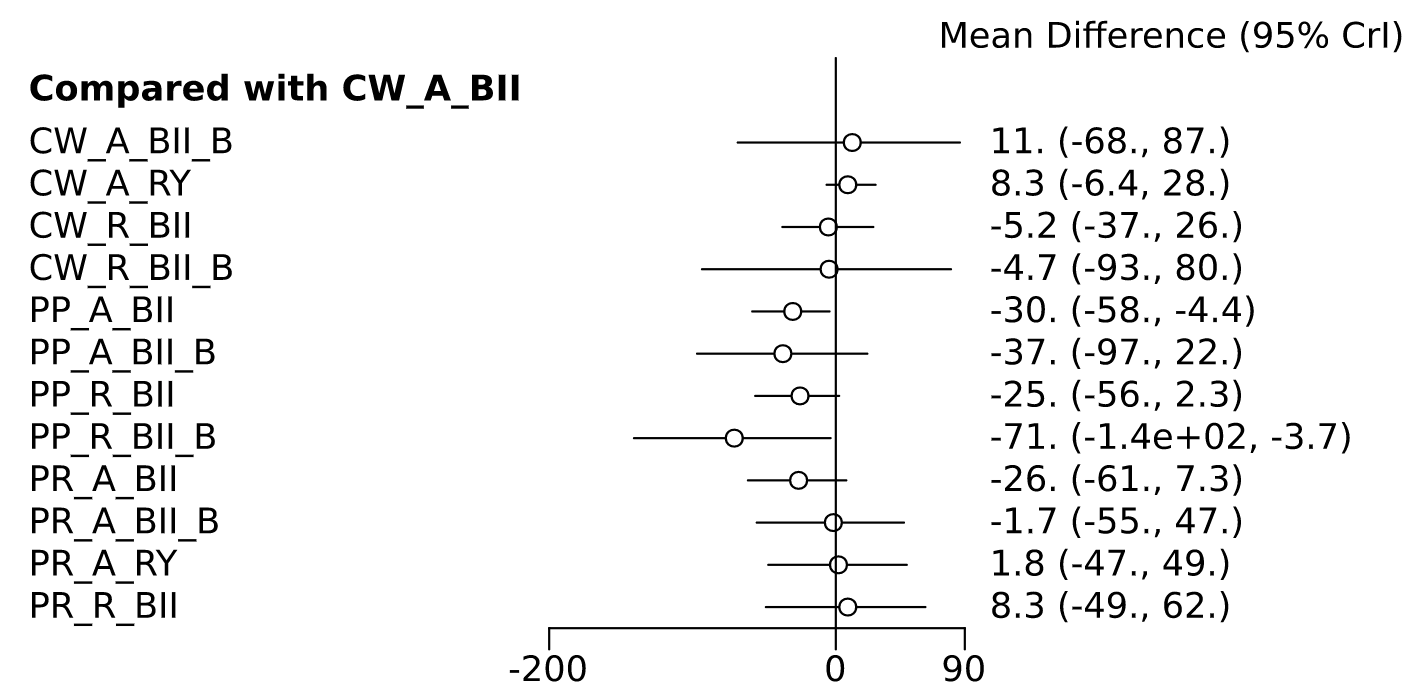

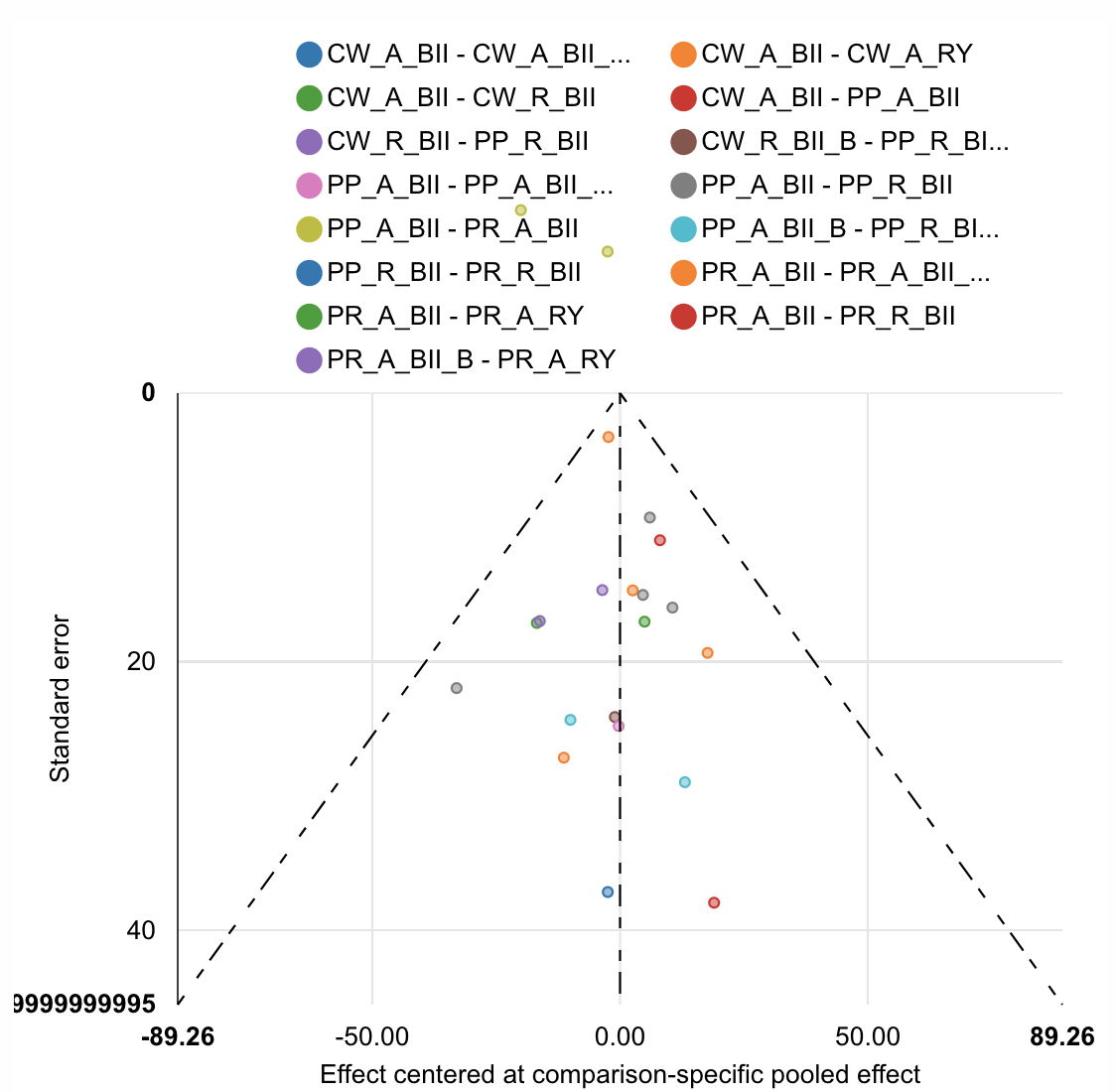

| Rank probabilities table | | | | | | | | | | | | | |
| --- | --- | --- | --- | --- | --- | --- | --- | --- | --- | --- | --- | --- | --- |
|  | **Rank 1** | **Rank 2** | **Rank 3** | **Rank 4** | **Rank 5** | **Rank 6** | **Rank 7** | **Rank 8** | **Rank 9** | **Rank 10** | **Rank 11** | **Rank 12** | **Rank 13** |
| CW_A_BII | 0.000 | 0.002 | 0.005 | 0.010 | 0.029 | 0.066 | 0.122 | 0.184 | 0.198 | 0.198 | 0.135 | 0.048 | 0.004 |
| CW_A_BII_B | 0.048 | 0.054 | 0.040 | 0.032 | 0.031 | 0.039 | 0.054 | 0.054 | 0.057 | 0.062 | 0.072 | 0.135 | 0.320 |
| CW_A_RY | 0.001 | 0.002 | 0.002 | 0.003 | 0.007 | 0.018 | 0.038 | 0.074 | 0.130 | 0.173 | 0.226 | 0.210 | 0.117 |
| CW_R_BII | 0.005 | 0.017 | 0.020 | 0.035 | 0.062 | 0.103 | 0.155 | 0.162 | 0.147 | 0.116 | 0.091 | 0.062 | 0.027 |
| CW_R_BII_B | 0.006 | 0.114 | 0.122 | 0.046 | 0.043 | 0.061 | 0.063 | 0.056 | 0.052 | 0.057 | 0.069 | 0.113 | 0.200 |
| PP_A_BII | 0.030 | 0.141 | 0.236 | 0.292 | 0.174 | 0.084 | 0.029 | 0.009 | 0.003 | 0.001 | 0.000 | 0.000 | 0.000 |
| PP_A_BII_B | 0.019 | 0.385 | 0.176 | 0.093 | 0.080 | 0.068 | 0.052 | 0.037 | 0.031 | 0.024 | 0.021 | 0.011 | 0.003 |
| PP_R_BII | 0.021 | 0.066 | 0.136 | 0.194 | 0.230 | 0.182 | 0.104 | 0.045 | 0.016 | 0.005 | 0.001 | 0.001 | 0.000 |
| PP_R_BII_B | 0.816 | 0.076 | 0.029 | 0.022 | 0.019 | 0.014 | 0.010 | 0.005 | 0.004 | 0.003 | 0.003 | 0.001 | 0.000 |
| PR_A_BII | 0.031 | 0.091 | 0.157 | 0.188 | 0.214 | 0.174 | 0.083 | 0.038 | 0.017 | 0.007 | 0.002 | 0.001 | 0.000 |
| PR_A_BII_B | 0.012 | 0.023 | 0.033 | 0.039 | 0.047 | 0.080 | 0.110 | 0.129 | 0.127 | 0.114 | 0.122 | 0.110 | 0.055 |
| PR_A_RY | 0.004 | 0.009 | 0.017 | 0.022 | 0.035 | 0.060 | 0.106 | 0.128 | 0.132 | 0.141 | 0.137 | 0.131 | 0.079 |
| PR_R_BII | 0.009 | 0.019 | 0.028 | 0.026 | 0.031 | 0.052 | 0.074 | 0.081 | 0.085 | 0.100 | 0.122 | 0.178 | 0.195 |

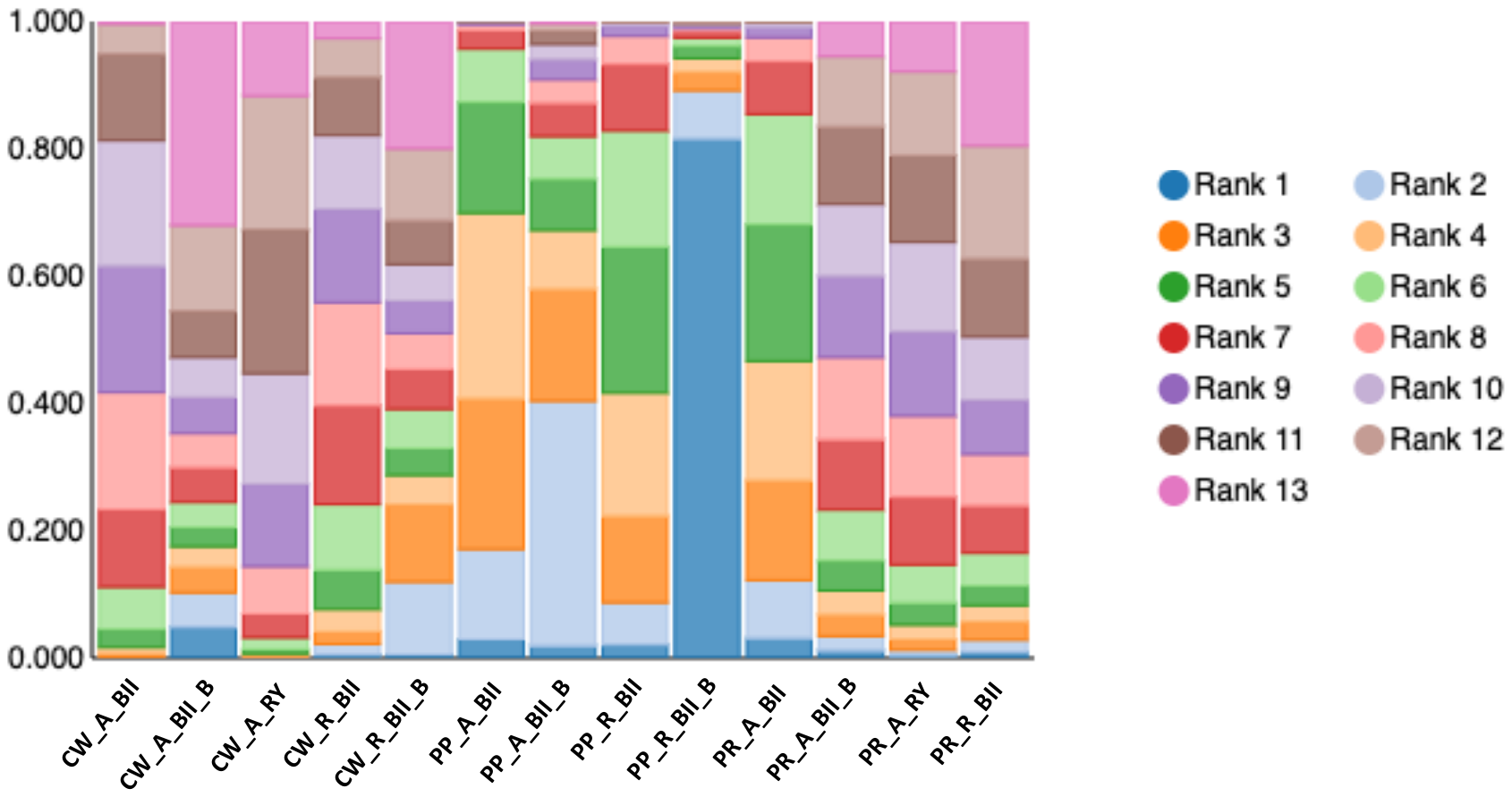

Intra-abdominal abscess

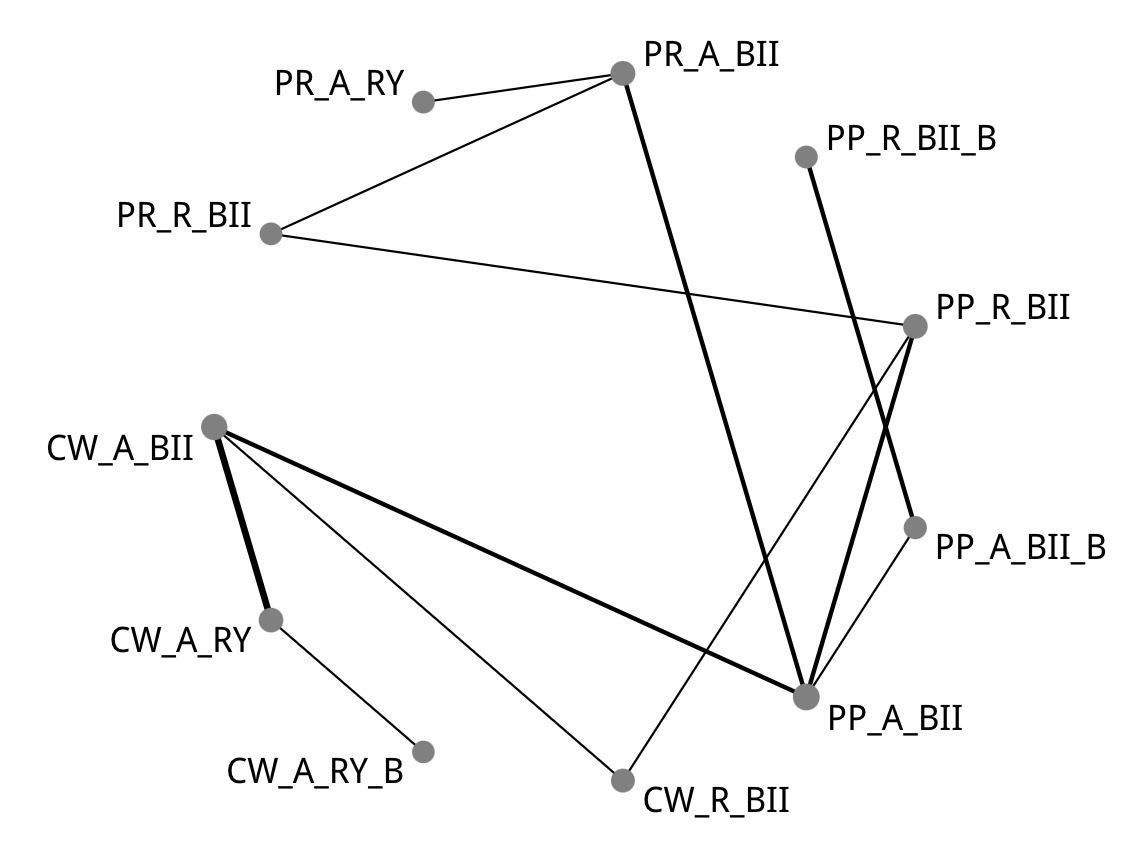

| Comparison of the included interventions: odds ratio (95% CrI). Each cell gives the effect of the column-defining intervention relative to the row-defining intervention. | | | | | | | | | | |
| --- | --- | --- | --- | --- | --- | --- | --- | --- | --- | --- |
| CW_A_BII | 1.205 ( 0.543, 2.716) | 1.464 ( 0.202, 12.098) | 0.764 ( 0.336, 1.912) | 1.009 ( 0.369, 2.745) | 0.926 ( 0.022, 37.207) | 1.254 ( 0.395, 3.980) | 0.667 ( 0.016, 31.893) | 0.785 ( 0.235, 2.746) | 0.516 ( 0.091, 3.068) | 4.790 ( 0.664, 53.282) |
|  | CW_A_RY | 1.214 ( 0.179, 8.380) | 0.628 ( 0.203, 2.197) | 0.829 ( 0.231, 3.079) | 0.763 ( 0.015, 32.596) | 1.031 ( 0.262, 4.315) | 0.554 ( 0.011, 26.133) | 0.649 ( 0.152, 2.947) | 0.429 ( 0.062, 3.189) | 4.024 ( 0.464, 51.785) |
|  |  | CW_A_RY_B | 0.514 ( 0.053, 4.695) | 0.683 ( 0.069, 6.276) | 0.624 ( 0.011, 36.749) | 0.862 ( 0.079, 8.817) | 0.451 ( 0.008, 31.897) | 0.538 ( 0.048, 5.544) | 0.353 ( 0.024, 5.275) | 3.336 ( 0.191, 80.705) |
|  |  |  | CW_R_BII | 1.312 ( 0.394, 4.186) | 1.180 ( 0.028, 50.199) | 1.630 ( 0.490, 5.368) | 0.861 ( 0.022, 37.917) | 1.026 ( 0.256, 3.936) | 0.671 ( 0.101, 4.093) | 6.301 ( 0.827, 72.574) |
|  |  |  |  | PP_A_BII | 0.926 ( 0.029, 31.284) | 1.238 ( 0.559, 2.977) | 0.670 ( 0.024, 27.295) | 0.786 ( 0.368, 1.637) | 0.521 ( 0.120, 2.144) | 4.732 ( 0.871, 51.096) |
|  |  |  |  |  | PP_A_BII_B | 1.361 ( 0.036, 48.284) | 0.741 ( 0.302, 1.829) | 0.869 ( 0.024, 33.162) | 0.555 ( 0.014, 24.369) | 5.633 ( 0.113, 314.316) |
|  |  |  |  |  |  | PP_R_BII | 0.534 ( 0.016, 23.594) | 0.636 ( 0.207, 1.781) | 0.413 ( 0.074, 2.127) | 3.769 ( 0.714, 34.918) |
|  |  |  |  |  |  |  | PP_R_BII_B | 1.187 ( 0.027, 40.993) | 0.777 ( 0.016, 32.408) | 7.665 ( 0.139, 398.936) |
|  |  |  |  |  |  |  |  | PR_A_BII | 0.660 ( 0.183, 2.304) | **6.012 ( 1.063, 65.825)** |
|  |  |  |  |  |  |  |  |  | PR_A_RY | **9.322 ( 1.078, 136.524)** |
|  |  |  |  |  |  |  |  |  |  | PR_R_BII |

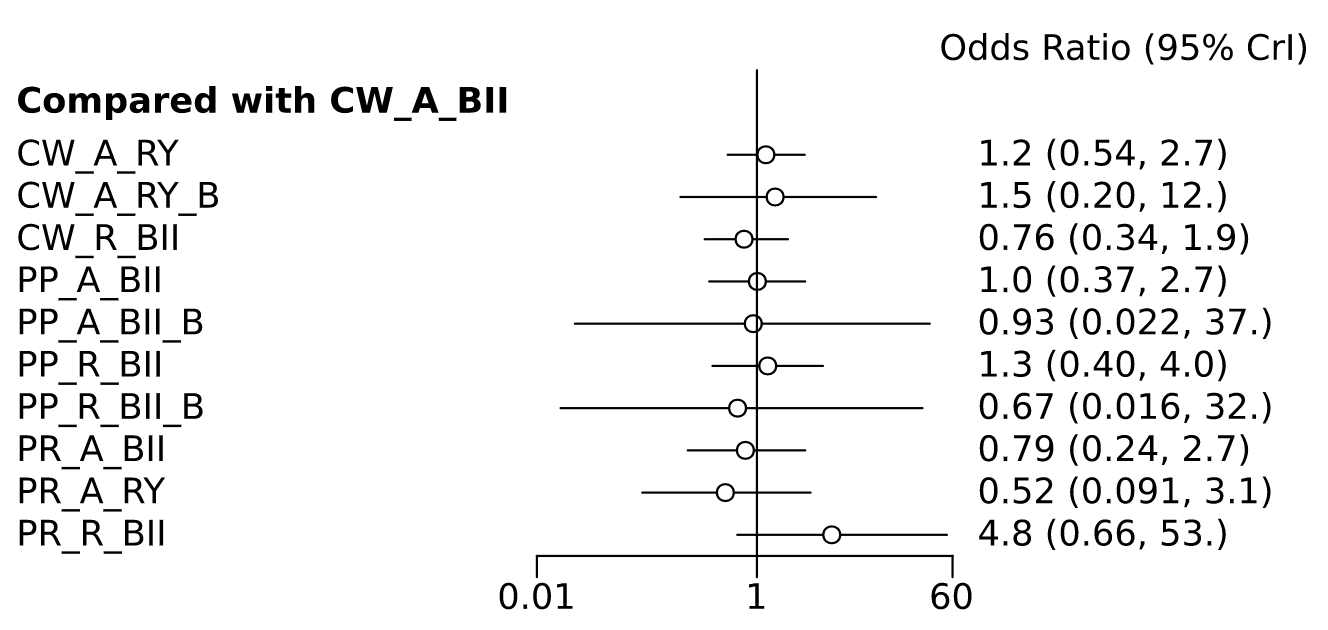

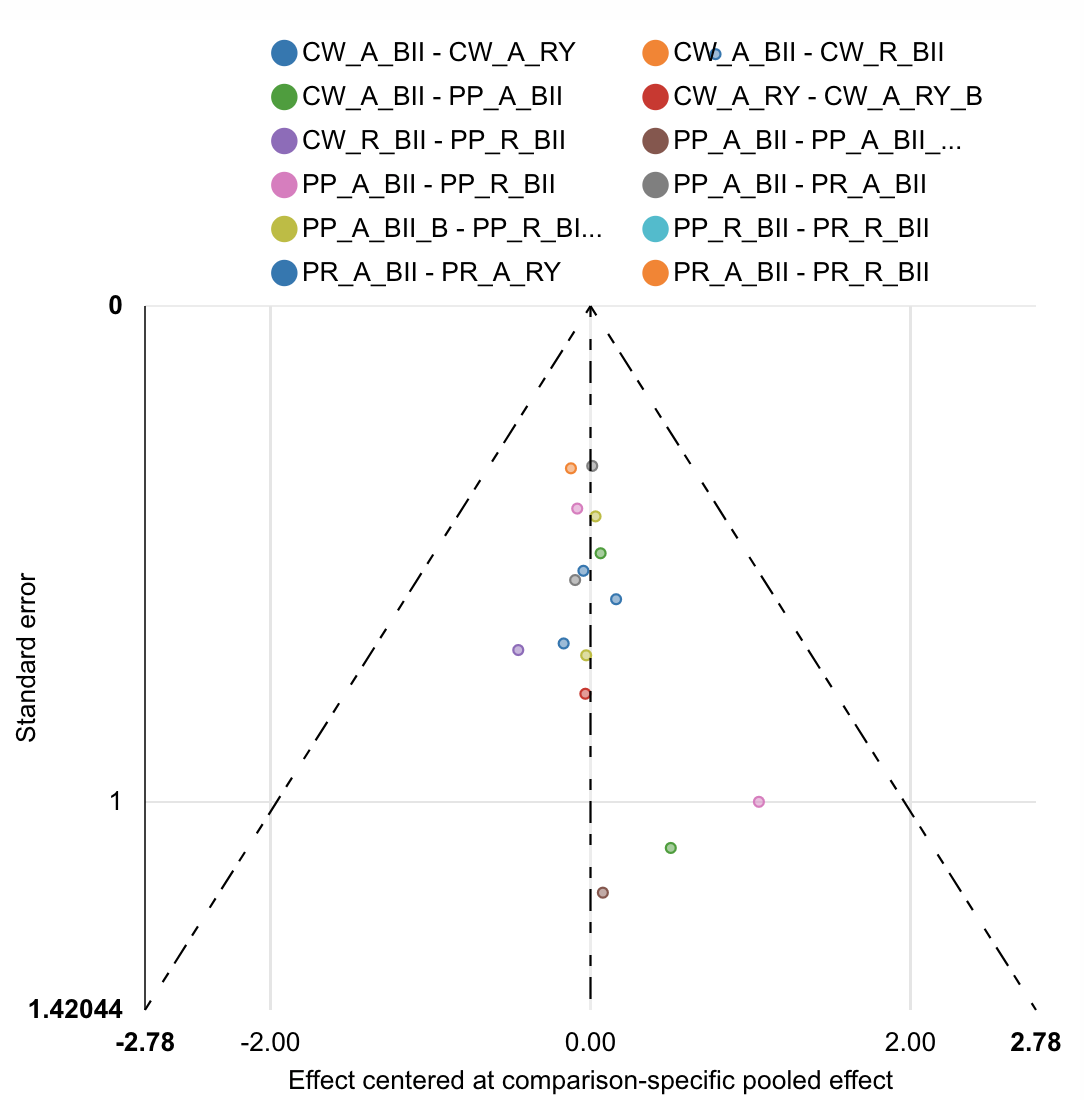

| Rank probabilities table | | | | | | | | | | | |
| --- | --- | --- | --- | --- | --- | --- | --- | --- | --- | --- | --- |
|  | **Rank 1** | **Rank 2** | **Rank 3** | **Rank 4** | **Rank 5** | **Rank 6** | **Rank 7** | **Rank 8** | **Rank 9** | **Rank 10** | **Rank 11** |
| CW_A_BII | 0.012 | 0.051 | 0.088 | 0.128 | 0.149 | 0.166 | 0.157 | 0.138 | 0.080 | 0.028 | 0.003 |
| CW_A_RY | 0.016 | 0.037 | 0.065 | 0.080 | 0.095 | 0.114 | 0.144 | 0.145 | 0.162 | 0.116 | 0.027 |
| CW_A_RY_B | 0.093 | 0.056 | 0.069 | 0.059 | 0.051 | 0.055 | 0.064 | 0.101 | 0.101 | 0.210 | 0.142 |
| CW_R_BII | 0.108 | 0.128 | 0.155 | 0.152 | 0.141 | 0.103 | 0.095 | 0.059 | 0.037 | 0.020 | 0.003 |
| PP_A_BII | 0.005 | 0.025 | 0.083 | 0.122 | 0.180 | 0.186 | 0.167 | 0.129 | 0.074 | 0.027 | 0.003 |
| PP_A_BII_B | 0.082 | 0.249 | 0.080 | 0.052 | 0.037 | 0.036 | 0.039 | 0.057 | 0.100 | 0.145 | 0.124 |
| PP_R_BII | 0.010 | 0.023 | 0.048 | 0.076 | 0.091 | 0.122 | 0.146 | 0.170 | 0.155 | 0.148 | 0.012 |
| PP_R_BII_B | 0.299 | 0.153 | 0.062 | 0.044 | 0.037 | 0.037 | 0.037 | 0.054 | 0.104 | 0.126 | 0.046 |
| PR_A_BII | 0.039 | 0.149 | 0.150 | 0.189 | 0.148 | 0.116 | 0.086 | 0.064 | 0.040 | 0.017 | 0.002 |
| PR_A_RY | 0.333 | 0.127 | 0.195 | 0.093 | 0.061 | 0.050 | 0.041 | 0.036 | 0.030 | 0.030 | 0.006 |
| PR_R_BII | 0.004 | 0.003 | 0.005 | 0.006 | 0.012 | 0.014 | 0.024 | 0.048 | 0.118 | 0.134 | 0.634 |

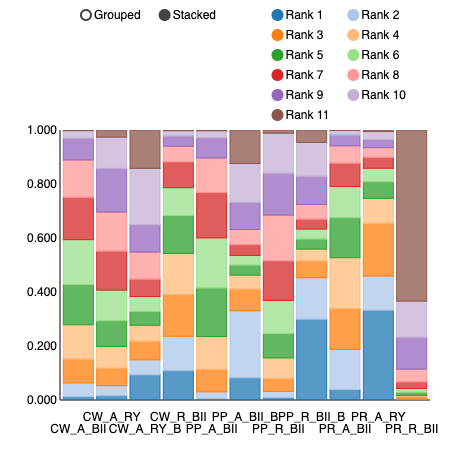

Intra-abdominal abscess sensitivity analysis with Tani et al. 2014 removed (single study comparing PR_A_RY.

| Comparison of the included interventions: odds ratio (95% CrI). Each cell gives the effect of the column-defining intervention relative to the row-defining intervention. | | | | | | | | | |
| --- | --- | --- | --- | --- | --- | --- | --- | --- | --- |
| CW_A_BII | 1.195 ( 0.546, 2.649) | 1.407 ( 0.183, 12.167) | 0.762 ( 0.348, 1.870) | 0.985 ( 0.370, 2.786) | 1.036 ( 0.016, 77.587) | 1.204 ( 0.391, 4.224) | 0.764 ( 0.012, 58.980) | 0.757 ( 0.225, 2.866) | 4.476 ( 0.713, 43.707) |
|  | CW_A_RY | 1.175 ( 0.170, 8.854) | 0.638 ( 0.207, 2.133) | 0.836 ( 0.226, 2.753) | 0.863 ( 0.012, 66.760) | 1.019 ( 0.256, 4.054) | 0.630 ( 0.009, 51.522) | 0.640 ( 0.146, 2.735) | 3.724 ( 0.511, 40.756) |
|  |  | CW_A_RY_B | 0.551 ( 0.052, 4.823) | 0.716 ( 0.067, 6.675) | 0.761 ( 0.008, 73.142) | 0.894 ( 0.075, 8.887) | 0.556 ( 0.006, 52.379) | 0.544 ( 0.048, 5.732) | 3.269 ( 0.193, 66.934) |
|  |  |  | CW_R_BII | 1.299 ( 0.399, 3.976) | 1.368 ( 0.021, 101.059) | 1.608 ( 0.499, 5.273) | 0.998 ( 0.014, 76.424) | 1.002 ( 0.248, 3.980) | 5.939 ( 0.851, 59.585) |
|  |  |  |  | PP_A_BII | 1.061 ( 0.020, 73.774) | 1.227 ( 0.534, 3.053) | 0.787 ( 0.013, 53.415) | 0.770 ( 0.364, 1.639) | 4.523 ( 0.824, 40.561) |
|  |  |  |  |  | PP_A_BII_B | 1.178 ( 0.019, 70.683) | 0.734 ( 0.295, 1.824) | 0.724 ( 0.009, 41.368) | 4.367 ( 0.047, 431.600) |
|  |  |  |  |  |  | PP_R_BII | 0.633 ( 0.010, 41.997) | 0.626 ( 0.198, 1.869) | 3.706 ( 0.669, 30.305) |
|  |  |  |  |  |  |  | PP_R_BII_B | 0.970 ( 0.012, 61.246) | 5.815 ( 0.062, 616.464) |
|  |  |  |  |  |  |  |  | PR_A_BII | 5.881 ( 1.063, 53.705) |
|  |  |  |  |  |  |  |  |  | PR_R_BII |

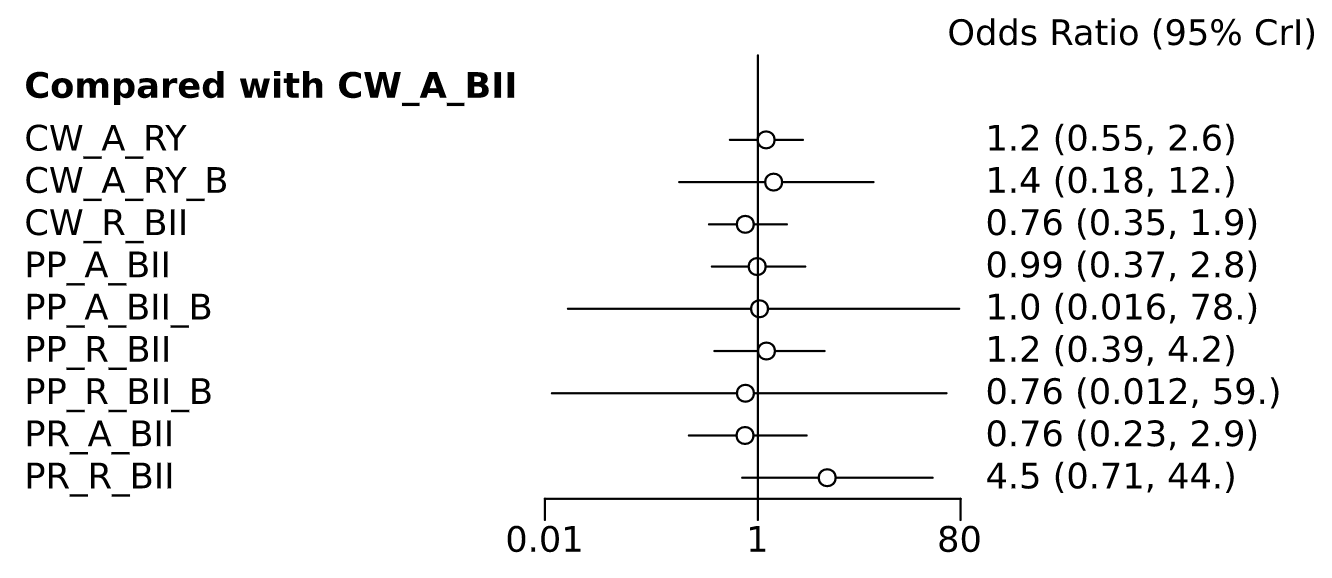

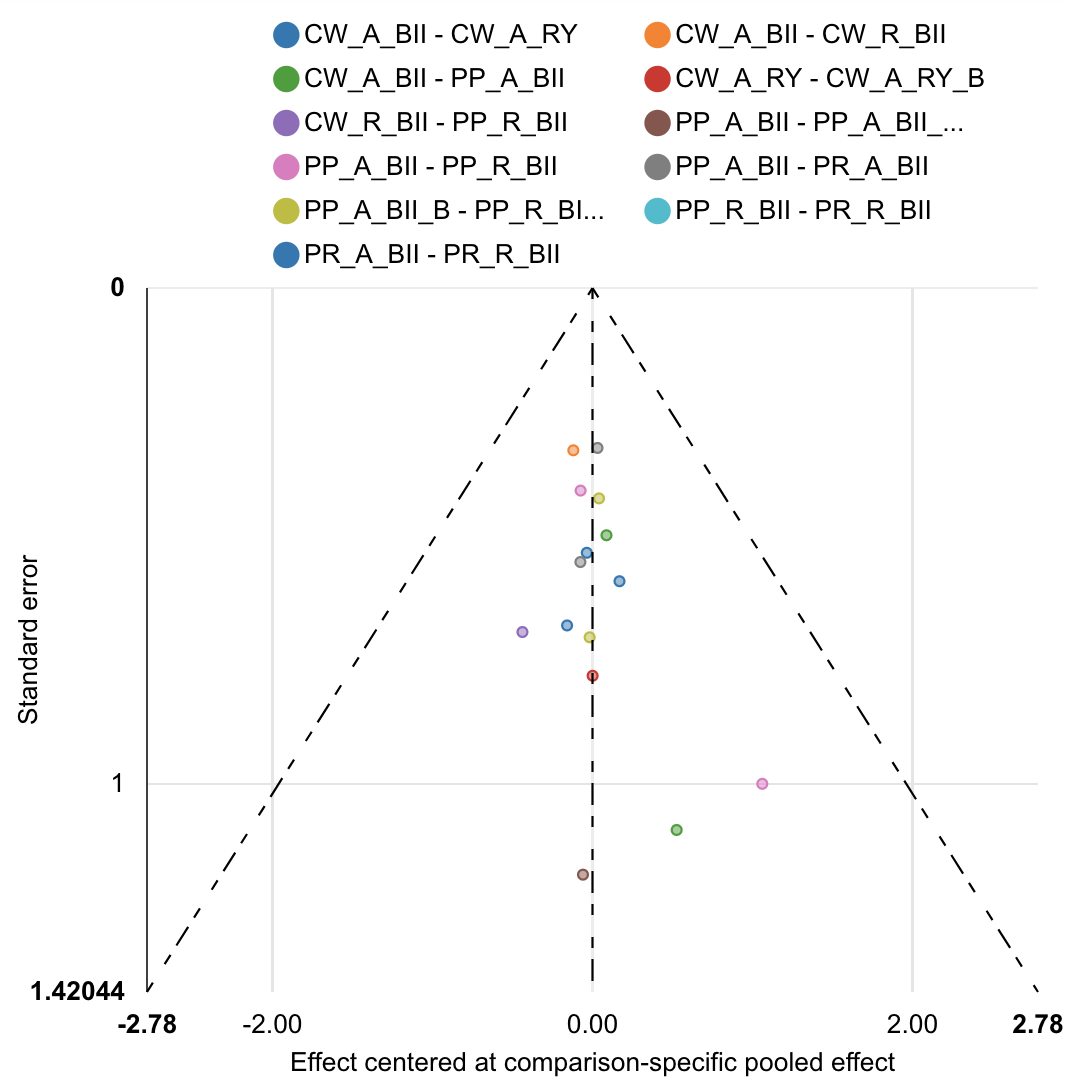

| Rank probabilities table | | | | | | | | | | |
| --- | --- | --- | --- | --- | --- | --- | --- | --- | --- | --- |
|  | **Rank 1** | **Rank 2** | **Rank 3** | **Rank 4** | **Rank 5** | **Rank 6** | **Rank 7** | **Rank 8** | **Rank 9** | **Rank 10** |
| CW_A_BII | 0.026 | 0.089 | 0.122 | 0.153 | 0.175 | 0.161 | 0.149 | 0.086 | 0.035 | 0.004 |
| CW_A_RY | 0.032 | 0.052 | 0.094 | 0.107 | 0.114 | 0.139 | 0.147 | 0.179 | 0.115 | 0.023 |
| CW_A_RY_B | 0.135 | 0.058 | 0.081 | 0.064 | 0.058 | 0.066 | 0.100 | 0.105 | 0.194 | 0.140 |
| CW_R_BII | 0.181 | 0.146 | 0.187 | 0.149 | 0.113 | 0.101 | 0.059 | 0.036 | 0.024 | 0.004 |
| PP_A_BII | 0.015 | 0.076 | 0.115 | 0.182 | 0.194 | 0.179 | 0.128 | 0.081 | 0.029 | 0.002 |
| PP_A_BII_B | 0.085 | 0.269 | 0.061 | 0.037 | 0.039 | 0.045 | 0.065 | 0.097 | 0.168 | 0.135 |
| PP_R_BII | 0.021 | 0.039 | 0.076 | 0.109 | 0.127 | 0.159 | 0.180 | 0.132 | 0.141 | 0.016 |
| PP_R_BII_B | 0.329 | 0.130 | 0.051 | 0.033 | 0.040 | 0.040 | 0.060 | 0.125 | 0.137 | 0.055 |
| PR_A_BII | 0.173 | 0.136 | 0.204 | 0.156 | 0.122 | 0.083 | 0.069 | 0.037 | 0.018 | 0.001 |
| PR_R_BII | 0.003 | 0.005 | 0.008 | 0.011 | 0.018 | 0.026 | 0.043 | 0.126 | 0.140 | 0.620 |

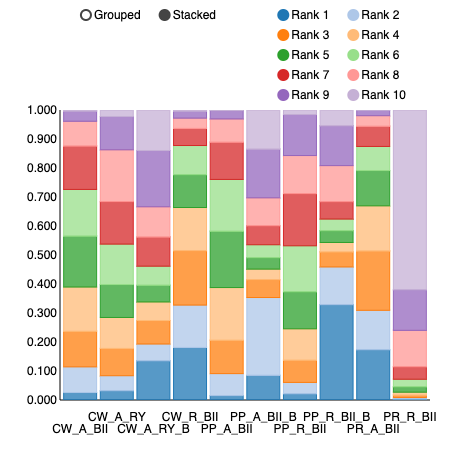

Wound infection

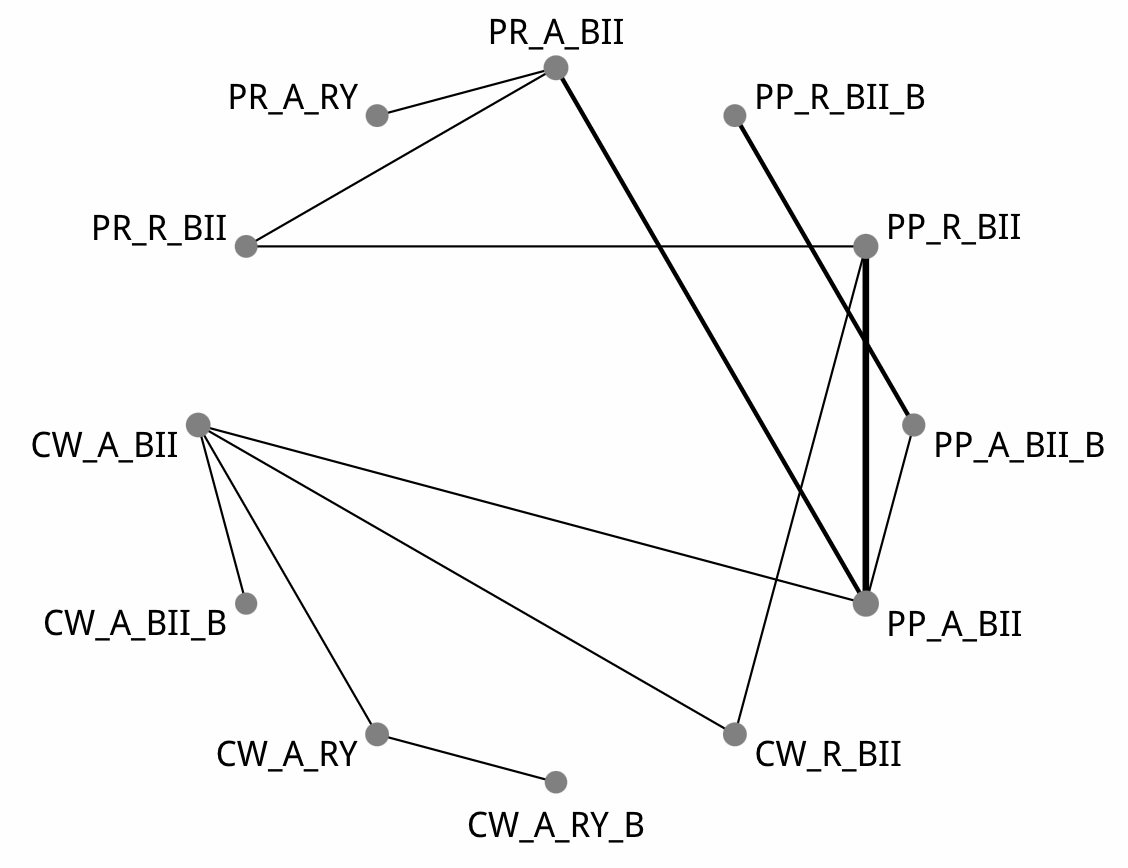

| Comparison of the included interventions: odds ratio (95% CrI). Each cell gives the effect of the column-defining intervention relative to the row-defining intervention. | | | | | | | | | | | |
| --- | --- | --- | --- | --- | --- | --- | --- | --- | --- | --- | --- |
| CW_A_BII | 2.472 ( 0.136, 108.429) | 0.815 ( 0.179, 3.745) | 0.707 ( 0.068, 7.089) | 0.838 ( 0.180, 3.638) | 0.850 ( 0.110, 7.034) | 2.848 ( 0.144, 85.183) | 0.707 ( 0.085, 5.154) | 2.732 ( 0.095, 97.719) | 0.588 ( 0.054, 7.189) | 0.201 ( 0.006, 5.225) | 0.572 ( 0.046, 7.553) |
|  | CW_A_BII_B | 0.324 ( 0.005, 8.996) | 0.295 ( 0.003, 12.309) | 0.333 ( 0.006, 8.453) | 0.337 ( 0.006, 12.014) | 1.122 ( 0.010, 80.375) | 0.292 ( 0.004, 8.549) | 1.036 ( 0.008, 95.756) | 0.229 ( 0.003, 10.221) | 0.079 ( 0.001, 6.515) | 0.230 ( 0.003, 9.546) |
|  |  | CW_A_RY | 0.885 ( 0.144, 5.157) | 1.031 ( 0.116, 9.249) | 1.039 ( 0.095, 14.517) | 3.482 ( 0.132, 149.965) | 0.868 ( 0.068, 10.532) | 3.383 ( 0.097, 170.835) | 0.734 ( 0.045, 12.816) | 0.251 ( 0.005, 9.761) | 0.701 ( 0.037, 14.559) |
|  |  |  | CW_A_RY_B | 1.190 ( 0.072, 17.315) | 1.197 ( 0.059, 30.997) | 3.967 ( 0.094, 245.476) | 0.998 ( 0.044, 19.269) | 3.805 ( 0.070, 303.142) | 0.839 ( 0.032, 25.747) | 0.283 ( 0.004, 17.609) | 0.808 ( 0.025, 26.525) |
|  |  |  |  | CW_R_BII | 1.018 ( 0.164, 7.215) | 3.406 ( 0.206, 82.401) | 0.843 ( 0.144, 4.434) | 3.318 ( 0.139, 109.093) | 0.699 ( 0.077, 7.089) | 0.243 ( 0.008, 5.438) | 0.677 ( 0.065, 7.128) |
|  |  |  |  |  | PP_A_BII | 3.278 ( 0.367, 39.666) | 0.846 ( 0.247, 2.017) | 3.176 ( 0.245, 50.978) | 0.690 ( 0.173, 2.545) | 0.242 ( 0.013, 2.723) | 0.691 ( 0.113, 3.481) |
|  |  |  |  |  |  | PP_A_BII_B | 0.246 ( 0.016, 2.615) | 0.970 ( 0.250, 3.533) | 0.210 ( 0.012, 2.643) | 0.071 ( 0.002, 1.940) | 0.202 ( 0.010, 3.045) |
|  |  |  |  |  |  |  | PP_R_BII | 3.867 ( 0.263, 77.285) | 0.847 ( 0.193, 4.370) | 0.293 ( 0.016, 4.157) | 0.823 ( 0.168, 4.207) |
|  |  |  |  |  |  |  |  | PP_R_BII_B | 0.216 ( 0.010, 3.885) | 0.074 ( 0.001, 2.602) | 0.212 ( 0.008, 4.116) |
|  |  |  |  |  |  |  |  |  | PR_A_BII | 0.349 ( 0.026, 2.835) | 0.980 ( 0.169, 5.075) |
|  |  |  |  |  |  |  |  |  |  | PR_A_RY | 2.900 ( 0.189, 56.724) |
|  |  |  |  |  |  |  |  |  |  |  | PR_R_BII |

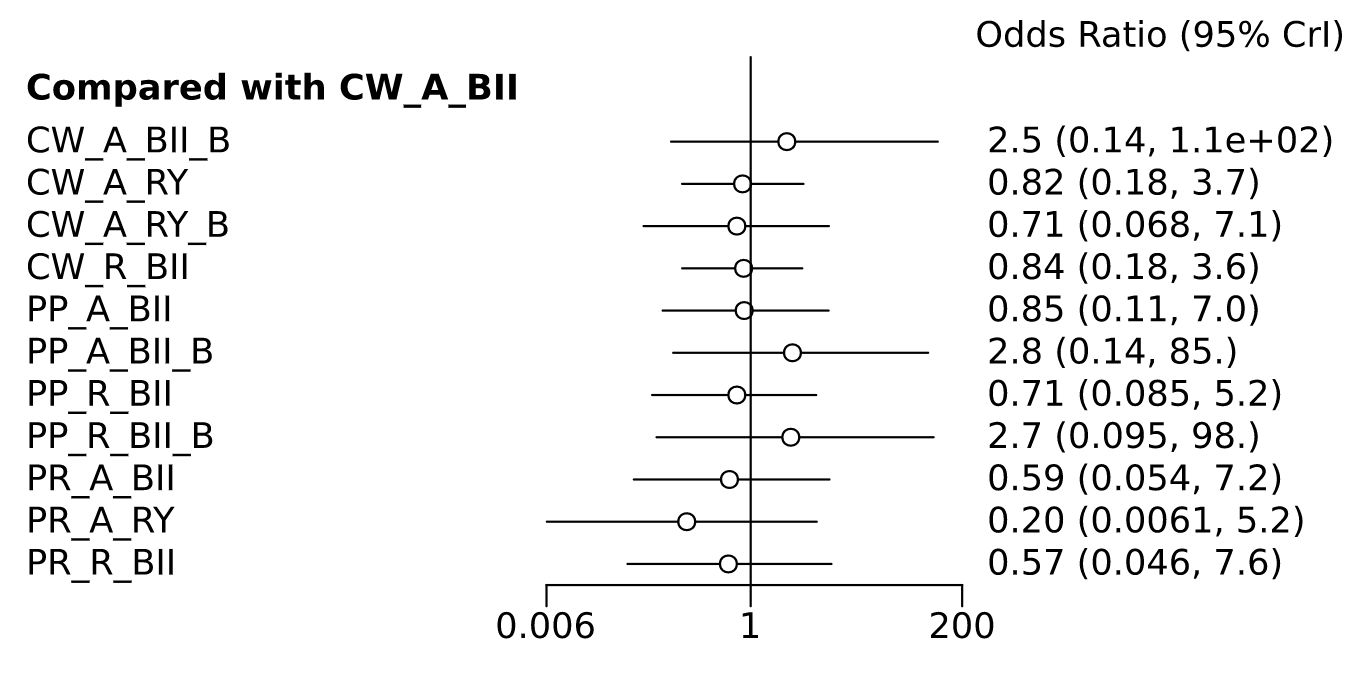

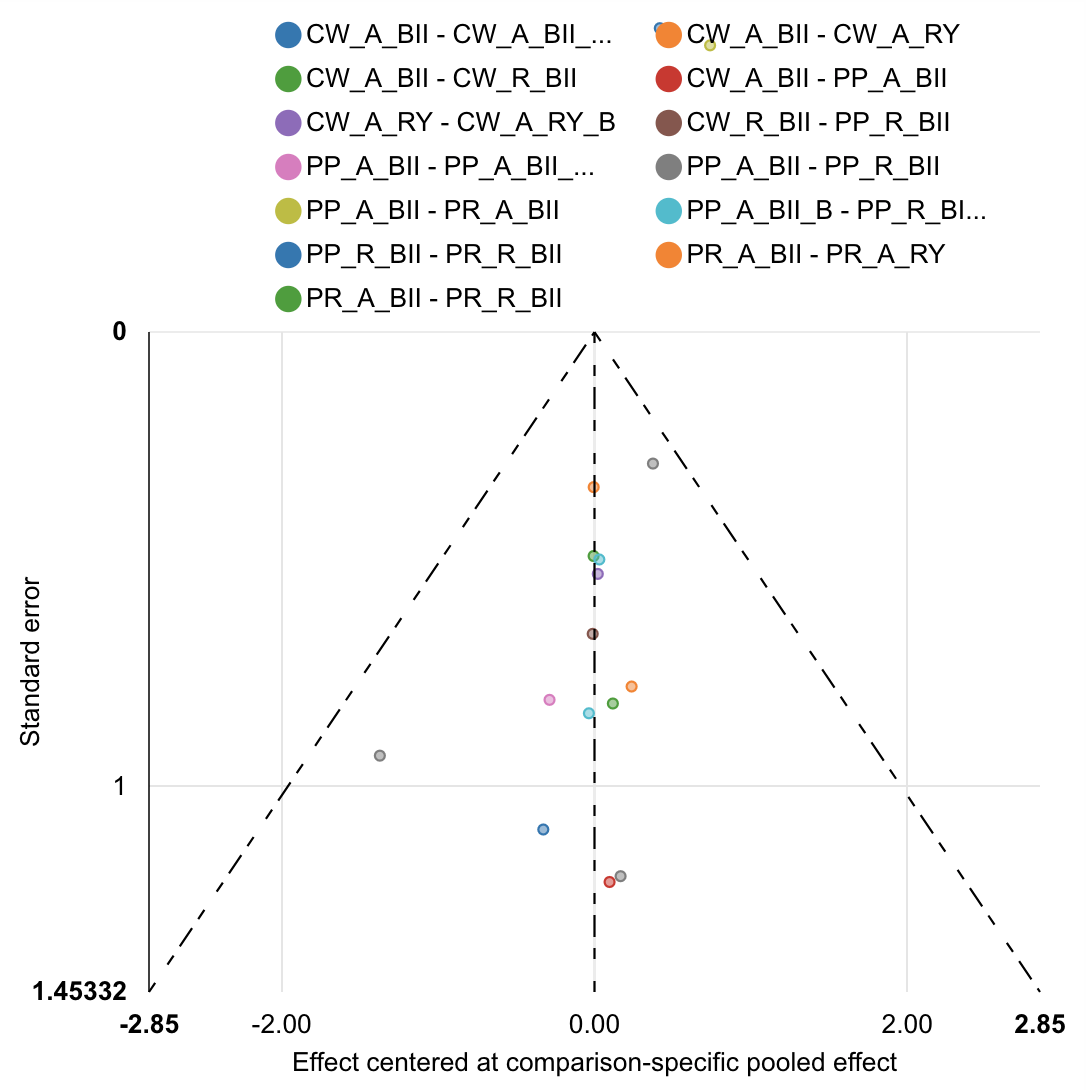

| Rank probabilities table | | | | | | | | | | | | |
| --- | --- | --- | --- | --- | --- | --- | --- | --- | --- | --- | --- | --- |
|  | **Rank 1** | **Rank 2** | **Rank 3** | **Rank 4** | **Rank 5** | **Rank 6** | **Rank 7** | **Rank 8** | **Rank 9** | **Rank 10** | **Rank 11** | **Rank 12** |
| CW_A_BII | 0.010 | 0.030 | 0.069 | 0.089 | 0.098 | 0.094 | 0.123 | 0.143 | 0.152 | 0.101 | 0.072 | 0.020 |
| CW_A_BII_B | 0.048 | 0.050 | 0.038 | 0.034 | 0.036 | 0.039 | 0.040 | 0.043 | 0.063 | 0.155 | 0.085 | 0.369 |
| CW_A_RY | 0.042 | 0.106 | 0.113 | 0.092 | 0.084 | 0.089 | 0.096 | 0.107 | 0.104 | 0.092 | 0.053 | 0.023 |
| CW_A_RY_B | 0.140 | 0.136 | 0.088 | 0.078 | 0.061 | 0.069 | 0.071 | 0.070 | 0.089 | 0.076 | 0.072 | 0.052 |
| CW_R_BII | 0.027 | 0.062 | 0.082 | 0.106 | 0.115 | 0.135 | 0.117 | 0.121 | 0.105 | 0.071 | 0.040 | 0.019 |
| PP_A_BII | 0.006 | 0.026 | 0.060 | 0.115 | 0.153 | 0.140 | 0.135 | 0.138 | 0.122 | 0.078 | 0.019 | 0.009 |
| PP_A_BII_B | 0.006 | 0.018 | 0.022 | 0.025 | 0.029 | 0.041 | 0.052 | 0.055 | 0.068 | 0.151 | 0.311 | 0.225 |
| PP_R_BII | 0.020 | 0.069 | 0.113 | 0.156 | 0.153 | 0.143 | 0.119 | 0.104 | 0.075 | 0.032 | 0.014 | 0.003 |
| PP_R_BII_B | 0.020 | 0.030 | 0.030 | 0.026 | 0.028 | 0.038 | 0.051 | 0.057 | 0.063 | 0.134 | 0.277 | 0.247 |
| PR_A_BII | 0.027 | 0.184 | 0.162 | 0.124 | 0.109 | 0.093 | 0.086 | 0.077 | 0.068 | 0.039 | 0.020 | 0.010 |
| PR_A_RY | 0.568 | 0.119 | 0.074 | 0.052 | 0.044 | 0.031 | 0.027 | 0.022 | 0.025 | 0.021 | 0.011 | 0.009 |
| PR_R_BII | 0.087 | 0.171 | 0.151 | 0.105 | 0.091 | 0.088 | 0.085 | 0.064 | 0.068 | 0.049 | 0.028 | 0.015 |

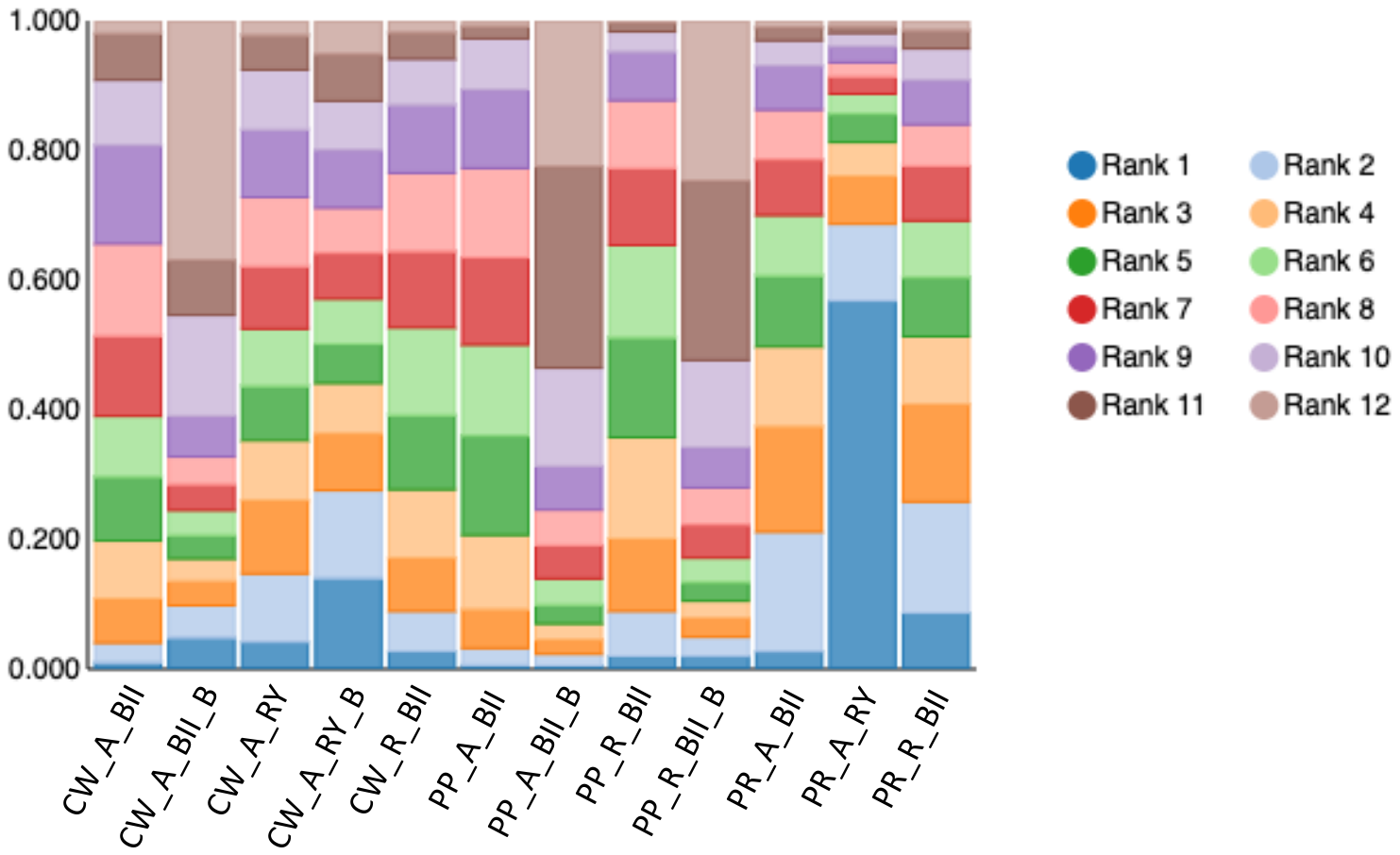

Bile leak

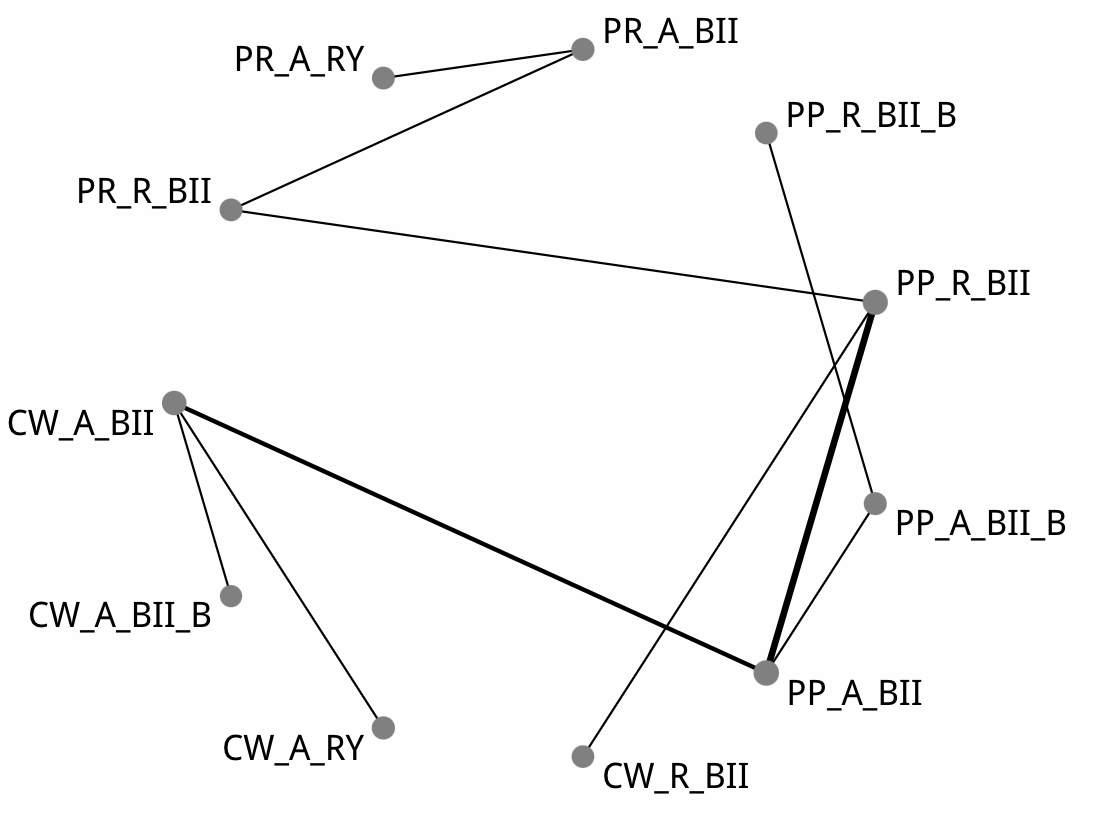

| Comparison of the included interventions: odds ratio (95% CrI). Each cell gives the effect of the column-defining intervention relative to the row-defining intervention. | | | | | | | | | | |
| --- | --- | --- | --- | --- | --- | --- | --- | --- | --- | --- |
| CW_A_BII | 0.393 ( 0.011, 6.624) | 0.629 ( 0.055, 5.684) | 4.021 ( 0.158, 105.330) | 2.114 ( 0.295, 19.846) | 0.846 ( 0.016, 28.310) | 3.906 ( 0.372, 48.482) | 0.319 ( 0.002, 30.750) | 8.033 ( 0.059, 1,405.854) | 2.774 ( 0.007, 1,210.635) | 23.203 ( 0.695, 1,943.605) |
|  | CW_A_BII_B | 1.607 ( 0.038, 95.603) | 10.898 ( 0.149, 1,231.761) | 5.679 ( 0.166, 331.193) | 2.276 ( 0.015, 310.443) | 10.964 ( 0.252, 761.584) | 0.851 ( 0.003, 231.296) | 23.580 ( 0.072, 9,713.772) | 8.156 ( 0.010, 7,755.829) | 66.873 ( 0.567, 14,407.440) |
|  |  | CW_A_RY | 6.782 ( 0.121, 337.107) | 3.603 ( 0.186, 80.302) | 1.403 ( 0.015, 97.719) | 6.411 ( 0.270, 185.286) | 0.526 ( 0.002, 99.883) | 13.293 ( 0.048, 4,283.818) | 4.468 ( 0.008, 3,379.219) | 39.833 ( 0.532, 6,049.313) |
|  |  |  | CW_R_BII | 0.541 ( 0.046, 7.273) | 0.211 ( 0.003, 10.606) | 1.004 ( 0.124, 9.781) | 0.080 ( 0.000, 10.109) | 2.082 ( 0.018, 316.303) | 0.705 ( 0.002, 240.207) | 5.646 ( 0.198, 389.553) |
|  |  |  |  | PP_A_BII | 0.399 ( 0.012, 6.745) | 1.867 ( 0.516, 6.881) | 0.147 ( 0.001, 9.233) | 3.618 ( 0.045, 423.943) | 1.228 ( 0.005, 388.192) | 10.046 ( 0.637, 547.137) |
|  |  |  |  |  | PP_A_BII_B | 4.712 ( 0.205, 198.106) | 0.415 ( 0.009, 7.063) | 9.824 ( 0.053, 3,885.862) | 3.314 ( 0.007, 3,094.247) | 28.076 ( 0.457, 5,466.534) |
|  |  |  |  |  |  | PP_R_BII | 0.078 ( 0.001, 5.880) | 1.938 ( 0.030, 186.345) | 0.689 ( 0.004, 165.753) | 5.294 ( 0.465, 218.416) |
|  |  |  |  |  |  |  | PP_R_BII_B | 26.340 ( 0.072, 20,694.019) | 8.925 ( 0.011, 12,637.198) | 73.744 ( 0.455, 27,092.169) |
|  |  |  |  |  |  |  |  | PR_A_BII | 0.362 ( 0.010, 6.012) | 2.836 ( 0.146, 86.965) |
|  |  |  |  |  |  |  |  |  | PR_A_RY | 8.331 ( 0.132, 985.944) |
|  |  |  |  |  |  |  |  |  |  | PR_R_BII |

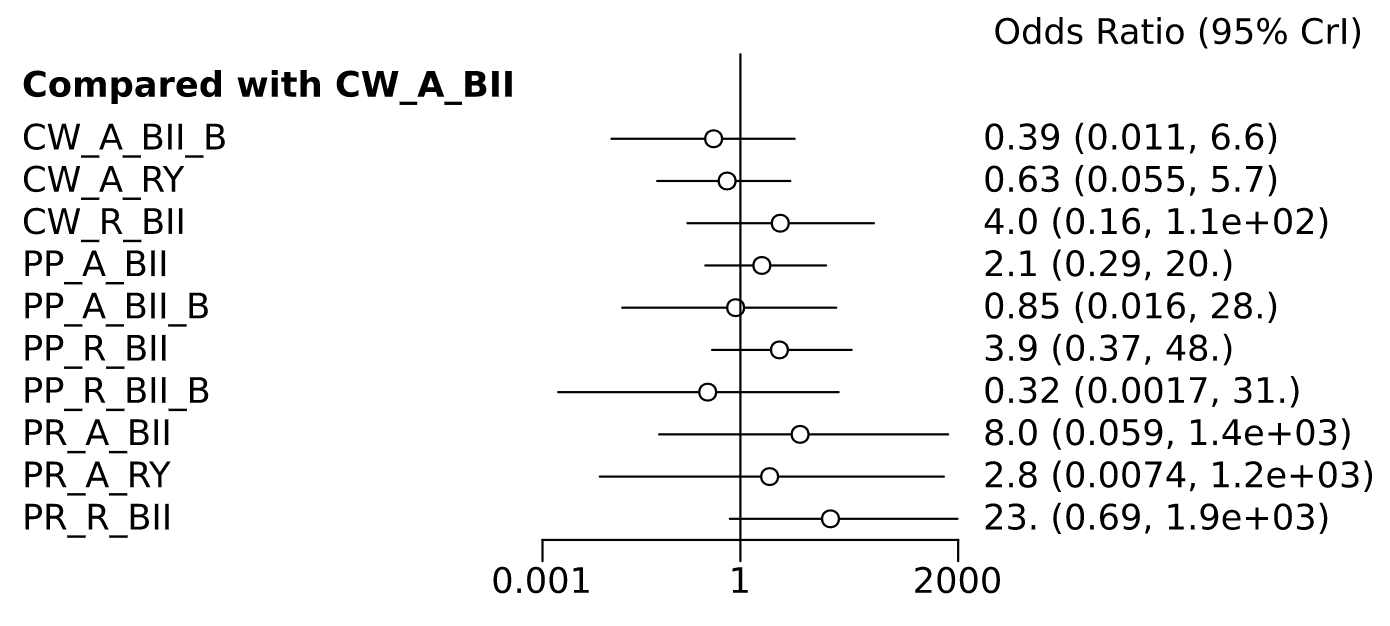

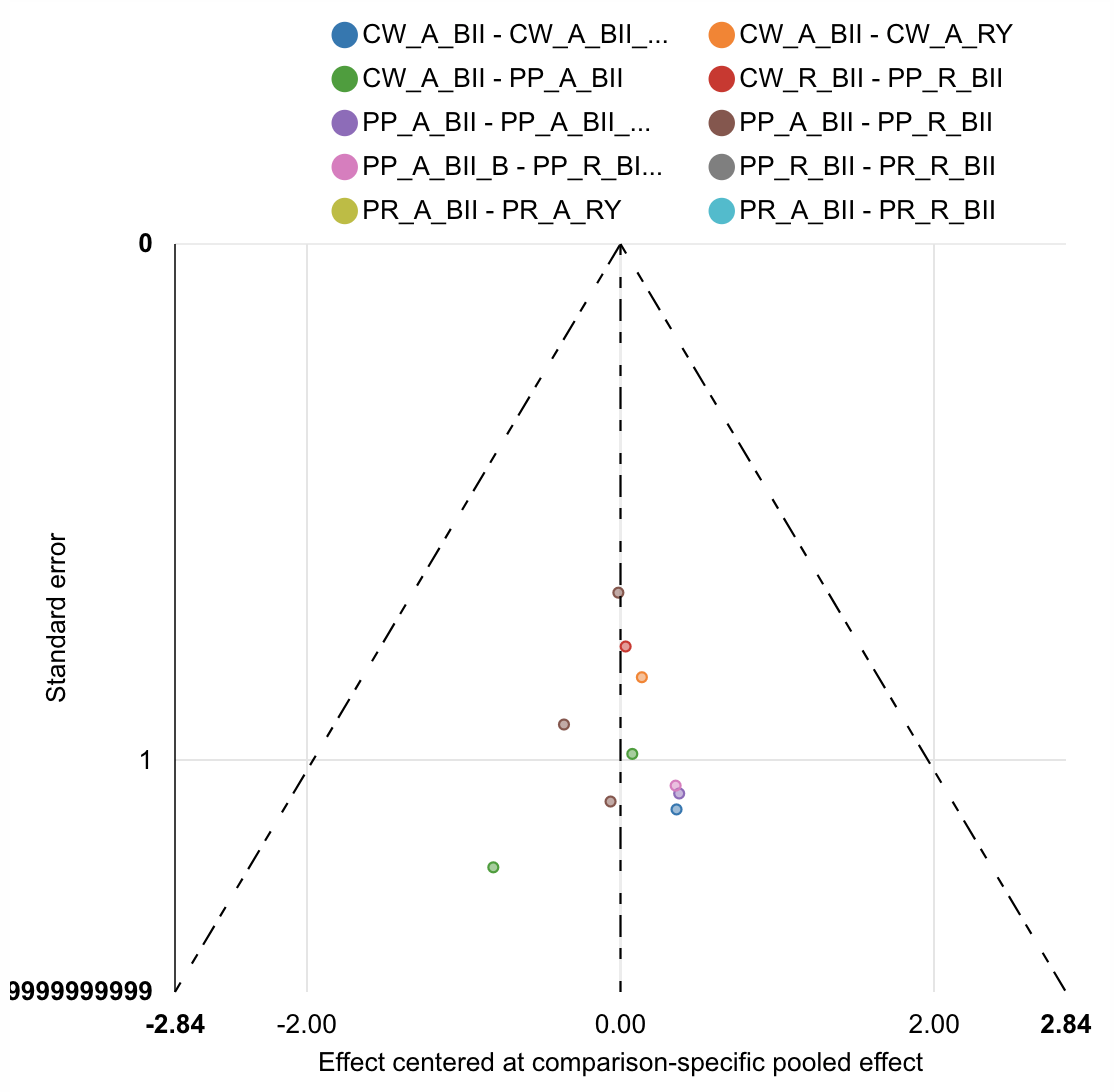

| Rank probabilities table | | | | | | | | | | | |
| --- | --- | --- | --- | --- | --- | --- | --- | --- | --- | --- | --- |
|  | **Rank 1** | **Rank 2** | **Rank 3** | **Rank 4** | **Rank 5** | **Rank 6** | **Rank 7** | **Rank 8** | **Rank 9** | **Rank 10** | **Rank 11** |
| CW_A_BII | 0.016 | 0.086 | 0.184 | 0.201 | 0.211 | 0.132 | 0.073 | 0.047 | 0.028 | 0.014 | 0.008 |
| CW_A_BII_B | 0.285 | 0.187 | 0.162 | 0.105 | 0.073 | 0.051 | 0.041 | 0.035 | 0.021 | 0.021 | 0.018 |
| CW_A_RY | 0.138 | 0.199 | 0.175 | 0.148 | 0.102 | 0.067 | 0.053 | 0.046 | 0.031 | 0.022 | 0.018 |
| CW_R_BII | 0.013 | 0.027 | 0.040 | 0.062 | 0.080 | 0.096 | 0.148 | 0.171 | 0.132 | 0.130 | 0.102 |
| PP_A_BII | 0.002 | 0.010 | 0.035 | 0.103 | 0.160 | 0.271 | 0.209 | 0.133 | 0.052 | 0.021 | 0.004 |
| PP_A_BII_B | 0.057 | 0.193 | 0.161 | 0.130 | 0.130 | 0.091 | 0.069 | 0.060 | 0.042 | 0.046 | 0.021 |
| PP_R_BII | 0.000 | 0.004 | 0.008 | 0.023 | 0.054 | 0.110 | 0.217 | 0.273 | 0.194 | 0.100 | 0.018 |
| PP_R_BII_B | 0.350 | 0.169 | 0.110 | 0.094 | 0.073 | 0.048 | 0.037 | 0.038 | 0.030 | 0.026 | 0.024 |
| PR_A_BII | 0.011 | 0.045 | 0.046 | 0.052 | 0.050 | 0.057 | 0.062 | 0.080 | 0.145 | 0.301 | 0.153 |
| PR_A_RY | 0.128 | 0.080 | 0.077 | 0.074 | 0.058 | 0.063 | 0.064 | 0.069 | 0.179 | 0.115 | 0.095 |
| PR_R_BII | 0.001 | 0.000 | 0.002 | 0.007 | 0.008 | 0.015 | 0.028 | 0.048 | 0.146 | 0.206 | 0.540 |

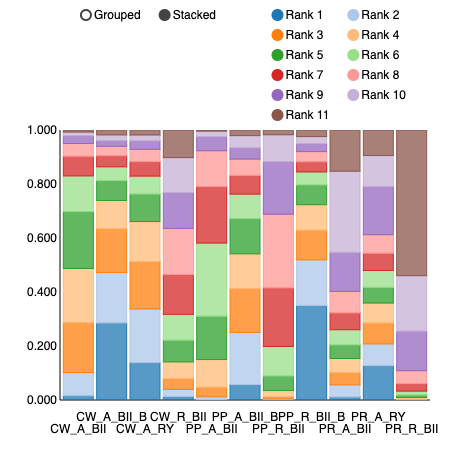

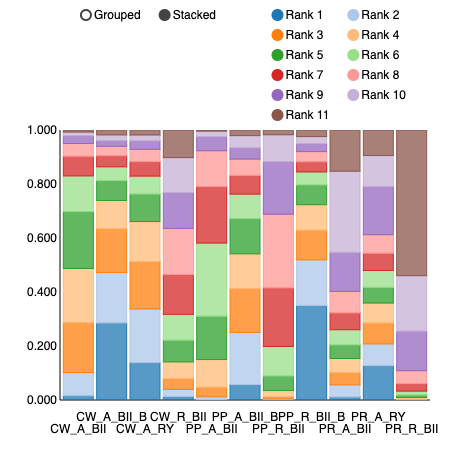

Reoperation

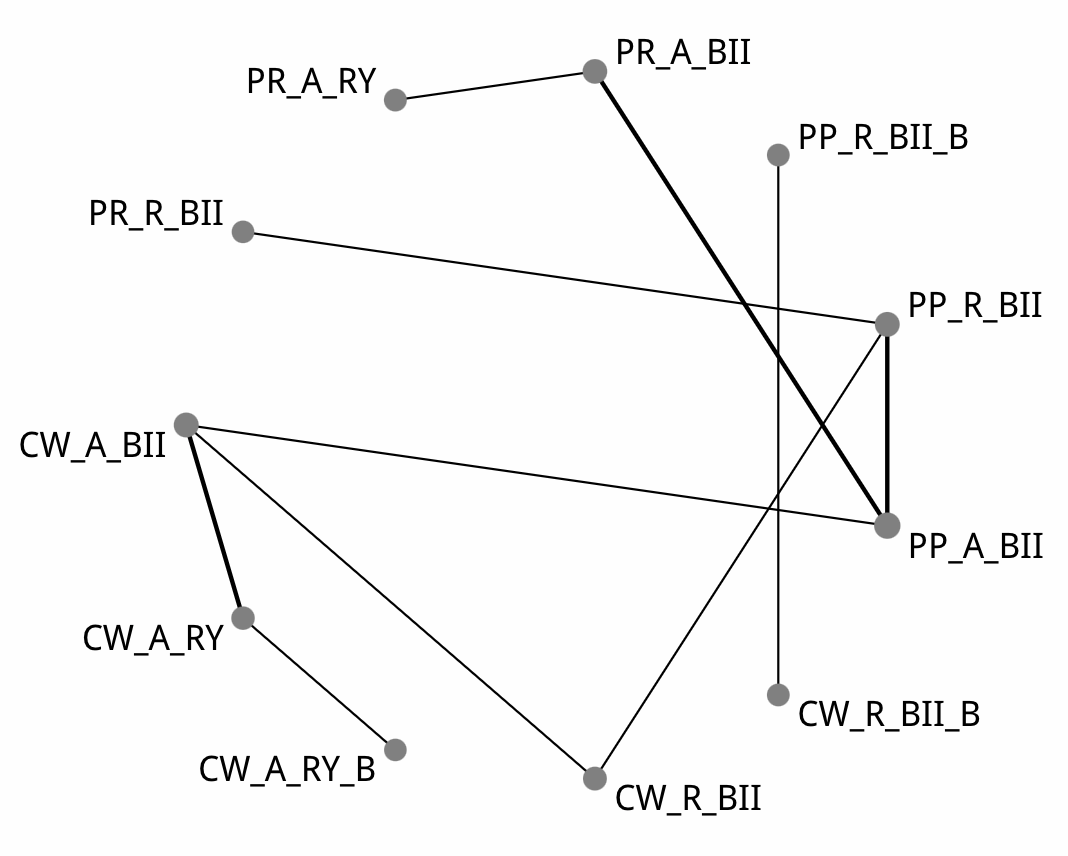

There were no studies other than Seiler et al. 2005 that reported reoperation rates that also compared PP_R_BII_B or CW_R_BII_B; hence this comparison had to be removed. The OR in Seiler et al. 2005 for reoperation was 0.48 (95% CI 0.043 – 5.48) favouring CW_R_BII_B. The following results exclude this from the network map.

| Comparison of the included interventions: odds ratio (95% CrI). Each cell gives the effect of the column-defining intervention relative to the row-defining intervention. | | | | | | | | |
| --- | --- | --- | --- | --- | --- | --- | --- | --- |
| CW_A_BII | 0.524 ( 0.118, 2.204) | 0.262 ( 0.006, 5.908) | 1.031 ( 0.264, 4.134) | 0.713 ( 0.236, 2.066) | 1.138 ( 0.283, 4.468) | 0.596 ( 0.130, 2.834) | 0.154 ( 0.004, 2.462) | 1.031 ( 0.026, 47.196) |
|  | CW_A_RY | 0.513 ( 0.016, 8.545) | 1.959 ( 0.273, 14.721) | 1.345 ( 0.222, 8.458) | 2.186 ( 0.315, 16.341) | 1.128 ( 0.146, 9.974) | 0.282 ( 0.007, 6.201) | 1.986 ( 0.039, 125.826) |
|  |  | CW_A_RY_B | 3.957 ( 0.124, 184.491) | 2.681 ( 0.102, 128.047) | 4.307 ( 0.146, 226.331) | 2.279 ( 0.075, 121.425) | 0.562 ( 0.006, 64.360) | 3.887 ( 0.029, 696.940) |
|  |  |  | CW_R_BII | 0.689 ( 0.168, 2.859) | 1.116 ( 0.297, 4.307) | 0.582 ( 0.094, 3.460) | 0.141 ( 0.004, 3.039) | 1.011 ( 0.025, 43.506) |
|  |  |  |  | PP_A_BII | 1.596 ( 0.595, 4.665) | 0.839 ( 0.282, 2.555) | 0.218 ( 0.007, 2.756) | 1.463 ( 0.041, 61.492) |
|  |  |  |  |  | PP_R_BII | 0.529 ( 0.111, 2.270) | 0.133 ( 0.004, 2.059) | 0.892 ( 0.028, 32.967) |
|  |  |  |  |  |  | PR_A_BII | 0.256 ( 0.010, 2.661) | 1.748 ( 0.042, 85.962) |
|  |  |  |  |  |  |  | PR_A_RY | 7.263 ( 0.079, 1,114.209) |
|  |  |  |  |  |  |  |  | PR_R_BII |

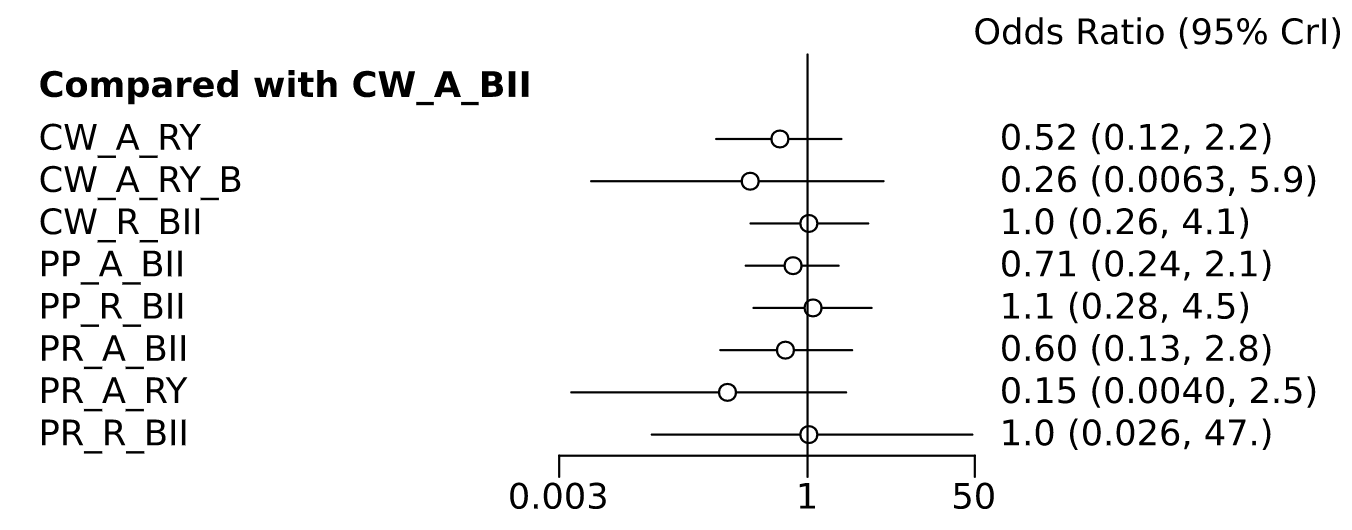

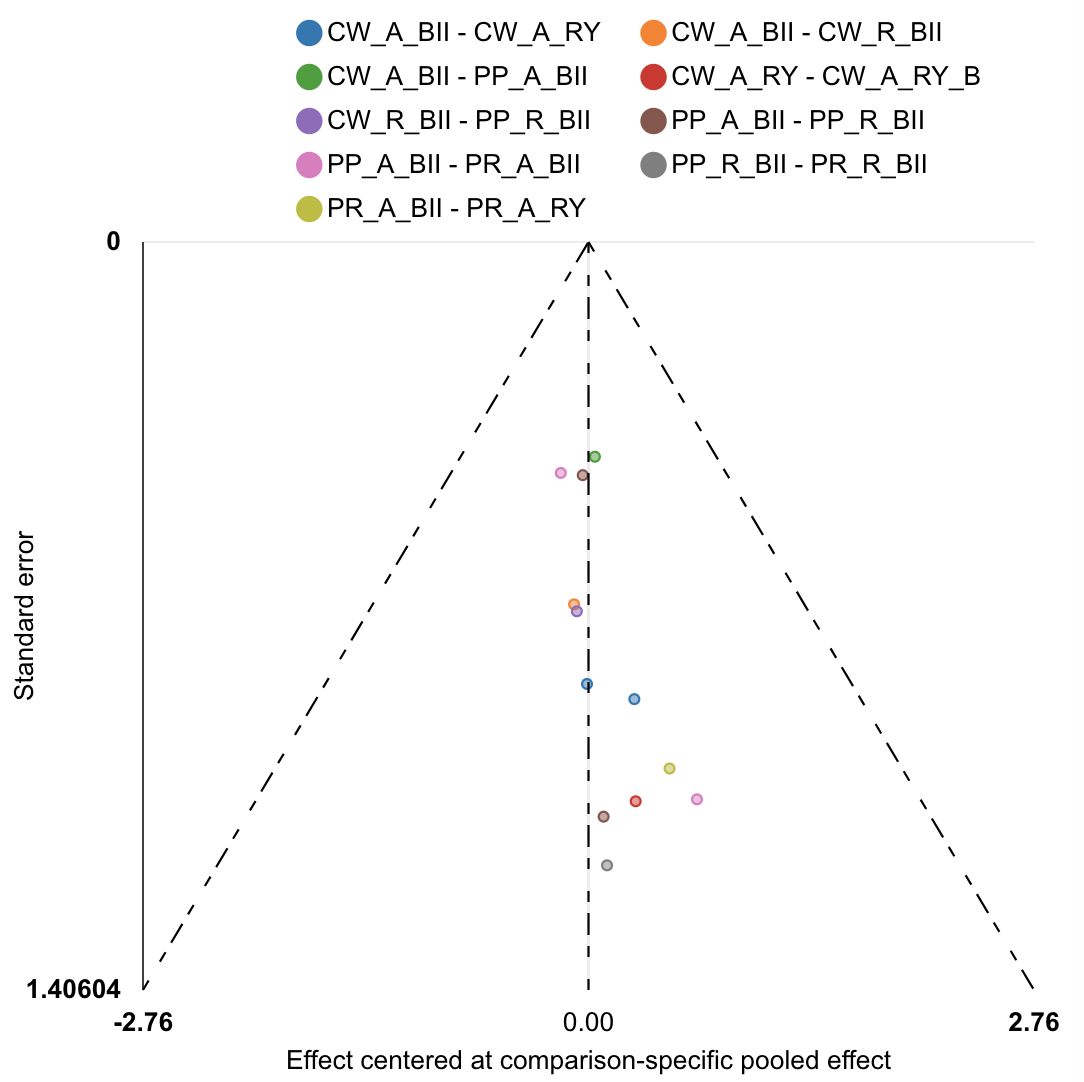

| Rank probabilities table | | | | | | | | | |
| --- | --- | --- | --- | --- | --- | --- | --- | --- | --- |
|  | **Rank 1** | **Rank 2** | **Rank 3** | **Rank 4** | **Rank 5** | **Rank 6** | **Rank 7** | **Rank 8** | **Rank 9** |
| CW_A_BII | 0.001 | 0.012 | 0.041 | 0.097 | 0.139 | 0.202 | 0.206 | 0.193 | 0.108 |
| CW_A_RY | 0.041 | 0.186 | 0.234 | 0.151 | 0.104 | 0.086 | 0.080 | 0.075 | 0.043 |
| CW_A_RY_B | 0.332 | 0.225 | 0.098 | 0.057 | 0.052 | 0.046 | 0.042 | 0.058 | 0.091 |
| CW_R_BII | 0.010 | 0.037 | 0.070 | 0.101 | 0.117 | 0.132 | 0.178 | 0.190 | 0.166 |
| PP_A_BII | 0.006 | 0.041 | 0.124 | 0.221 | 0.252 | 0.196 | 0.103 | 0.046 | 0.012 |
| PP_R_BII | 0.002 | 0.011 | 0.032 | 0.057 | 0.102 | 0.155 | 0.223 | 0.266 | 0.153 |
| PR_A_BII | 0.019 | 0.141 | 0.201 | 0.198 | 0.152 | 0.107 | 0.085 | 0.062 | 0.037 |
| PR_A_RY | 0.484 | 0.230 | 0.107 | 0.048 | 0.033 | 0.026 | 0.023 | 0.023 | 0.026 |
| PR_R_BII | 0.106 | 0.118 | 0.094 | 0.071 | 0.049 | 0.050 | 0.062 | 0.086 | 0.365 |

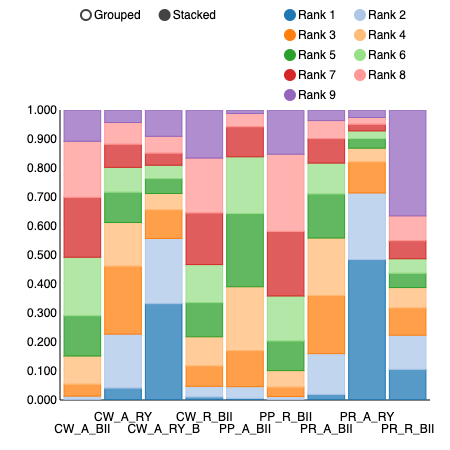

Mortality

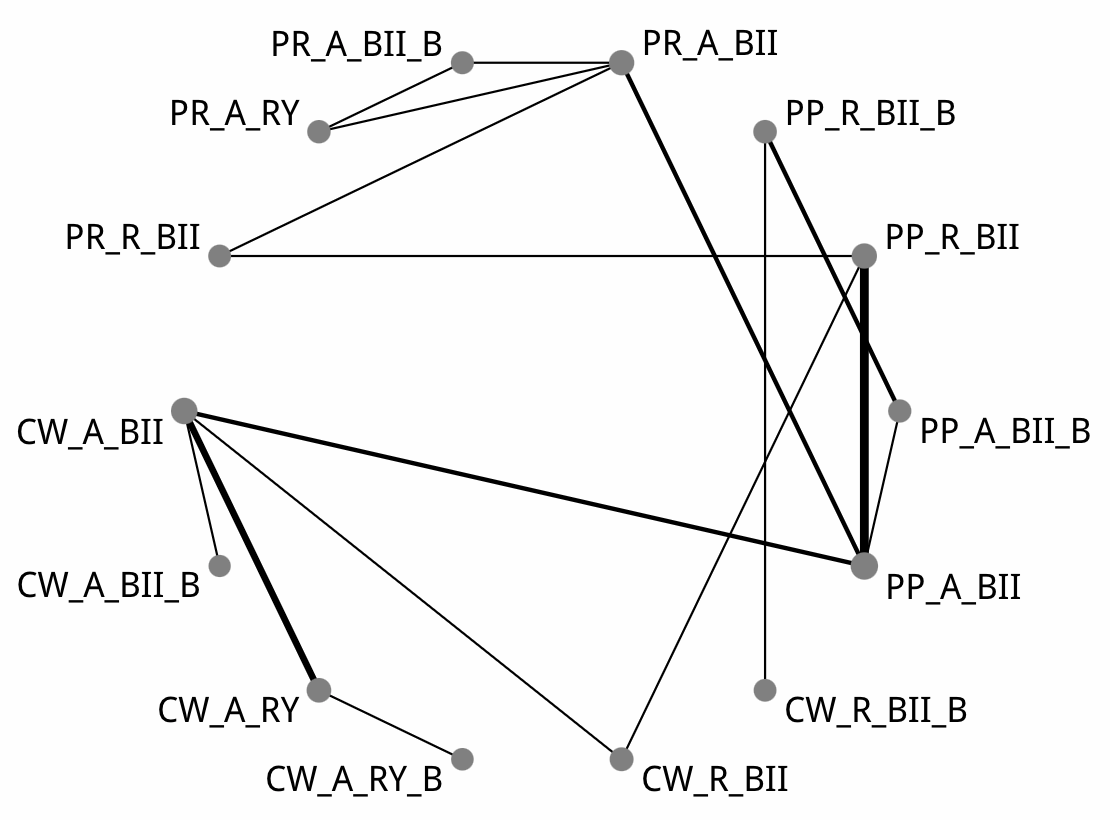

| Comparison of the included interventions: odds ratio (95% CrI). Each cell gives the effect of the column-defining intervention relative to the row-defining intervention. | | | | | | | | | | | | | |
| --- | --- | --- | --- | --- | --- | --- | --- | --- | --- | --- | --- | --- | --- |
| CW_A_BII | 0.387 ( 0.014, 6.048) | 2.596 ( 0.641, 13.217) | 1.998 ( 0.134, 25.977) | 0.752 ( 0.088, 6.018) | 1.557 ( 0.009, 412.320) | 0.692 ( 0.189, 2.564) | 0.672 ( 0.012, 27.941) | 1.143 ( 0.232, 5.924) | 0.675 ( 0.007, 66.023) | 0.726 ( 0.098, 6.320) | 0.708 ( 0.027, 27.077) | 0.742 ( 0.023, 22.200) | 0.815 ( 0.050, 15.499) |
|  | CW_A_BII_B | 6.848 ( 0.303, 282.083) | 5.440 ( 0.098, 347.443) | 2.057 ( 0.067, 76.577) | 4.296 ( 0.011, 2,413.420) | 1.866 ( 0.090, 60.892) | 1.823 ( 0.012, 255.775) | 2.998 ( 0.129, 102.709) | 1.925 ( 0.007, 387.959) | 1.951 ( 0.066, 90.026) | 2.044 ( 0.024, 225.698) | 1.981 ( 0.021, 201.724) | 2.273 ( 0.045, 190.776) |
|  |  | CW_A_RY | 0.743 ( 0.078, 5.930) | 0.283 ( 0.019, 3.587) | 0.570 ( 0.002, 167.972) | 0.266 ( 0.033, 1.823) | 0.239 ( 0.003, 14.356) | 0.416 ( 0.047, 4.069) | 0.241 ( 0.002, 26.555) | 0.276 ( 0.023, 3.448) | 0.272 ( 0.008, 10.546) | 0.282 ( 0.006, 11.504) | 0.323 ( 0.014, 7.554) |
|  |  |  | CW_A_RY_B | 0.375 ( 0.012, 12.642) | 0.802 ( 0.002, 406.954) | 0.347 ( 0.020, 7.380) | 0.330 ( 0.003, 37.978) | 0.561 ( 0.028, 15.484) | 0.335 ( 0.002, 76.677) | 0.371 ( 0.014, 11.050) | 0.392 ( 0.007, 25.585) | 0.381 ( 0.005, 25.992) | 0.426 ( 0.011, 19.648) |
|  |  |  |  | CW_R_BII | 2.128 ( 0.009, 602.266) | 0.904 ( 0.128, 6.982) | 0.908 ( 0.014, 41.368) | 1.487 ( 0.262, 9.970) | 0.879 ( 0.008, 98.623) | 1.004 ( 0.080, 11.958) | 0.986 ( 0.026, 39.338) | 0.983 ( 0.022, 38.333) | 1.160 ( 0.054, 25.222) |
|  |  |  |  |  | CW_R_BII_B | 0.456 ( 0.002, 70.640) | 0.439 ( 0.009, 13.185) | 0.747 ( 0.003, 139.087) | 0.460 ( 0.014, 6.826) | 0.461 ( 0.001, 102.935) | 0.518 ( 0.001, 215.810) | 0.476 ( 0.001, 196.409) | 0.553 ( 0.002, 194.358) |
|  |  |  |  |  |  | PP_A_BII | 0.960 ( 0.023, 27.040) | 1.612 ( 0.610, 4.532) | 0.922 ( 0.011, 66.780) | 1.041 ( 0.232, 5.256) | 1.050 ( 0.052, 24.354) | 1.065 ( 0.042, 22.247) | 1.227 ( 0.100, 16.000) |
|  |  |  |  |  |  |  | PP_A_BII_B | 1.674 ( 0.053, 76.907) | 1.002 ( 0.110, 10.036) | 1.086 ( 0.025, 71.300) | 1.203 ( 0.011, 148.487) | 1.076 ( 0.012, 167.151) | 1.286 ( 0.022, 122.633) |
|  |  |  |  |  |  |  |  | PP_R_BII | 0.572 ( 0.006, 45.404) | 0.647 ( 0.115, 3.956) | 0.661 ( 0.028, 17.141) | 0.658 ( 0.023, 14.910) | 0.745 ( 0.063, 8.947) |
|  |  |  |  |  |  |  |  |  | PP_R_BII_B | 1.122 ( 0.011, 128.471) | 1.199 ( 0.006, 227.534) | 1.127 ( 0.006, 237.983) | 1.245 ( 0.011, 216.459) |
|  |  |  |  |  |  |  |  |  |  | PR_A_BII | 1.010 ( 0.070, 17.281) | 0.995 ( 0.062, 13.670) | 1.154 ( 0.096, 15.307) |
|  |  |  |  |  |  |  |  |  |  |  | PR_A_BII_B | 0.983 ( 0.060, 16.514) | 1.109 ( 0.029, 45.119) |
|  |  |  |  |  |  |  |  |  |  |  |  | PR_A_RY | 1.149 ( 0.031, 50.014) |
|  |  |  |  |  |  |  |  |  |  |  |  |  | PR_R_BII |

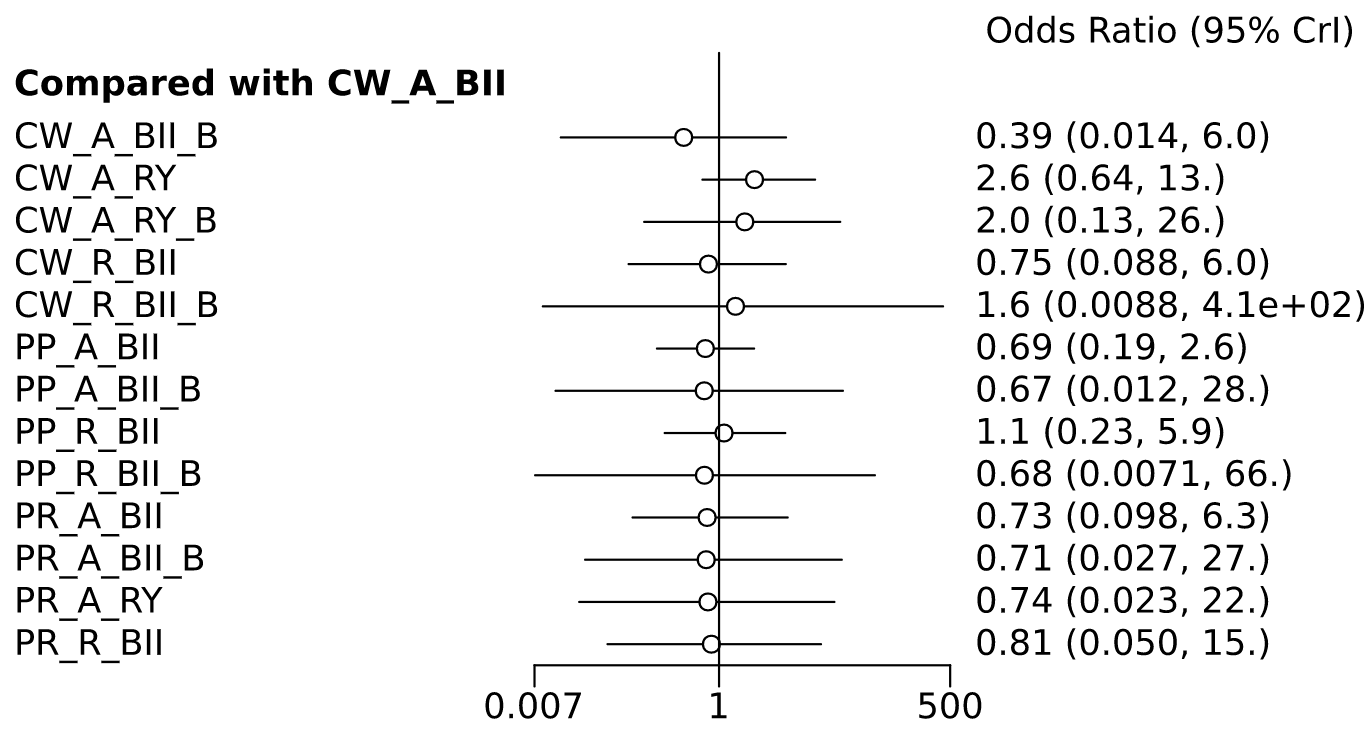

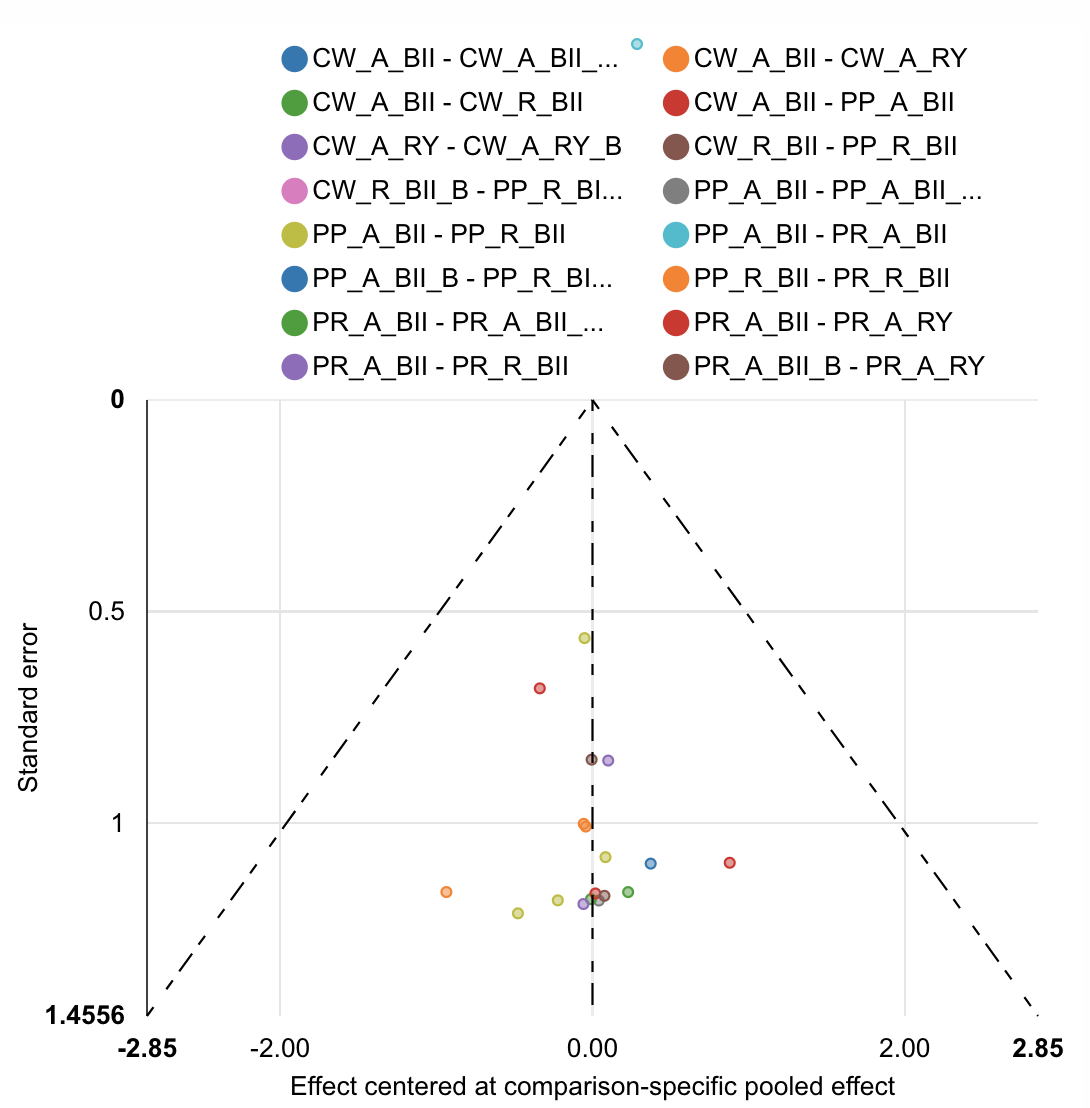

| Rank probabilities table | | | | | | | | | | | | | | |
| --- | --- | --- | --- | --- | --- | --- | --- | --- | --- | --- | --- | --- | --- | --- |
|  | **Rank 1** | **Rank 2** | **Rank 3** | **Rank 4** | **Rank 5** | **Rank 6** | **Rank 7** | **Rank 8** | **Rank 9** | **Rank 10** | **Rank 11** | **Rank 12** | **Rank 13** | **Rank 14** |
| CW_A_BII | 0.006 | 0.023 | 0.048 | 0.070 | 0.087 | 0.099 | 0.108 | 0.119 | 0.131 | 0.110 | 0.100 | 0.073 | 0.024 | 0.003 |
| CW_A_BII_B | 0.267 | 0.125 | 0.084 | 0.085 | 0.065 | 0.052 | 0.045 | 0.039 | 0.043 | 0.047 | 0.043 | 0.043 | 0.031 | 0.032 |
| CW_A_RY | 0.001 | 0.005 | 0.010 | 0.015 | 0.020 | 0.026 | 0.039 | 0.046 | 0.064 | 0.100 | 0.125 | 0.158 | 0.218 | 0.175 |
| CW_A_RY_B | 0.040 | 0.037 | 0.036 | 0.046 | 0.042 | 0.044 | 0.045 | 0.049 | 0.054 | 0.073 | 0.090 | 0.110 | 0.156 | 0.180 |
| CW_R_BII | 0.060 | 0.086 | 0.085 | 0.086 | 0.091 | 0.083 | 0.084 | 0.082 | 0.080 | 0.074 | 0.072 | 0.053 | 0.036 | 0.029 |
| CW_R_BII_B | 0.089 | 0.080 | 0.084 | 0.050 | 0.035 | 0.034 | 0.034 | 0.033 | 0.035 | 0.044 | 0.051 | 0.076 | 0.089 | 0.269 |
| PP_A_BII | 0.011 | 0.031 | 0.072 | 0.118 | 0.163 | 0.168 | 0.164 | 0.124 | 0.081 | 0.034 | 0.024 | 0.008 | 0.002 | 0.000 |
| PP_A_BII_B | 0.092 | 0.109 | 0.121 | 0.081 | 0.063 | 0.059 | 0.052 | 0.058 | 0.054 | 0.065 | 0.067 | 0.096 | 0.055 | 0.029 |
| PP_R_BII | 0.001 | 0.010 | 0.020 | 0.039 | 0.063 | 0.091 | 0.112 | 0.136 | 0.145 | 0.134 | 0.107 | 0.078 | 0.049 | 0.016 |
| PP_R_BII_B | 0.123 | 0.142 | 0.104 | 0.072 | 0.051 | 0.038 | 0.041 | 0.044 | 0.048 | 0.048 | 0.063 | 0.073 | 0.107 | 0.047 |
| PR_A_BII | 0.021 | 0.062 | 0.093 | 0.106 | 0.114 | 0.113 | 0.106 | 0.087 | 0.086 | 0.077 | 0.067 | 0.037 | 0.026 | 0.005 |
| PR_A_BII_B | 0.106 | 0.112 | 0.081 | 0.074 | 0.068 | 0.058 | 0.054 | 0.056 | 0.053 | 0.062 | 0.062 | 0.059 | 0.077 | 0.078 |
| PR_A_RY | 0.111 | 0.097 | 0.082 | 0.085 | 0.069 | 0.064 | 0.051 | 0.059 | 0.058 | 0.060 | 0.062 | 0.069 | 0.066 | 0.068 |
| PR_R_BII | 0.074 | 0.081 | 0.082 | 0.076 | 0.070 | 0.071 | 0.068 | 0.068 | 0.068 | 0.072 | 0.070 | 0.067 | 0.064 | 0.070 |

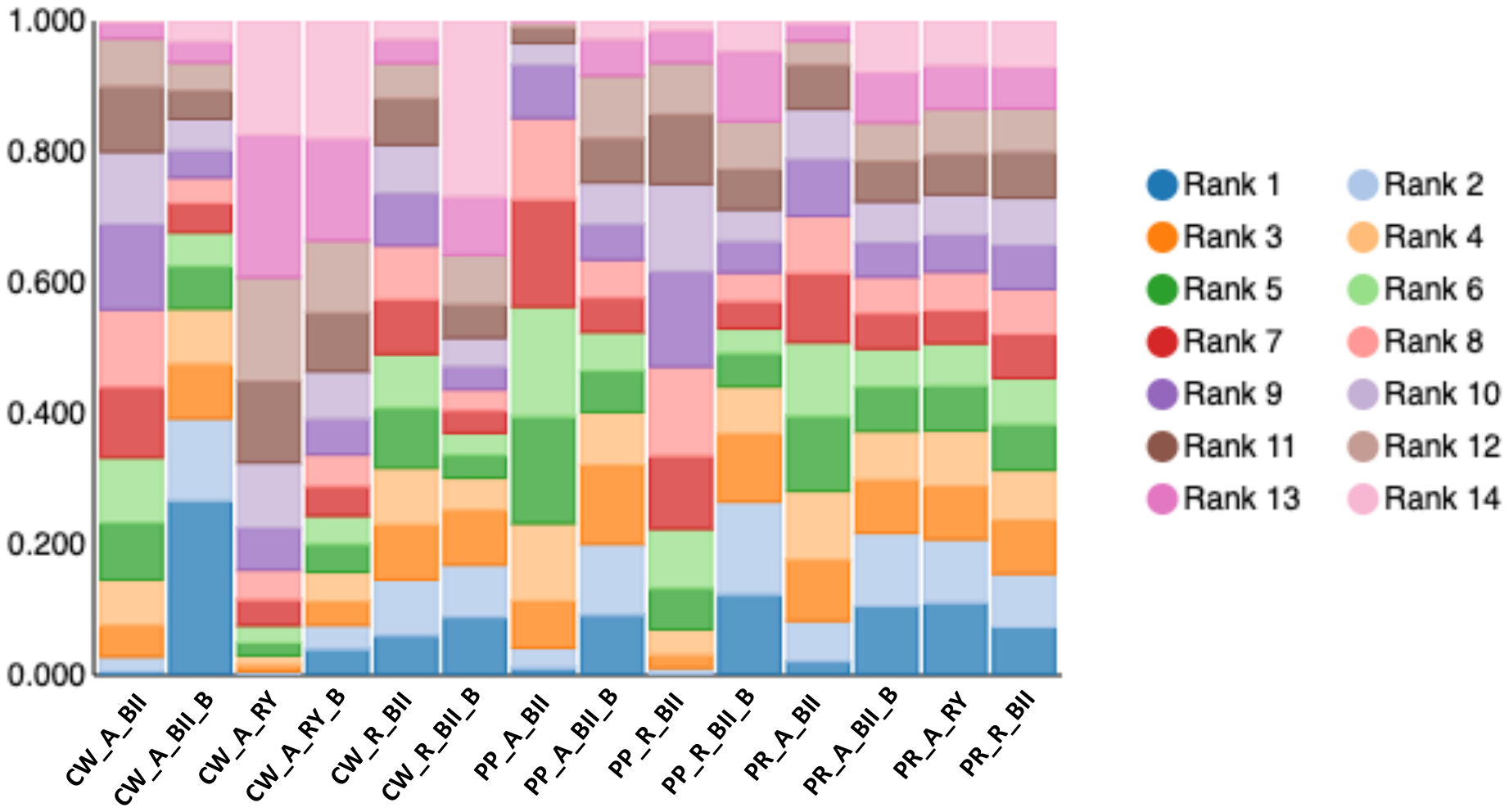

Length of stay (days)

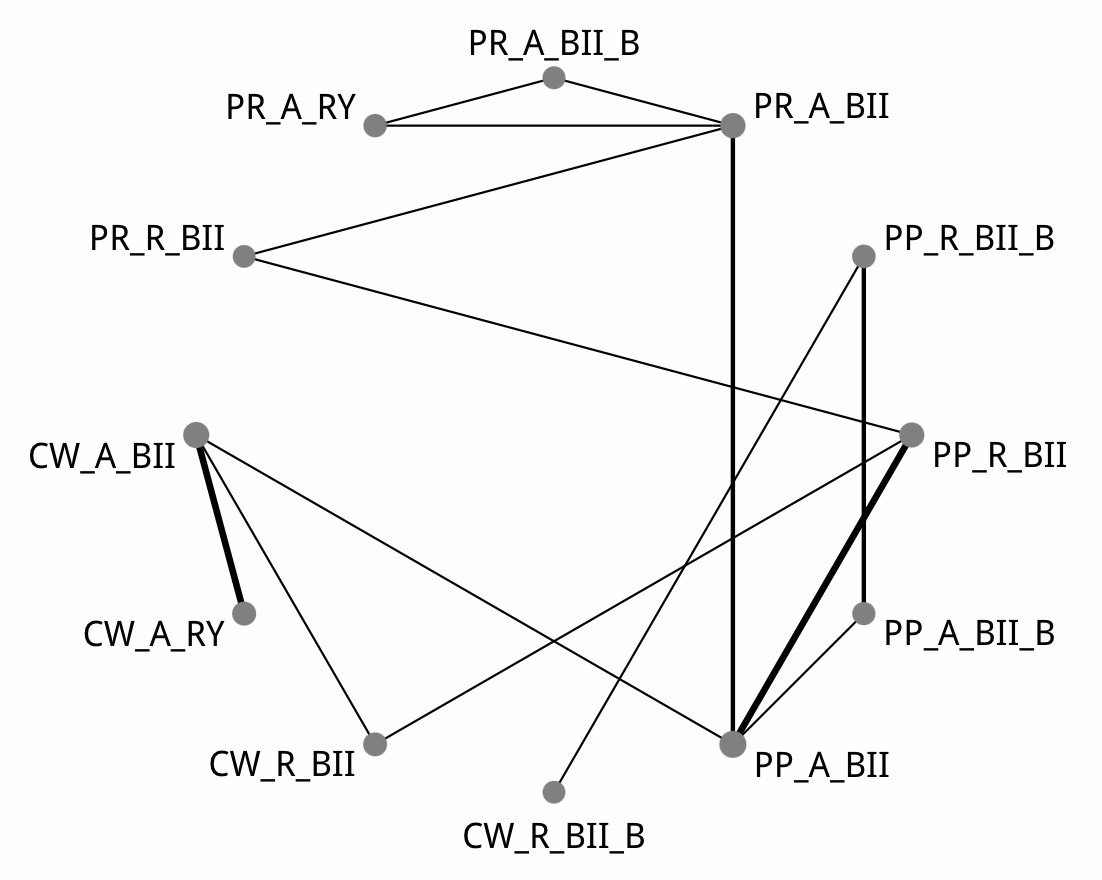

| Comparison of the included interventions: mean difference (95% CrI). Each cell gives the effect of the column-defining intervention relative to the row-defining intervention. | | | | | | | | | | | |
| --- | --- | --- | --- | --- | --- | --- | --- | --- | --- | --- | --- |
| CW_A_BII | -0.445 ( -3.788, 2.214) | 0.560 ( -5.040, 6.548) | -11.341 ( -24.496, 2.132) | -2.204 ( -8.996, 4.477) | -3.292 ( -12.242, 6.005) | -2.327 ( -8.633, 4.812) | -9.778 ( -20.640, 1.353) | -3.224 ( -10.605, 4.747) | -3.213 ( -13.258, 8.607) | 1.768 ( -7.575, 12.482) | 1.299 ( -8.032, 10.894) |
|  | CW_A_RY | 1.047 ( -5.086, 7.706) | -10.875 ( -23.936, 3.114) | -1.785 ( -8.889, 5.601) | -2.828 ( -12.306, 7.149) | -1.888 ( -8.735, 6.126) | -9.309 ( -20.446, 2.596) | -2.794 ( -10.556, 5.878) | -2.735 ( -13.266, 9.437) | 2.251 ( -7.483, 13.713) | 1.758 ( -7.645, 11.791) |
|  |  | CW_R_BII | -11.837 ( -24.660, 1.056) | -2.829 ( -8.817, 2.660) | -3.840 ( -12.885, 4.573) | -2.842 ( -7.778, 2.661) | -10.243 ( -21.083, 0.021) | -3.755 ( -10.348, 3.037) | -3.616 ( -12.880, 6.829) | 1.223 ( -7.371, 10.908) | 0.732 ( -7.759, 9.438) |
|  |  |  | CW_R_BII_B | 9.046 ( -2.516, 20.143) | 8.064 ( -1.569, 17.180) | 9.009 ( -2.817, 21.059) | 1.554 ( -5.573, 8.530) | 8.090 ( -3.688, 20.110) | 8.200 ( -5.293, 22.552) | **13.142 ( 0.124, 27.162)** | 12.698 ( -0.298, 26.169) |
|  |  |  |  | PP_A_BII | -1.093 ( -7.442, 5.350) | -0.127 ( -2.770, 4.143) | -7.515 ( -16.301, 1.621) | -0.969 ( -4.441, 3.240) | -0.802 ( -8.723, 8.341) | 3.963 ( -2.415, 12.174) | 3.612 ( -2.860, 10.162) |
|  |  |  |  |  | PP_A_BII_B | 1.036 ( -5.718, 8.784) | -6.400 ( -12.496, -0.396) | 0.149 ( -7.161, 7.562) | 0.289 ( -9.841, 11.308) | 5.033 ( -4.205, 15.344) | 4.577 ( -4.668, 13.711) |
|  |  |  |  |  |  | PP_R_BII | -7.469 ( -17.393, 1.943) | -0.869 ( -5.998, 3.639) | -0.833 ( -9.158, 8.195) | 4.047 ( -3.219, 12.272) | 3.672 ( -3.837, 10.211) |
|  |  |  |  |  |  |  | PP_R_BII_B | 6.633 ( -3.094, 16.047) | 6.657 ( -5.349, 19.027) | 11.571 ( 0.462, 23.765) | 11.179 ( 0.118, 22.131) |
|  |  |  |  |  |  |  |  | PR_A_BII | 0.160 ( -7.216, 7.981) | 4.968 ( -0.858, 11.624) | 4.491 ( -1.310, 10.432) |
|  |  |  |  |  |  |  |  |  | PR_A_BII_B | 4.935 ( -1.866, 11.519) | 4.336 ( -5.143, 13.562) |
|  |  |  |  |  |  |  |  |  |  | PR_A_RY | -0.518 ( -9.007, 7.496) |
|  |  |  |  |  |  |  |  |  |  |  | PR_R_BII |

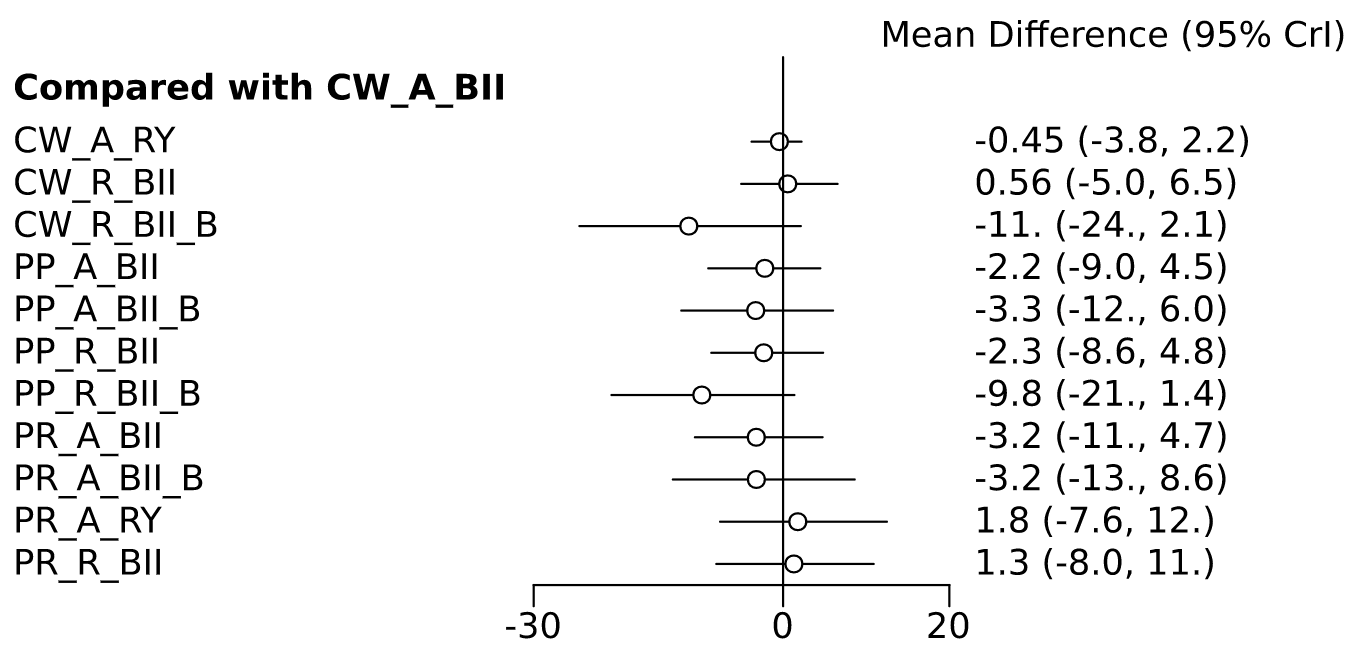

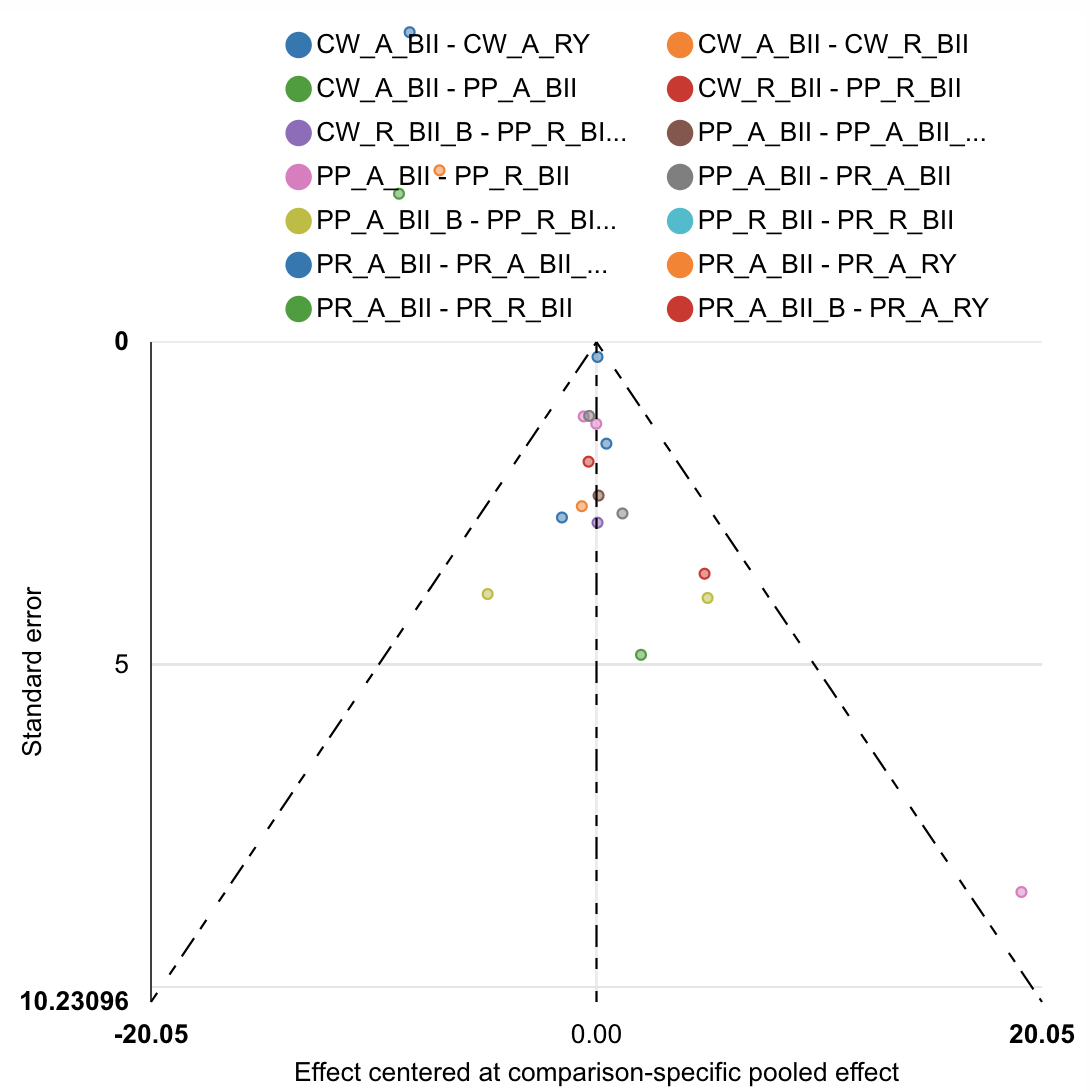

| Rank probabilities table | | | | | | | | | | | | |
| --- | --- | --- | --- | --- | --- | --- | --- | --- | --- | --- | --- | --- |
|  | **Rank 1** | **Rank 2** | **Rank 3** | **Rank 4** | **Rank 5** | **Rank 6** | **Rank 7** | **Rank 8** | **Rank 9** | **Rank 10** | **Rank 11** | **Rank 12** |
| CW_A_BII | 0.005 | 0.012 | 0.021 | 0.059 | 0.057 | 0.061 | 0.072 | 0.107 | 0.160 | 0.172 | 0.167 | 0.109 |
| CW_A_RY | 0.016 | 0.020 | 0.065 | 0.068 | 0.067 | 0.062 | 0.087 | 0.133 | 0.150 | 0.154 | 0.126 | 0.053 |
| CW_R_BII | 0.002 | 0.004 | 0.014 | 0.022 | 0.029 | 0.047 | 0.070 | 0.117 | 0.165 | 0.215 | 0.176 | 0.140 |
| CW_R_BII_B | 0.631 | 0.233 | 0.046 | 0.022 | 0.016 | 0.010 | 0.010 | 0.008 | 0.008 | 0.004 | 0.006 | 0.006 |
| PP_A_BII | 0.002 | 0.007 | 0.032 | 0.098 | 0.187 | 0.244 | 0.217 | 0.117 | 0.059 | 0.030 | 0.008 | 0.001 |
| PP_A_BII_B | 0.004 | 0.016 | 0.306 | 0.155 | 0.106 | 0.086 | 0.086 | 0.071 | 0.065 | 0.046 | 0.035 | 0.025 |
| PP_R_BII | 0.006 | 0.014 | 0.064 | 0.131 | 0.178 | 0.200 | 0.174 | 0.113 | 0.072 | 0.031 | 0.013 | 0.004 |
| PP_R_BII_B | 0.253 | 0.588 | 0.073 | 0.024 | 0.017 | 0.013 | 0.012 | 0.007 | 0.006 | 0.005 | 0.002 | 0.001 |
| PR_A_BII | 0.013 | 0.034 | 0.150 | 0.248 | 0.216 | 0.130 | 0.087 | 0.058 | 0.040 | 0.019 | 0.003 | 0.000 |
| PR_A_BII_B | 0.064 | 0.064 | 0.208 | 0.131 | 0.075 | 0.071 | 0.091 | 0.062 | 0.062 | 0.081 | 0.071 | 0.021 |
| PR_A_RY | 0.001 | 0.004 | 0.009 | 0.017 | 0.027 | 0.036 | 0.042 | 0.102 | 0.093 | 0.113 | 0.197 | 0.361 |
| PR_R_BII | 0.002 | 0.005 | 0.012 | 0.026 | 0.027 | 0.042 | 0.054 | 0.104 | 0.120 | 0.132 | 0.197 | 0.280 |

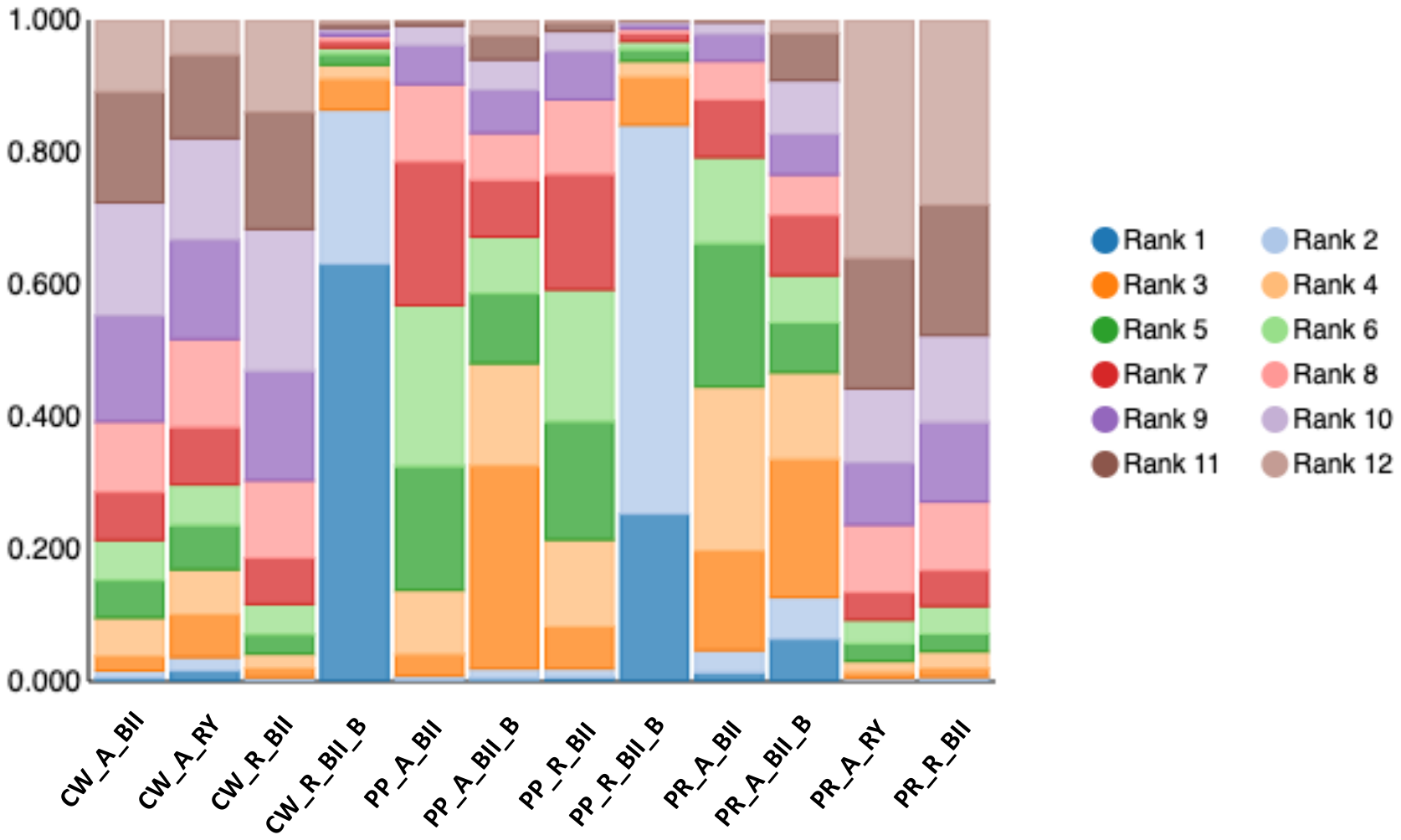

**Excluding Braun Enteroenterostomy Comparison Studies**

DGE Overall

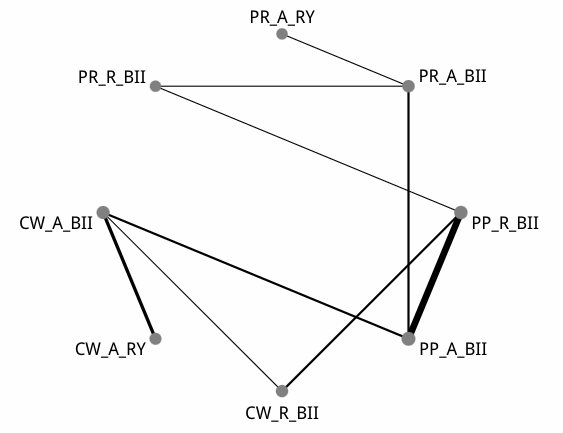

| Comparison of the included interventions: odds ratio (95% CrI). Each cell gives the effect of the column-defining intervention relative to the row-defining intervention. | | | | | | | |
| --- | --- | --- | --- | --- | --- | --- | --- |
| CW_A_BII | 0.815 ( 0.246, 2.552) | 3.172 ( 0.791, 17.549) | 1.081 ( 0.310, 4.342) | 1.982 ( 0.551, 10.068) | 0.593 ( 0.094, 4.006) | 0.765 ( 0.046, 12.967) | 1.514 ( 0.191, 15.076) |
|  | CW_A_RY | 3.897 ( 0.670, 32.308) | 1.323 ( 0.245, 8.286) | 2.417 ( 0.454, 19.430) | 0.734 ( 0.085, 6.555) | 0.969 ( 0.044, 18.818) | 1.809 ( 0.174, 23.531) |
|  |  | CW_R_BII | 0.339 ( 0.074, 1.301) | 0.622 ( 0.172, 2.360) | 0.187 ( 0.023, 1.081) | 0.242 ( 0.011, 3.475) | 0.473 ( 0.054, 3.923) |
|  |  |  | PP_A_BII | 1.843 ( 0.853, 4.698) | 0.562 ( 0.139, 1.958) | 0.721 ( 0.052, 7.708) | 1.415 ( 0.239, 8.444) |
|  |  |  |  | PP_R_BII | 0.302 ( 0.054, 1.167) | 0.384 ( 0.023, 4.364) | 0.763 ( 0.126, 3.946) |
|  |  |  |  |  | PR_A_BII | 1.300 ( 0.150, 10.299) | 2.534 ( 0.487, 15.605) |
|  |  |  |  |  |  | PR_A_RY | 1.996 ( 0.144, 33.852) |
|  |  |  |  |  |  |  | PR_R_BII |

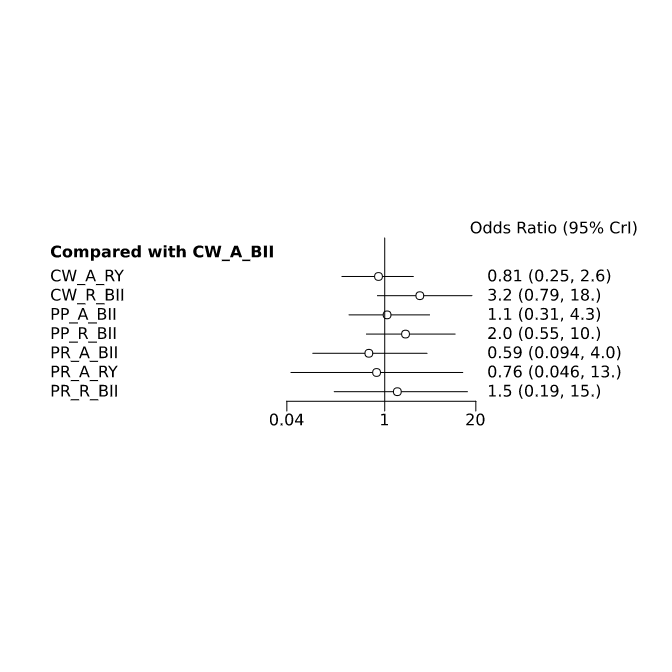

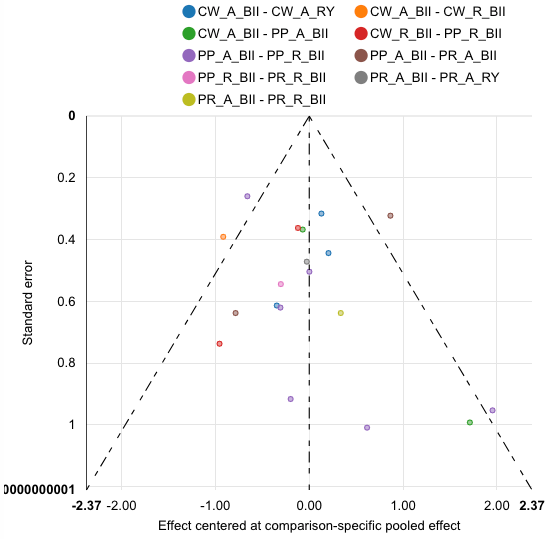

| Rank probabilities table | | | | | | | | |
| --- | --- | --- | --- | --- | --- | --- | --- | --- |
|  | **Rank 1** | **Rank 2** | **Rank 3** | **Rank 4** | **Rank 5** | **Rank 6** | **Rank 7** | **Rank 8** |
| CW_A_BII | 0.066 | 0.166 | 0.182 | 0.213 | 0.182 | 0.119 | 0.060 | 0.012 |
| CW_A_RY | 0.246 | 0.166 | 0.167 | 0.139 | 0.113 | 0.080 | 0.061 | 0.028 |
| CW_R_BII | 0.002 | 0.005 | 0.014 | 0.022 | 0.046 | 0.100 | 0.206 | 0.605 |
| PP_A_BII | 0.029 | 0.093 | 0.226 | 0.257 | 0.245 | 0.128 | 0.020 | 0.004 |
| PP_R_BII | 0.001 | 0.006 | 0.022 | 0.055 | 0.137 | 0.282 | 0.393 | 0.105 |
| PR_A_BII | 0.312 | 0.322 | 0.165 | 0.115 | 0.049 | 0.024 | 0.010 | 0.003 |
| PR_A_RY | 0.295 | 0.168 | 0.106 | 0.088 | 0.090 | 0.081 | 0.083 | 0.088 |
| PR_R_BII | 0.049 | 0.075 | 0.120 | 0.111 | 0.138 | 0.186 | 0.166 | 0.156 |

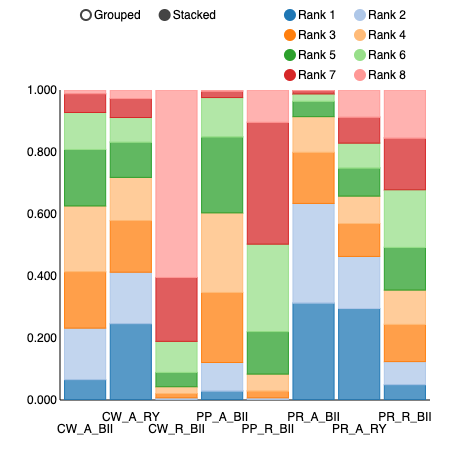

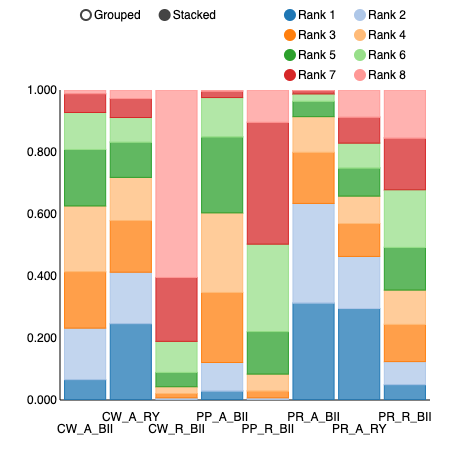

**Rank probability**

Network meta regression to assess impact of Braun enteroenterostomy

No Braun:

| Rank probabilities table | | | | | | | | |
| --- | --- | --- | --- | --- | --- | --- | --- | --- |
|  | **Rank 1** | **Rank 2** | **Rank 3** | **Rank 4** | **Rank 5** | **Rank 6** | **Rank 7** | **Rank 8** |
| CW_A_BII | 0.010 | 0.054 | 0.122 | 0.181 | 0.213 | 0.185 | 0.170 | 0.065 |
| CW_A_RY | 0.028 | 0.062 | 0.085 | 0.114 | 0.130 | 0.168 | 0.168 | 0.246 |
| CW_R_BII | 0.591 | 0.214 | 0.106 | 0.046 | 0.024 | 0.013 | 0.004 | 0.003 |
| PP_A_BII | 0.005 | 0.024 | 0.127 | 0.253 | 0.261 | 0.217 | 0.085 | 0.029 |
| PP_R_BII | 0.110 | 0.393 | 0.280 | 0.133 | 0.058 | 0.019 | 0.007 | 0.002 |
| PR_A_BII | 0.003 | 0.010 | 0.023 | 0.052 | 0.112 | 0.171 | 0.326 | 0.304 |
| PR_A_RY | 0.092 | 0.074 | 0.077 | 0.088 | 0.091 | 0.109 | 0.167 | 0.302 |
| PR_R_BII | 0.161 | 0.170 | 0.181 | 0.134 | 0.112 | 0.118 | 0.074 | 0.050 |

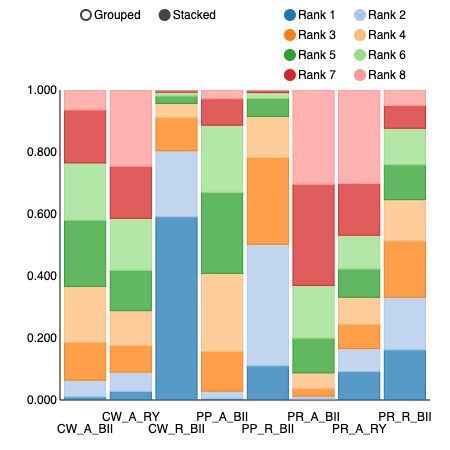

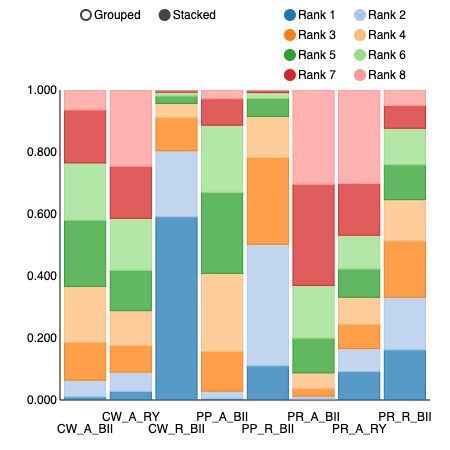

**Rank probability**

Braun:

| Rank probabilities table | | | | | | | | |
| --- | --- | --- | --- | --- | --- | --- | --- | --- |
|  | **Rank 1** | **Rank 2** | **Rank 3** | **Rank 4** | **Rank 5** | **Rank 6** | **Rank 7** | **Rank 8** |
| CW_A_BII | 0.325 | 0.056 | 0.045 | 0.042 | 0.042 | 0.049 | 0.067 | 0.375 |
| CW_A_RY | 0.020 | 0.051 | 0.092 | 0.125 | 0.166 | 0.173 | 0.209 | 0.164 |
| CW_R_BII | 0.400 | 0.335 | 0.140 | 0.071 | 0.031 | 0.014 | 0.006 | 0.002 |
| PP_A_BII | 0.003 | 0.019 | 0.100 | 0.254 | 0.308 | 0.213 | 0.086 | 0.019 |
| PP_R_BII | 0.072 | 0.297 | 0.339 | 0.192 | 0.073 | 0.020 | 0.006 | 0.001 |
| PR_A_BII | 0.002 | 0.008 | 0.019 | 0.048 | 0.124 | 0.258 | 0.346 | 0.198 |
| PR_A_RY | 0.066 | 0.074 | 0.078 | 0.097 | 0.114 | 0.153 | 0.211 | 0.208 |
| PR_R_BII | 0.112 | 0.162 | 0.188 | 0.173 | 0.143 | 0.121 | 0.069 | 0.033 |

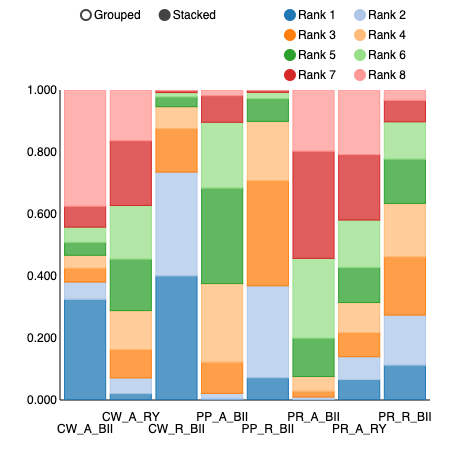

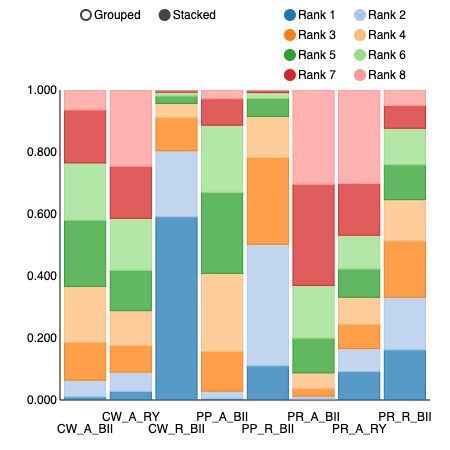

**Rank probability**

**Separated meta-analysis**

**Gastric Resection**

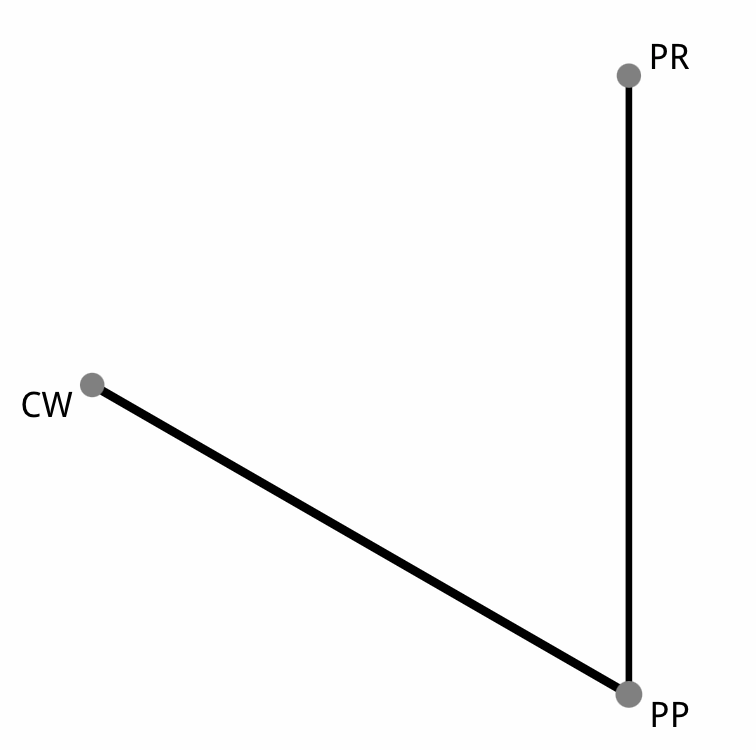

| Comparison of the included interventions: odds ratio (95% CrI). Each cell gives the effect of the column-defining intervention relative to the row-defining intervention. | | |
| --- | --- | --- |
| CW | 0.816 ( 0.243, 3.296) | 0.491 ( 0.070, 3.479) |
|  | PP | 0.609 ( 0.133, 2.412) |
|  |  | PR |

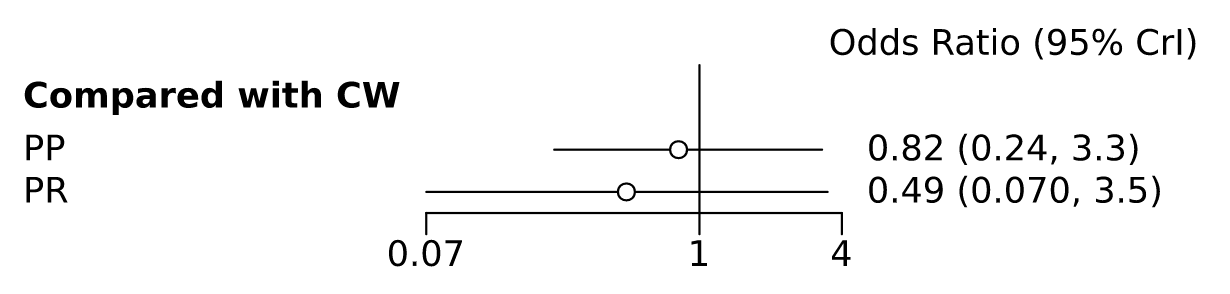

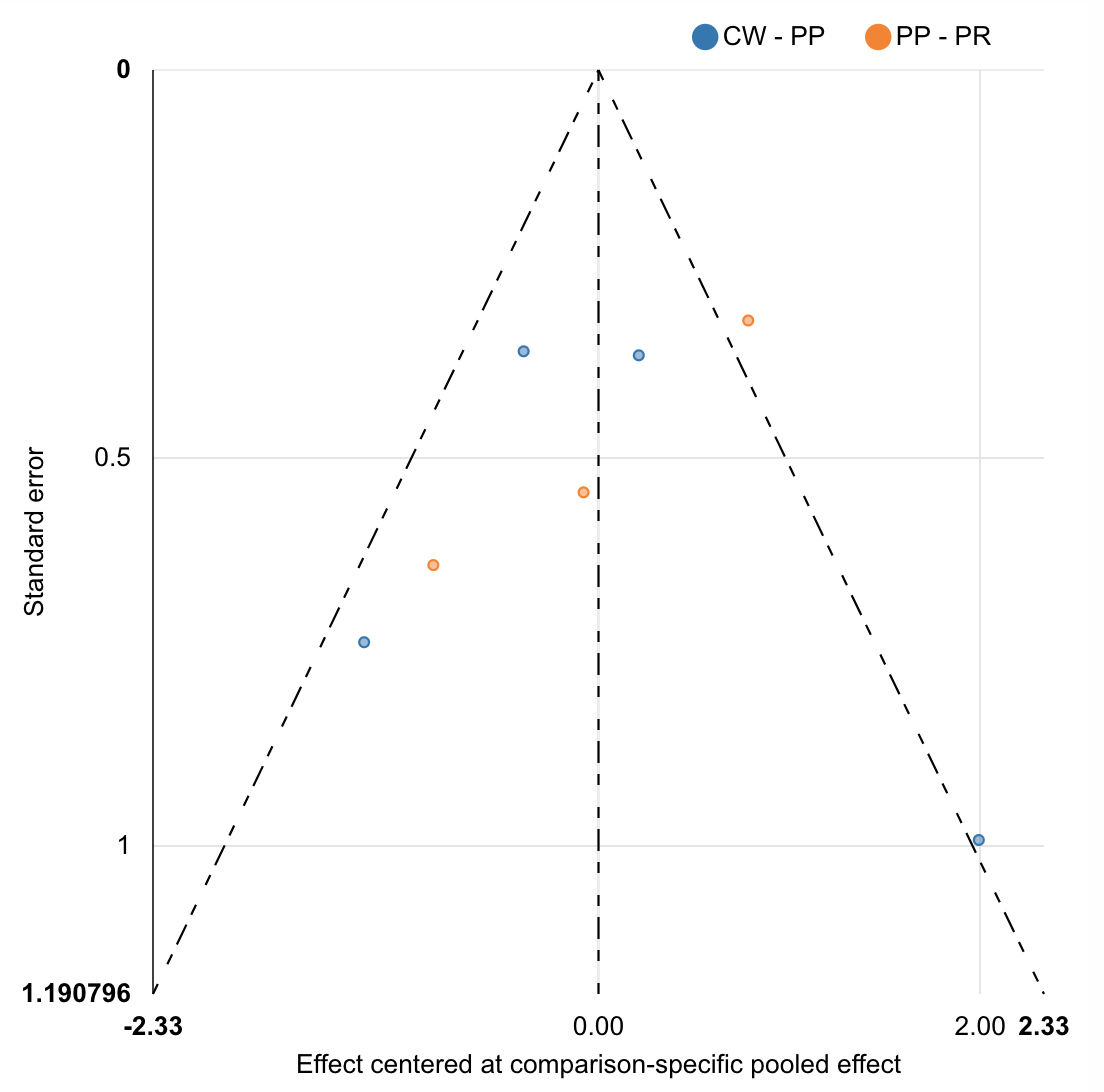

| Rank probabilities table | | | |
| --- | --- | --- | --- |
|  | **Rank 1** | **Rank 2** | **Rank 3** |
| CW | 0.157 | 0.254 | 0.590 |
| PP | 0.129 | 0.584 | 0.287 |
| PR | 0.714 | 0.162 | 0.124 |

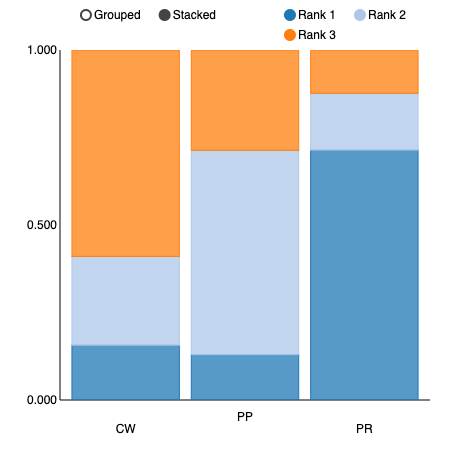

**Antecolic vs Retrocolic Route of Gastro- or duodenojejunostomy**

| Comparison of the included interventions: odds ratio (95% CrI). Each cell gives the effect of the column-defining intervention relative to the row-defining intervention. | |
| --- | --- |
| A | 2.013 ( 1.035, 5.182) |
|  | R |

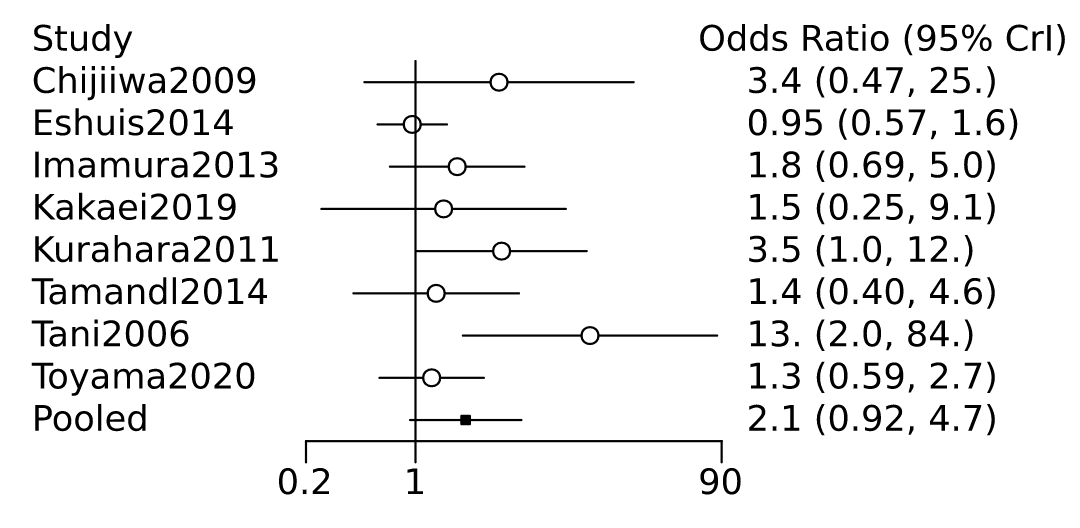

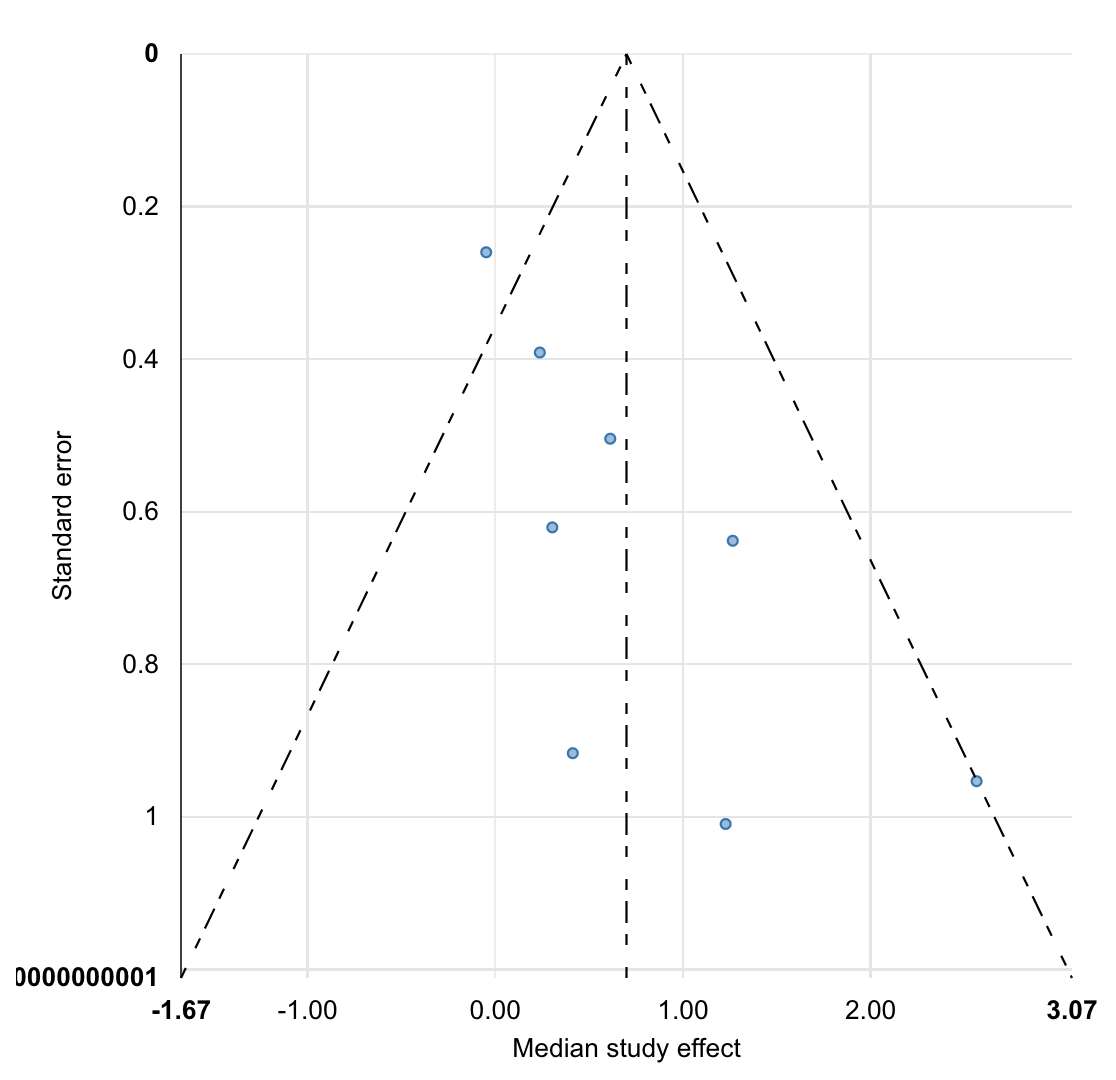

| Rank probabilities table | | |
| --- | --- | --- |
|  | **Rank 1** | **Rank 2** |
| A | 0.979 | 0.021 |
| R | 0.021 | 0.979 |

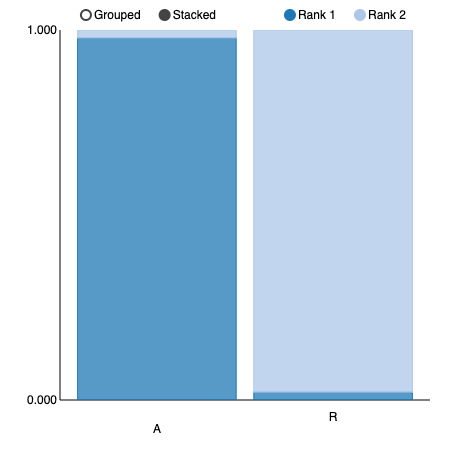

**Anastomotic Configuration – Billroth II vs Roux-en-Y**

| Comparison of the included interventions: odds ratio (95% CrI). Each cell gives the effect of the column-defining intervention relative to the row-defining intervention. | |
| --- | --- |
| BII | 1.145 ( 0.591, 2.391) |
|  | RY |

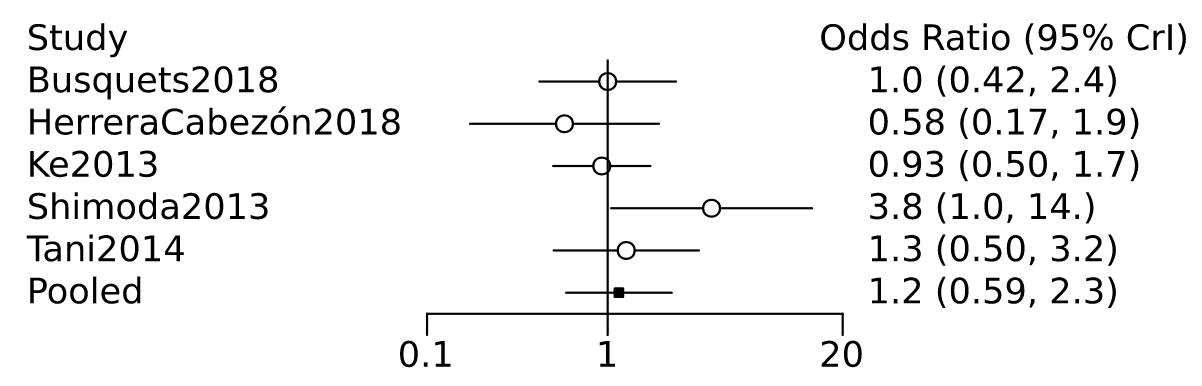

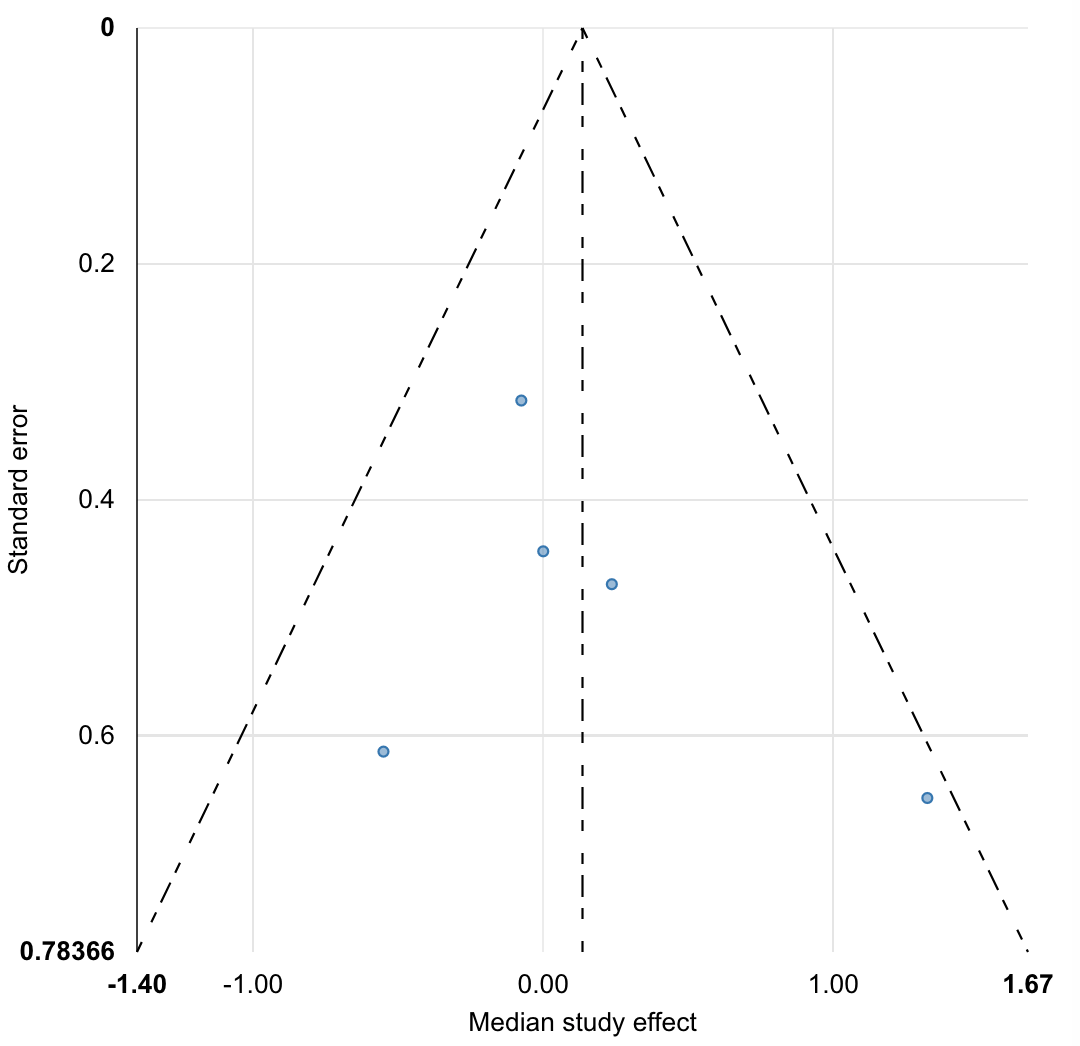

| Rank probabilities table | | |
| --- | --- | --- |
|  | **Rank 1** | **Rank 2** |
| BII | 0.677 | 0.323 |
| RY | 0.323 | 0.677 |

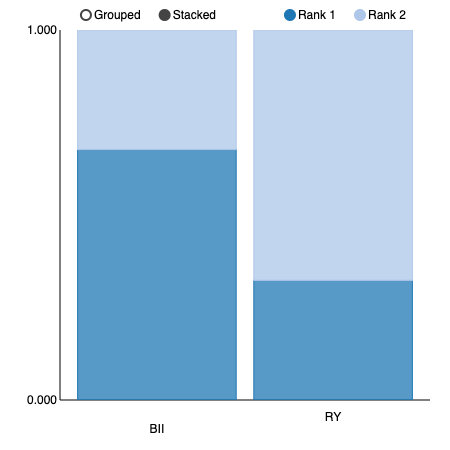

**Braun Enteroenterostomy**

| Comparison of the included interventions: odds ratio (95% CrI). Each cell gives the effect of the column-defining intervention relative to the row-defining intervention. | |
| --- | --- |
| Braun | 1.874 ( 0.924, 4.023) |
|  | Control |

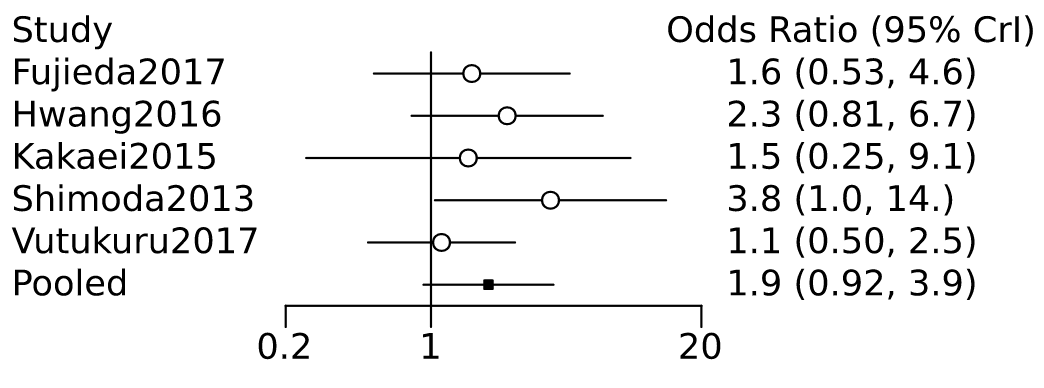

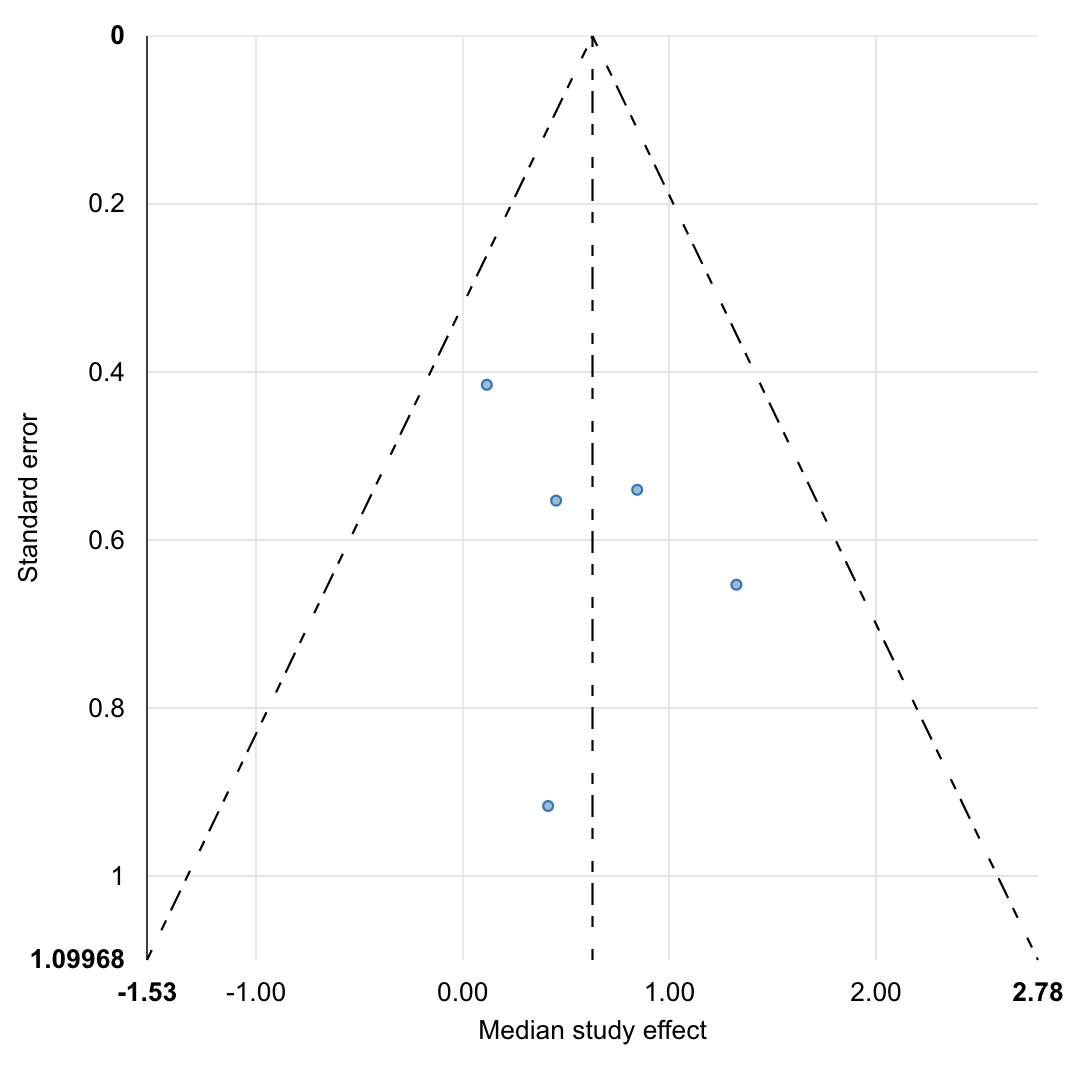

| Rank probabilities table | | |
| --- | --- | --- |
|  | **Rank 1** | **Rank 2** |
| Braun | 0.963 | 0.037 |
| Control | 0.037 | 0.963 |

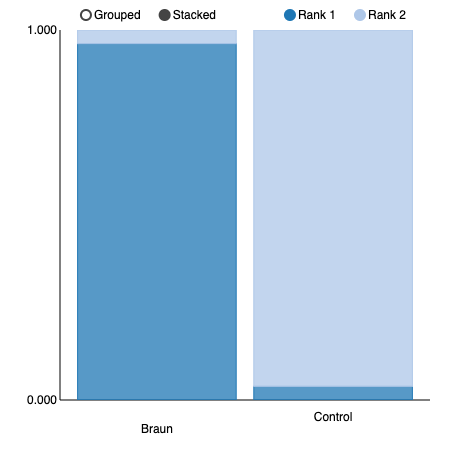

### Appendix S4: Heterogeneity and inconsistency

DGE overall

Random effects standard deviation 0.899 (0.335, 1.831); some heterogeneity. All node-splitting inconsistency p values were >0.05.

POPF overall

Random effects standard deviation 0.221 (0.008, 0.748); no heterogeneity. All node-splitting inconsistency p values were >0.05.

POH

Random effects standard deviation 0.390 (0.020, 1.081); some heterogeneity. Node-splitting analysis not possible for this model due to limited dataset.

Intraoperative blood loss

Random effects standard deviation 49.374 (2.405, 221.370); some heterogeneity. All node-splitting inconsistency p values were >0.05.

Duration of operation

Random effects standard deviation 6.836 (0.246, 23.471); no heterogeneity. All node-splitting inconsistency p values were >0.05.

Intra-abdominal abscess

Random effects standard deviation 0.199 (0.009, 0.858); no heterogeneity. All node-splitting inconsistency p values were >0.05.

Wound infection

Random effects standard deviation 0.476 (0.026, 1.360); some heterogeneity. All node-splitting inconsistency p values were >0.05.

Bile leak

Random effects standard deviation 0.456 (0.013, 1.108); some heterogeneity. Node-splitting analysis not possible for this model due to limited dataset.

Reoperation

Random effects standard deviation 0.306 (0.014, 0.815); no heterogeneity. All node-splitting inconsistency p values were >0.05.

Mortality

Random effects standard deviation 0.293 (0.020, 0.833); no heterogeneity. All node-splitting inconsistency p values were >0.05.

Length of stay

Random effects standard deviation 1.052 (0.051, 5.767); no heterogeneity. On node-splitting analysis, inconsistency was found in comparison of PR_A_BII vs PR_A_BII_B (inconsistency factor 16.1, 95% CI; 2.3 to 28.7, p = 0.024), PR_A_BII vs PR_A_RY (inconsistency factor -16.5, 95% CI; -30.3 to -1.9, p = 0.024) and PR_A_BII_B vs PR_A_RY (inconsistency factor 16.0, 95% CI; 1.8 to 29.7, p = 0.025). All other node-splitting inconsistency p values were >0.05.

No braun DGE overall

Random effects standard deviation 0.804 (0.264, 1.644); some heterogeneity. All node-splitting inconsistency p values were >0.05.

Gastric resection

Random effects standard deviation 1.037 (0.240, 1.735); high heterogeneity. Node-splitting analysis not possible for this model due to limited dataset.

A vs R

Random effects standard deviation 0.728 (0.101, 1.864); no heterogeneity.

BII vs RY

Random effects standard deviation 0.437 (0.020, 1.227); no heterogeneity.

Braun vs no Braun

Random effects standard deviation 0.399 (0.017, 1.198); no heterogeneity.

### Figure S1: Risk of bias summary graph

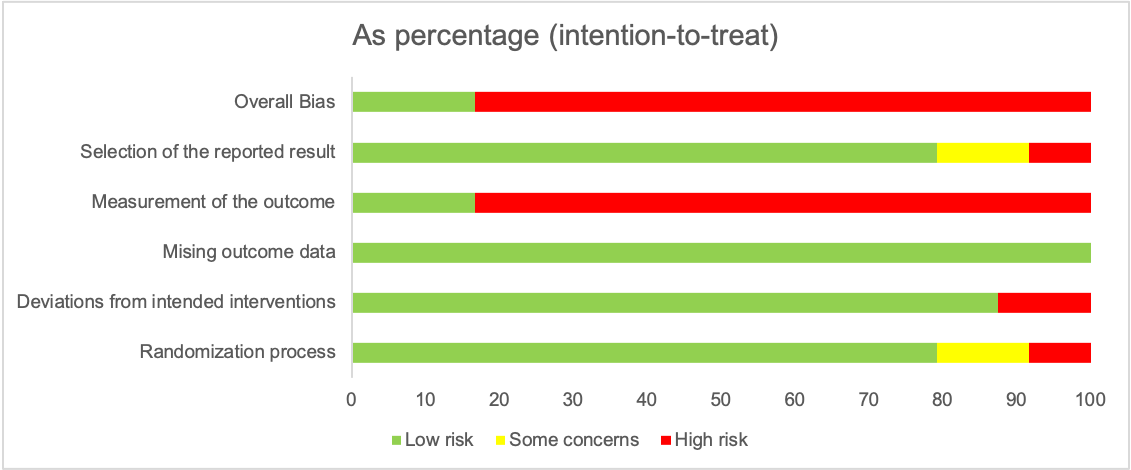

### Figure S2: Risk of bias figure

Green positive = low risk of bias, red negative = high risk of bias, yellow question mark = unclear risk of bias.

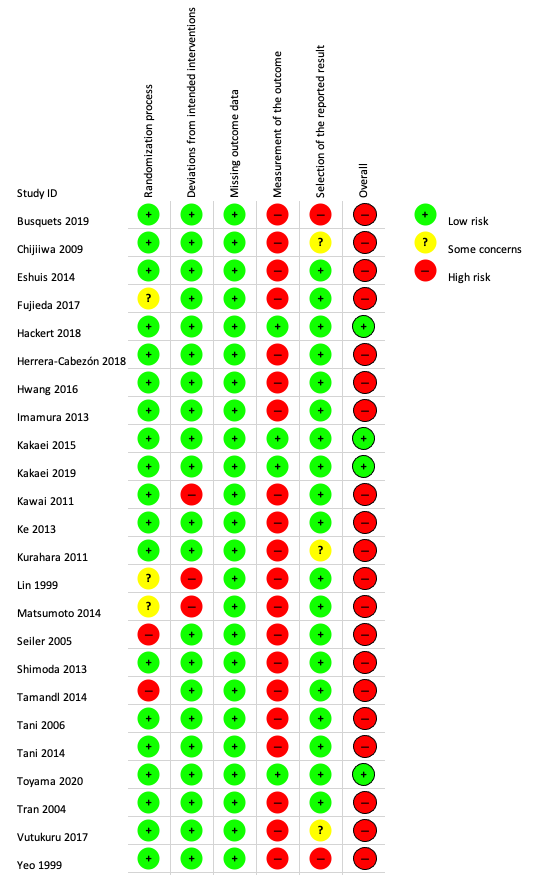
