## Supplementary material for "Impact of gastric resection and enteric anastomotic configuration on delayed gastric emptying after pancreaticoduodenectomy: a network meta-analysis of randomized trials": Table 1

Table 1. Summary of included trials

|  |  |  |  |  |  |  | **Trial Location** | | |
| --- | --- | --- | --- | --- | --- | --- | --- | --- | --- |
| **Trial Comparison** | **No. of trials** | **No. of patients**^†^ | **Publication Years** | **Age (years)** | **Women (%)** | **BMI (kg/m2)** | **Asia** | **North America** | **Europe** |
| PP_A_BII vs PP_R_BII | 4 | 393 | 2006-2019 | 62.1 | 42.1 | 25 | 2 |  | 2 |
| CW_A_BII vs CW_A_RY | 3 | 360 | 2013-2018 | 66.9 | 64.2 | 24.7 | 1 |  | 2 |
| CW_A_BII vs PP_A_BII | 2 | 201 | 1999-2004 | 59.7 | 37.3 |  | 1 |  | 1 |
| PP_A_BII vs PR_A_BII | 2 | 328 | 2011-2018 | 65.4 | 45 | 25.5 | 1 |  | 1 |
| PP_A_BII_B vs PP_R_BII_B | 2 | 155 | 2009-2013 | 67.4 | 41.7 | 21.9 | 2 |  |  |
| CW_A_BII_B vs CW_A_BII | 1 | 30 | 2015 | 56.3 | 33.3 | NA | 1 |  |  |
| CW_A_RY vs CW_A_RY_B | 1 | 104 | 2017 | 53.9 | 38.5 | NA | 1 |  |  |
| CW_R_BII vs CW_A_BII | 1 | 214 | 2020 | NA | 38.2 | 21.8 | 1 |  |  |
| CW_R_BII_B vs PP_R_BII_B | 1 | 214 | 2005 | 61 | 46.9 | NA |  |  | 1 |
| PP_A_BII_B vs PP_A_BII | 1 | 60 | 2016 | 66 | 36.7 | NA | 1 |  |  |
| PP_R_BII vs CW_R_BII | 1 | 114 | 1999 | 65 | 46.5 | NA |  | 1 |  |
| PP_R_BII vs PR_R_BII | 1 | 106 | 2014 | 66.5 | 36 | 22.8 | 1 |  |  |
| PR_A_BII vs PR_A_BII_B | 1 | 68 | 2017 | 67.1 | 35.3 | 21.7 | 1 |  |  |
| PR_A_BII vs PR_A_RY | 1 | 153 | 2014 | 68.8 | 46.4 | NA | 1 |  |  |
| PR_A_BII_B vs PR_A_RY | 1 | 101 | 2013 | 66.1 | 40.6 | 21.5 | 1 |  |  |
| PR_R_BII vs PR_A_BII | 1 | 46 | 2011 | NA | 29.2 | 23.1 | 1 |  |  |

*Values are mean (SD). PP, pylorus preserving; CW, classic Whipple; PR, pylorus resecting; A, antecolic; R, retrocolic; BII, Billroth II; RY, Roux-en-Y; B, Braun enteroenterostomy.

†Number of patients randomized and used in intention-to-treat analysis
