## Supplementary material for "Impact of gastric resection and enteric anastomotic configuration on delayed gastric emptying after pancreaticoduodenectomy: a network meta-analysis of randomized trials": Table 3

**Table 3. Rank probability of being the best ranked approach for pancreaticoduodenectomy in direct and indirect comparison in the network meta-analysis**

| **Rank probability of being first ranked technique of closure** | | | | | | | | | | | | | |
| --- | --- | --- | --- | --- | --- | --- | --- | --- | --- | --- | --- | --- | --- |
|  | **Overall DGE** | **Overall POPF** | **Overall POPF*** | **Intra-abdominal abscess** | **Intra-abdominal abscess*** | **Would infection** | **Bile Leak** | **Postpancreatectomy haemorrhage** | **Duration of operation** | **Intraoperative blood loss** | **Reoperation** | **Mortality** | **Length of stay** |
| CW_A_BII | 0.005 | 0.001 | 0.001 | 0.012 | 0.026 | 0.01 | 0.016 | 0.001 | 0 | 0.003 | 0.001 | 0.006 | 0.005 |
| CW_A_BII_B | 0.215 |  |  |  |  | 0.048 | 0.285 | 0.273 | 0.048 | 0.216 |  | 0.267† |  |
| CW_A_RY | 0.027 | 0.001 | 0.004 | 0.016 | 0.032 | 0.042 | 0.138 | 0.12 | 0.001 | 0.029 | 0.041 | 0.001 | 0.016 |
| CW_A_RY_B | 0.123 | 0.01 | 0.026 | 0.093 | 0.135 | 0.14 |  |  |  |  | 0.332 | 0.04 |  |
| CW_R_BII | 0.002 | 0.002 | 0.005 | 0.108 | 0.181 | 0.027 | 0.013 | 0.03 | 0.005 | 0.01 | 0.01 | 0.06 | 0.002 |
| CW_R_BII_B | 0.032 | 0.786† |  |  |  |  |  | 0.118 | 0.006 | 0.013 |  | 0.089 | 0.631† |
| PP_A_BII | 0.001 | 0 | 0.001 | 0.005 | 0.015 | 0.006 | 0.002 | 0.01 | 0.03 | 0.009 | 0.006 | 0.011 | 0.002 |
| PP_A_BII_B | 0.17 | 0.014 | 0.092 | 0.082 | 0.085 | 0.006 | 0.057 | 0.115 | 0.019 | 0.014 |  | 0.092 | 0.004 |
| PP_R_BII | 0.001 | 0.002 | 0.005 | 0.01 | 0.021 | 0.02 | 0 | 0.006 | 0.021 | 0.01 | 0.002 | 0.001 | 0.006 |
| PP_R_BII_B | 0.027 | 0.074 | 0.51† | 0.299 | 0.329† | 0.02 | 0.35† | 0.293† | 0.816† | 0.574† |  | 0.123 | 0.253 |
| PR_A_BII | 0.02 | 0.008 | 0.031 | 0.039 | 0.173 | 0.027 | 0.011 | 0.025 | 0.031 | 0.034 | 0.019 | 0.021 | 0.013 |
| PR_A_BII_B | 0.346† | 0.015 | 0.047 |  |  |  |  |  | 0.012 | 0.054 |  | 0.106 | 0.064 |
| PR_A_RY | 0.022 | 0.081 | 0.257 | 0.333† |  | 0.568† | 0.128 | 0.009 | 0.004 | 0.033 | 0.484† | 0.111 | 0.001 |
| PR_R_BII | 0.01 | 0.006 | 0.02 | 0.004 | 0.003 | 0.087 | 0.001 |  | 0.009 | 0.001 | 0.106 | 0.074 | 0.002 |

CW, Classic Whipple; PP, pylorus-preserving; PR, pylorus-resecting; A, antecolic; R, retrocolic; BII, Billroth II; RY, Roux-en-Y; B, Braun enteroenterostomy.

*Results of sensitivity analysis performed by omitting trial with single comparison. †PD approach with the highest probability of ranking first.
